# One in seven has severe pain: A point-prevalence study of hospitalised patients across Africa

**DOI:** 10.64898/2026.01.08.26343598

**Authors:** African Critical Illness Outcomes Study (ACIOS) Investigators, Gillian J Bedwell, Romy Parker, Victoria J Madden, Juan Scribante, Muhammed Elhadi, Adesoji O Ademuyiwa, Babatunde Osinaike, Christian Owoo, Daniel Sottie, Karima Khalid, Adam Hewitt-Smith, Arthur Kwizera, Fitsum Kifle Belachew, Degsew Dersso Mengistu, Yared Boru Firissa, Tirunesh Busha Gemechu, Gaudencia Dausab, Unotjari Kauta, Kaveto Sikuvi, Nahla Kechiche, Kelan Bertille Ki, Martin Mukenga, Dolly Munlemvo, Mustapha Bittaye, Abubacarr Jagne, Mohamed Abdinor Omar, Hassan Ali Daoud, Mohamed Faisal, Mahmoud Elfiky, Mpho Seleke, Tarig Fadalla, Alshaima Koko, Alemayehu G Bedada, Gilles Niengo Outsouta, Marie Elombila, Ahmed Rhassane El Adib, Meryem Essafti, Dino Lopes, Atilio Morais, Pisirai Ndarukwa, Newten Handireketi, Fred Bulamba, Busisiwe Mrara, Hyla-Louise Kluyts, Marian Kinnes, Abdullahi Said Hashi, Abigail Kusi Amponsah, Aderonke Omobonike Akinpelu, Andrew Amata, Aunel Mallier Peter, Elizabeth Nwasor, Jeremie Kalambay, Hellen N Kariuki, Nana Fening, Nomaqhawe Moyo, Patrick Mukuna, Sean Chetty, Sudarshanie Bechan, Tania Pretorius, Bamidele Victor Owoyele, Luyanduthando Mqadi, Hanél Duvenage, Gwendoline Arendse, Luke Hannan, Landon Myer, Carl Otto Schell, Tim Baker, Rupert M Pearse, Bruce M Biccard

**Author notes:** **Corresponding author**, Postal address: D23.0 Department of Anaesthesia and Perioperative Medicine, Groote Schuur Hospital, Anzio Rd, Observatory, 7925, South Africa. **Contributors** All members of the writing committee were involved in the design and conduct of the study. Data collection was undertaken by ACIOS local investigators. Data analysis was performed by GJB, GA, and BB. The first draft of the paper was prepared by GJB, RP, BB, and VJM before critical review by all members of the writing committee.

## Abstract

Acute pain in hospitalised patients is common, yet African data are scarce. We conducted a prospective point-prevalence study of hospitalised adult patients across Africa to determine acute pain prevalence and severity, and investigate associations of pain with critical illness and 7-day mortality. On a single day, investigators assessed patients’ worst pain in the preceding 24 hours using a 0–10 visual-numerical scale and recorded vital signs. Critical illness was defined as ≥1 vital sign out of range, in line with an international consensus definition. In-hospital mortality was assessed at day 7. Data are presented as median [IQR], n (%), and odds ratios with 95% confidence intervals. Between September and December 2023, 19438 patients from 180 hospitals in 22 African nations were included (age 40 [29; 59] years; 10874 (56%) female). Pain prevalence [95%CI] was 67.9% [67.9 – 68.5], with 2795 (14.4%) patients reporting severe pain (≥7/10). Pain was more common among patients admitted for emergency or trauma care, and in surgical and high-care wards. Pain severity did not differ meaningfully between sexes. There was no evidence of an association between pain severity and critical illness; however, increasing pain severity was associated with a higher likelihood of 7-day mortality. These findings are comparable to data from high-income countries despite substantial differences in healthcare resources, suggesting that acute pain remains a persistent and under-addressed problem globally. This study provides the first large-scale evidence of acute pain burden in African hospitals and lays a foundation for future research aimed at comprehensively characterising in-hospital pain.

## 1. Introduction

Acute pain is common among hospitalised patients, yet data on its prevalence and severity in African settings remains scarce. International data reflect considerable variability in pain prevalence among hospitalised patients, ranging from 38–84%, while between 9% and 36% of patients report severe pain (defined as pain of ≥7 out of 10) [23]. However, all these data come from high-income countries and thus shed little light on acute pain prevalence among patients hospitalised in African contexts, where all countries are currently classified as low– or middle-income.

Our own systematic search identified only two studies of pain prevalence among hospitalised patients outside high-income countries: one from Colombia (an upper-middle-income country) [19], and one from Morocco (an African, lower-middle-income country) [25] (Supplementary Material S1). In the Moroccan study, 41.6% of hospitalised patients reported pain, with a mean ±SD pain severity of 4.7 ± 2.01 on a 0 to 10 scale. This shortage of research limits our understanding of the burden of acute pain in African hospitals.

Understanding the burden of acute pain in African hospitals is important because uncontrolled acute pain is linked to worse health outcomes and a higher burden on healthcare systems. In high-income settings, poorly controlled acute pain is associated with adverse clinical outcomes including impaired immune function, hypercoagulability, poor glycaemic control, delayed mobilisation, prolonged hospitalisation, and an increased risk of developing persistent pain [15, 16, 18, 41, 47]. Severe acute pain also strains healthcare systems through longer hospital stays and increased readmission rates, thus raising healthcare costs [22, 27]. Although these associations do not establish causality, they do suggest that acute pain represents a potentially modifiable factor that may improve both patient outcomes and health system capacity.

In addition, several African countries have endorsed the assertion that access to pain assessment and relief is a fundamental human right, as articulated in the International Association for the Study of Pain Declaration of Montréal [29, 30] – and understanding the burden of acute pain is a crucial first step towards protecting this right. As such, there is a pressing need to understand the burden of acute pain across African hospitals.

This study aimed to address this priority by reporting the prevalence and severity of acute pain among hospitalised adults across multiple African hospitals. Specifically, we aimed to determine the (i) prevalence of any pain (≥1/10), (ii) severity of pain, and (iii) prevalence of severe pain (≥7/10) among hospitalised adults across Africa. Our secondary aims were to investigate the associations of pain rating (0 – 10) with (i) critical illness and (ii) in-hospital seven-day mortality, and (iii) to describe in-hospital mortality over time, for patients with and without severe pain. We hypothesised that greater pain severity would be associated with a higher likelihood of critical illness and in-hospital seven-day mortality.

## 2. Methods

### 2.1 Study design

This is a substudy of the African Critical Illness Outcomes Study (ACIOS), an international, prospective, point-prevalence study of critical illness amongst adult hospitalised patients across Africa [6]. The ACIOS was open to all African countries or territories, and hospitals were recruited through the African Perioperative Research Group (APORG, http://www.asos.org.za/index.php/aporg) and the Essential Emergency and Critical Care Network (www.eeccnetwork.org). The primary ethics approval was provided by the University of Cape Town’s Human Research Ethics Committee (REF 260/2023). Ethics approval processes varied between countries (Supplementary Material S2). All national ethics committees approved a waiver of informed consent given that the dataset included only variables documented as part of routine clinical care. ‘Broadcasting’ signage was used to inform patients and families that the hospital was participating in the study (Supplementary Material S3). The protocol and original statistical analysis plan (Supplementary Material S4) were prospectively registered on ClinicalTrials.gov (NCT06051526). An amended statistical analysis plan (Supplementary Material S5) was uploaded on ClinicalTrials.gov on 31 October 2025. The results of this study are reported in accordance with the STROBE statement [48].

### 2.2 Setting and patients

#### 2.2.1 Hospital site eligibility

Any hospital admitting acutely unwell patients was eligible to participate, regardless of hospital funding source. Hospitals admitting patients exclusively for elective surgery, psychiatric illness, or rehabilitation were ineligible.

#### 2.2.2 Patient eligibility

All adult (≥18 years old) patients receiving inpatient care in any department or ward of a participating hospital on the day of data collection were eligible for inclusion. This included patients admitted to maternity and emergency departments, provided they had been formally admitted for inpatient care. Patients with a primary psychiatric diagnosis were excluded, as were individuals not admitted for inpatient care (i.e. outpatients, and emergency department attendees managed without hospital admission). All eligible patients were approached for participation and included unless they opted out of participation.

### 2.3 Data collection

The data collection period ran from 6 September to 27 December 2023. Local investigators at each hospital selected a single day within this period to collect data. Participating hospitals’ characteristics were collected, including hospital level (Level 1: district, Level 2: regional, or Level 3: university, central or national), total number of beds, number of beds in the high care unit(s) and in the intensive care unit(s), and the size of the population served by the hospital (Supplementary Material S6). Local investigators recorded data on standardised paper case record forms from patients’ notes and by direct patient observation at one point in time i.e. when the local investigator was at the patient’s bedside (Supplementary Material S7 and S8). Patients were managed by local clinical staff in line with local hospital standards and protocols. Clinical staff were immediately notified if investigators identified a patient needing urgent care. Pain severity ratings and vital signs (respiratory rate, oxygen saturation, blood pressure, heart rate, consciousness level) were measured by local investigators, unless impossible, in which case these data were extracted from the documented clinical observations. We did not record whether pain severity ratings were obtained via direct patient self-report or extracted from clinical documentation, in order to minimise data collection burden across participating sites in this large, multi-country study. All included patients were followed up at day seven to ascertain in-hospital seven-day mortality. Data were pseudo-anonymised using a unique numeric code before entry into an internet-based electronic case record form. Identifiable patient data were stored in a locked office at each hospital.

### 2.4 Outcome measures

#### 2.4.1 Primary outcomes: Pain prevalence and severity

Pain severity was self-reported using a vertical visual-numerical scale ranging from 0 (no pain) to 10 (the worst pain you can imagine), reflecting the worst pain experienced in the preceding 24 hours. For patients with reduced level of consciousness, pain ratings were obtained from the last clinical pain documentation where available; if not available, pain ratings were recorded as missing.

#### 2.4.2 Secondary outcome: Critical illness

In line with an international consensus definition [32], patients were classified as critically ill if one or more vital sign was out of range [4, 5, 11]. This was operationalised as: respiratory rate <8 or >30 breaths per minute, oxygen saturation <90% (pulse oximetry), systolic blood pressure <90 mmHg, heart rate <40 or >130 beats per minute, or reduced level of consciousness (responsive to pain or unresponsive on the alert, voice, pain, unresponsive (AVPU) scale). For women in active labour, vital signs were assessed between contractions. In a small number of cases where it was inappropriate to measure a particular vital sign, e.g. blood pressure in a patient on an end-of-life care pathway, the most recent recorded value in the patient’s records was used.

#### 2.4.3 Secondary outcome: In-hospital seven-day mortality

In-hospital seven-day mortality was assessed seven days after the initial pain and critical illness assessments. Included patients were classified as “alive” if they had been discharged alive or remained in the hospital receiving treatment at day seven, or “deceased” if they had died at any time in hospital between the initial and the follow-up assessments.

### 2.5 Data quality assurance

Data quality was protected in two steps. Electronic case record forms incorporated validation rules to prevent implausible values at the point of entry. Following data submission, each site received a summary of included patients and aggregated outcomes to verify completeness and accuracy. Any potentially implausible or inconsistent values were queried and resolved in consultation with local investigators.

### 2.6 Sample size

There was no prespecified sample size, as we aimed to include as many hospitals as possible and enrol all eligible patients at each hospital site. However, a post hoc sample size calculation confirmed that the study had adequate statistical power to include the eight independent predictors of death at seven days [6] and ‘severe pain’ predictor variable in the survival status regression model, as the number of candidate variables did not exceed 10 events per variable [36]. The primary ACIOS study [6] enrolled 19 872 patients; the current analysis included the 19 438 hospitalised patients with complete pain severity data (97.82% of the ACIOS sample).

### 2.7 Statistical analysis

Data analysis followed a prespecified statistical analysis plan, which was amended prior to analysis and posted on ClinicalTrials.gov (NCT06051526). Data were prepared using the following packages: haven [53], dplyr [51], magrittr [34], tidyverse [50], tidyr [55], data.table [8], janitor [20], lubridate [24], purrr [52], and scales [54]. Data were analysed using the following packages: lme4 [9], coxme [45], survival [46], ggsurvfit [43], survminer [31], ROCR [42], Matrix [10], binom [17], broom [37], broom.mixed [13], and ggplot2 [49]. Demographic characteristics are presented as n (%).

#### 2.7.1 Primary aims: Pain prevalence and severity

We calculated the prevalence [95% confidence interval] of any pain (≥1/10) and of severe pain (≥7/10) for all patients, and for predefined clinical subgroups (age category, pregnancy, comorbidities, urgency of admission, admission diagnosis, ward type, ward level, critical illness status, and in-hospital seven-day mortality status). Pain prevalence was also calculated for each hospital to demonstrate the range in prevalence across sites. For subgroup comparisons, we prioritised absolute effect sizes rather than p values. Specifically, between-group differences were described using absolute risk differences, expressed as percentage-point differences in prevalence relative to a clinically relevant reference group. This approach was chosen because the large sample size would make even small differences likely to reach statistical significance, whereas absolute risk differences provide a more clinically interpretable indication of the magnitude of between-group differences. Pain severity was primarily analysed as a continuous variable (0 – 10). Pain ratings were not normally distributed and, as expected for a bounded 0 – 10 scale, showed clustering around some values (Supplementary material S9). Pain severity is therefore reported as the median [IQR] to best represent the current data, but also as mean ±SD to facilitate comparison with existing pain literature where mean ±SD is commonly reported. We also report pain severity separately for males vs females and compare distribution of pain ratings between sexes using the Wilcoxon rank-sum test.

For descriptive and clinically meaningful analyses, pain was additionally categorised into severity groups (0: no pain; 1 – 3: mild pain; 4 – 6: moderate pain; ≥7: severe pain [2]). Although categorisation reduces measurement resolution, these thresholds are widely used and support clinical interpretation by identifying patients experiencing severe pain, a level associated with functional impairment and adverse outcomes [1, 7, 16, 41]. Reporting categorical pain severity also facilitates comparison with similar published studies and provides additional insight into the distribution of pain across clinically relevant strata, particularly at the extremes, which may not be fully captured by summary statistics alone.

#### 2.7.2 Secondary aims: Relationship between pain rating (0 – 10) and critical illness and in-hospital seven-day mortality

We conducted unadjusted and multivariable logistic regression analyses to determine the relationship between pain rating (0 – 10) and (i) critical illness and (ii) in-hospital seven-day mortality. Pain rating (0 – 10) was modelled as the continuous independent variable. Critical illness and in-hospital seven-day mortality were each included as binary dependent variables. The logistic regressions were three-level models with patients nested within hospital, and hospital nested within country. Random effects were assumed to be normally distributed, and the default link function was the logit function. Model diagnostics and fit were assessed using simulated residuals generated by the DHARMa package in R [26]. Associations are presented as odds ratios (OR) with 95% confidence intervals and p-values, where p<0.05 was the threshold for statistical significance.

For the multivariable logistic regression analyses, we pragmatically identified candidate covariates a priori, based on their clinical importance for in-hospital critical illness and death, and the availability of data in routine hospital records. These candidate covariates were: age, sex, urgency of admission (elective vs emergency/acute), admission diagnosis (non-communicable disease, trauma, infection, or maternal health), pregnancy, and known comorbidities (hypertension, diabetes, cancer, chronic obstructive pulmonary disease or asthma, heart disease, HIV/AIDS, tuberculosis, or other). All these covariates were included as fixed effects in the multivariable regression analyses investigating the relationship between pain and critical illness.

For the multivariable logistic regression analyses investigating the relationship between pain and in-hospital seven-day mortality, covariate selection was facilitated by the results of the primary ACIOS study [6], which had identified eight independent predictors of seven-day mortality: age, cancer, HIV infection, emergency admission, admission diagnosis of infection, non-communicable disease, or trauma, and critical illness. Accordingly, these eight covariates were included as fixed effects in the multivariable regression analyses investigating the relationship between pain and in-hospital seven-day mortality.

A Kaplan-Meier survival plot compared in-hospital mortality over seven days for patients with and without severe pain. For the Kaplan-Meier plot, patients were censored if they were discharged before day seven or if they were still alive and in hospital at day seven

#### 2.7.3 Sensitivity analyses

To assess for disproportionate influence over the results from a small number of high-contributing countries (particularly given the heterogeneity in healthcare systems, case mix, and pain management practices across settings), we conducted a sensitivity analyses for the secondary aims by excluding data from countries that had contributed more than 10% of the total sample (threshold defined a priori).

#### 2.7.4 Exploratory analyses

We conducted three exploratory analyses. In the first, we examined the role of sex and age in the odds of having severe pain (severe pain: ≥7; non-severe pain: <7), given that both sex and age can shape behaviours such as pain reporting and care delivery [28, 39]. We fitted a univariate logistic regression model with severe pain status as the binary dependent variable and sex as the binary independent variable. Age was included as a continuous variable and was standardised (centred on the mean and scaled by the standard deviation) to aid interpretation and model stability. We also conducted a univariate regression in which we included a sex-by-age interaction term to assess whether the association between sex and severe pain varied across age.

Given that ‘severe pain’ has been identified as a clinically meaningful threshold associated with adverse outcomes [15, 21, 47], in the second and third exploratory analyses, we examined whether severe pain status (severe pain: ≥7; non-severe pain: <7) was associated with critical illness status (2^nd^ exploratory analysis) and in-hospital mortality seven-day mortality (3^rd^ exploratory analysis). Severe pain (≤7 /10) was the binary independent variable. Critical illness and in-hospital mortality seven-day mortality were binary dependent variables. The logistic regressions were three-level models with patients nested within hospital, and hospital nested within country. Random effects were assumed to be normally distributed, and the default link function was the logit function. Model diagnostics and fit were assessed using simulated residuals generated by the DHARMa package in R [26].

#### 2.7.5 Missing data

Patients with missing data (excluding pain severity data) were included without imputation and any missing data were reported descriptively, as data missingness was <1%.

## 3. Results

### 3.1 Study sites and participants

The primary ACIOS study enrolled 19 872 patients from 180 hospitals across 22 African countries and territories (Botswana, Burkina Faso, Congo, Democratic Republic of Congo, Egypt, Ethiopia, The Gambia, Ghana, Lesotho, Libya, Morocco, Mozambique, Namibia, Nigeria, Somalia, Somaliland, South Africa, Sudan, Tanzania, Tunisia, Uganda, Zimbabwe) (Fig 1 and Supplementary Material S10), of whom 19 438 (97.82%) had pain severity data and are included in this analysis.

**Figure 1:**
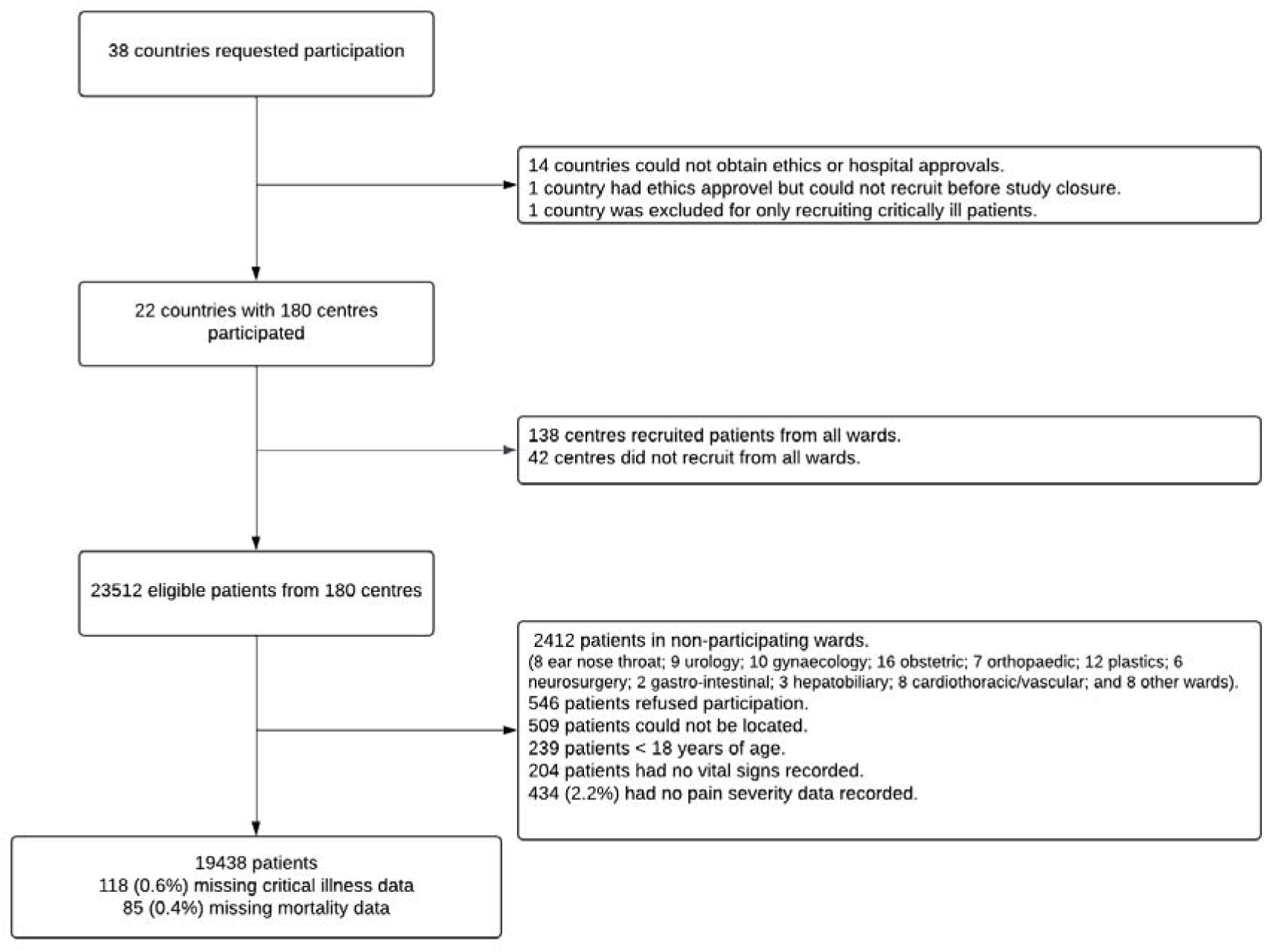
Patient recruitment.

The characteristics of the 19 438 included patients are shown in Table 1. The median [IQR] age was 40 [29; 59] years old, and 56% (10 874/19 429) of the patients were female. One quarter (4 869/19 438 (25%)) of the sample was known to have hypertension. Most (14 740/19 350 (76.2%)) had been admitted for emergency/acute care. Almost half (9 102/19 375 (47.0%)) had an admission diagnosis of non-communicable disease(s). Most patients were receiving care in either a medical (7 179/19 432 (36.9%)) or surgical ward (7 341/19 432 (37.8%)).

**Table 1:**
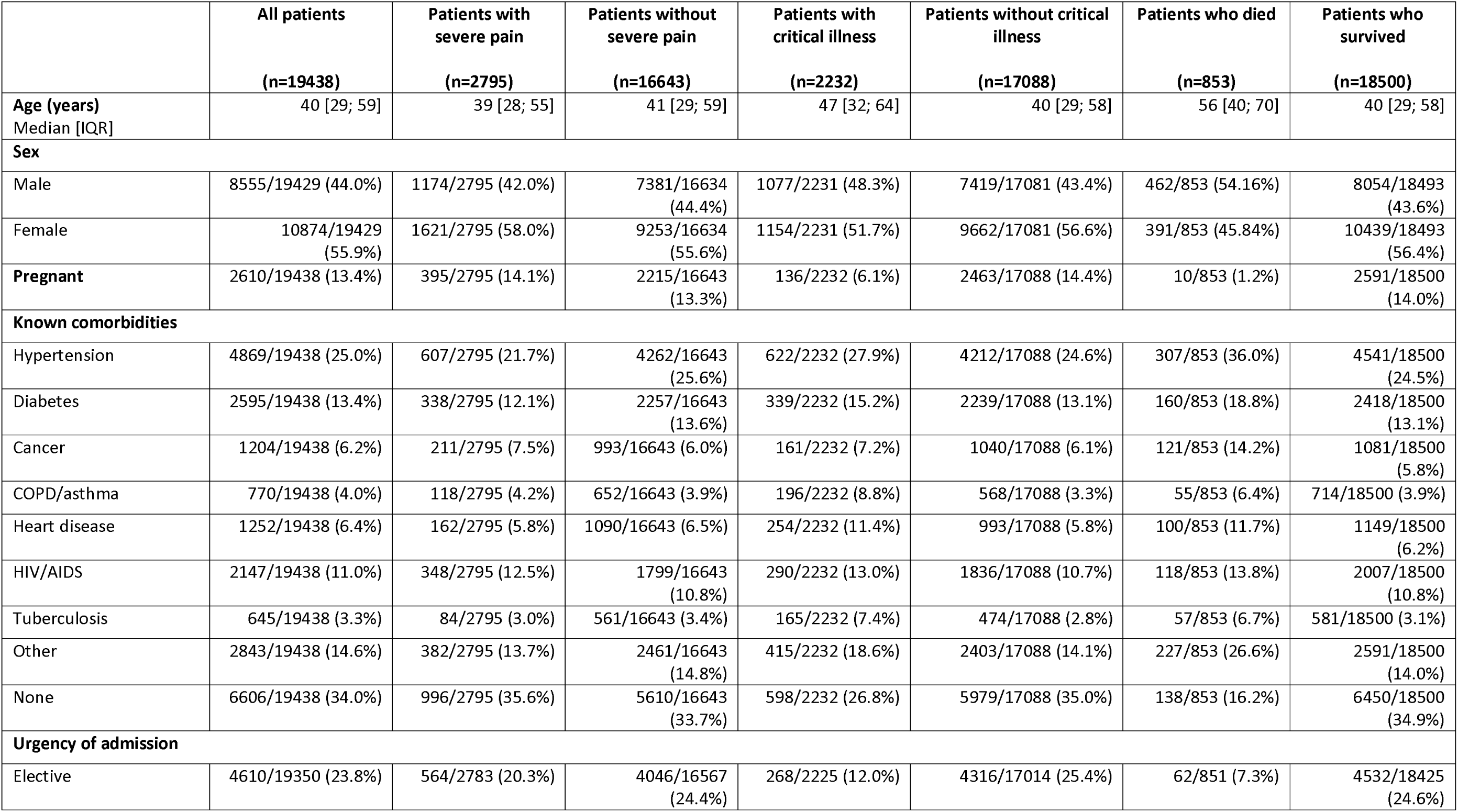
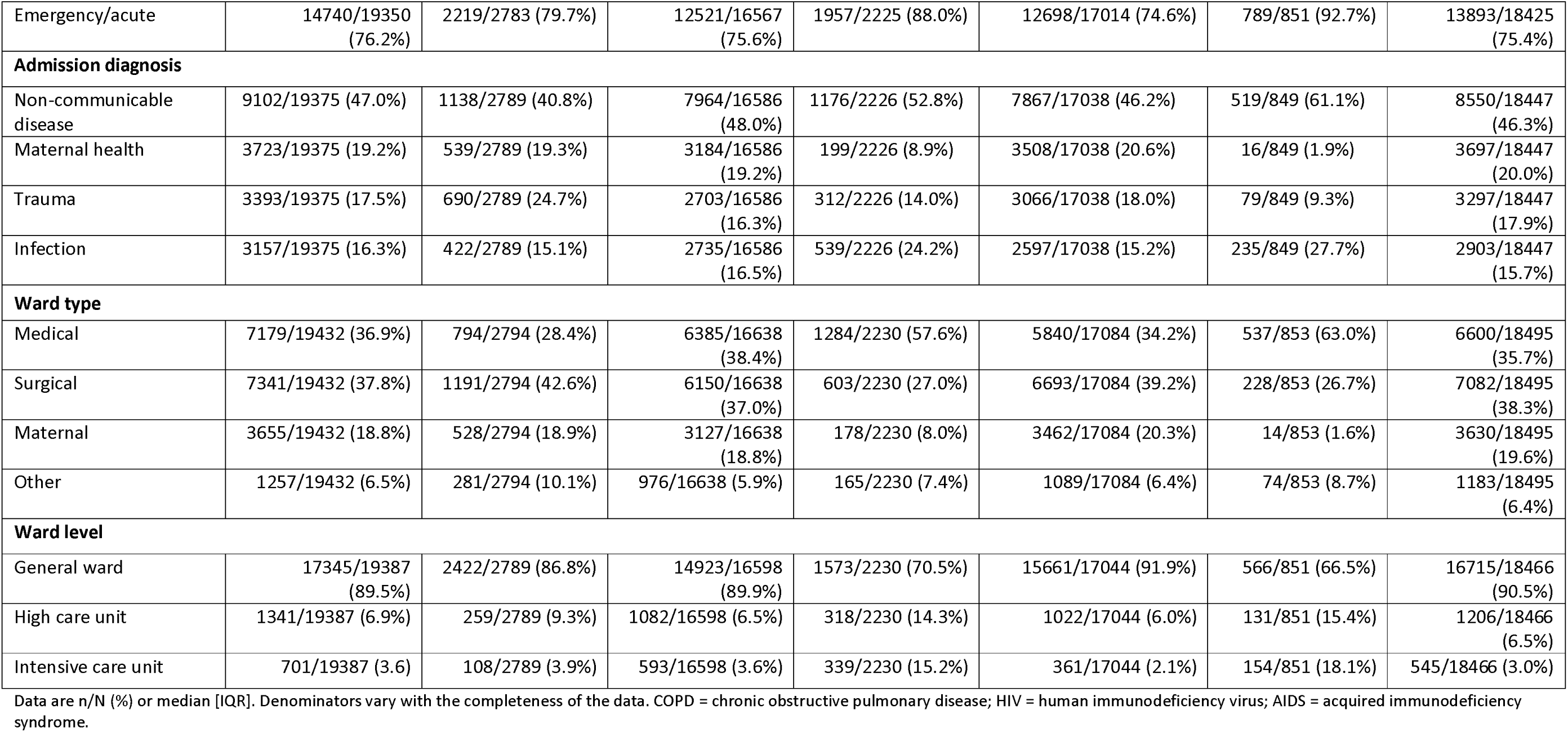
Descriptive characteristics.

|  | All patients<br>(n=19438) | Patients with<br>severe pain<br>(n=2795) | Patients without<br>severe pain<br>(n=16643) | Patients with<br>critical illness<br>(n=2232) | Patients without critical<br>illness<br>(n=17088) | Patients who died<br>(n=853) | Patients who<br>survived<br>(n=18500) |
| --- | --- | --- | --- | --- | --- | --- | --- |
| Age (years)<br>Median [IQR] | 40 [29; 59] | 39 [28; 55] | 41 [29; 59] | 47 [32; 64] | 40 [29; 58] | 56 [40; 70] | 40 [29; 58] |
| **Sex** |  |  |  |  |  |  |  |
| Male | 8555/19429 (44.0%) | 1174/2795 (42.0%) | 7381/16634<br>(44.4%) | 1077/2231 (48.3%) | 7419/17081 (43.4%) | 462/853 (54.16%) | 8054/18493<br>(43.6%) |
| Female | 10874/19429<br>(55.9%) | 1621/2795 (58.0%) | 9253/16634<br>(55.6%) | 1154/2231 (51.7%) | 9662/17081 (56.6%) | 391/853 (45.84%) | 10439/18493<br>(56.4%) |
| Pregnant | 2610/19438 (13.4%) | 395/2795 (14.1%) | 2215/16643<br>(13.3%) | 136/2232 (6.1%) | 2463/17088 (14.4%) | 10/853 (1.2%) | 2591/18500<br>(14.0%) |
| **Known comorbidities** |  |  |  |  |  |  |  |
| Hypertension | 4869/19438 (25.0%) | 607/2795 (21.7%) | 4262/16643<br>(25.6%) | 622/2232 (27.9%) | 4212/17088 (24.6%) | 307/853 (36.0%) | 4541/18500<br>(24.5%) |
| Diabetes | 2595/19438 (13.4%) | 338/2795 (12.1%) | 2257/16643<br>(13.6%) | 339/2232 (15.2%) | 2239/17088 (13.1%) | 160/853 (18.8%) | 2418/18500<br>(13.1%) |
| Cancer | 1204/19438 (6.2%) | 211/2795 (7.5%) | 993/16643 (6.0%) | 161/2232 (7.2%) | 1040/17088 (6.1%) | 121/853 (14.2%) | 1081/18500<br>(5.8%) |
| COPD/asthma | 770/19438 (4.0%) | 118/2795 (4.2%) | 652/16643 (3.9%) | 196/2232 (8.8%) | 568/17088 (3.3%) | 55/853 (6.4%) | 714/18500 (3.9%) |
| Heart disease | 1252/19438 (6.4%) | 162/2795 (5.8%) | 1090/16643 (6.5%) | 254/2232 (11.4%) | 993/17088 (5.8%) | 100/853 (11.7%) | 1149/18500<br>(6.2%) |
| HIV/AIDS | 2147/19438 (11.0%) | 348/2795 (12.5%) | 1799/16643<br>(10.8%) | 290/2232 (13.0%) | 1836/17088 (10.7%) | 118/853 (13.8%) | 2007/18500<br>(10.8%) |
| Tuberculosis | 645/19438 (3.3%) | 84/2795 (3.0%) | 561/16643 (3.4%) | 165/2232 (7.4%) | 474/17088 (2.8%) | 57/853 (6.7%) | 581/18500 (3.1%) |
| Other | 2843/19438 (14.6%) | 382/2795 (13.7%) | 2461/16643<br>(14.8%) | 415/2232 (18.6%) | 2403/17088 (14.1%) | 227/853 (26.6%) | 2591/18500<br>(14.0%) |
| None | 6606/19438 (34.0%) | 996/2795 (35.6%) | 5610/16643<br>(33.7%) | 598/2232 (26.8%) | 5979/17088 (35.0%) | 138/853 (16.2%) | 6450/18500<br>(34.9%) |
| **Urgency of admission** |  |  |  |  |  |  |  |
| Elective | 4610/19350 (23.8%) | 564/2783 (20.3%) | 4046/16567<br>(24.4%) | 268/2225 (12.0%) | 4316/17014 (25.4%) | 62/851 (7.3%) | 4532/18425<br>(24.6%) |
| Emergency/acute | 14740/19350<br>(76.2%) | 2219/2783 (79.7%) | 12521/16567<br>(75.6%) | 1957/2225 (88.0%) | 12698/17014 (74.6%) | 789/851 (92.7%) | 13893/18425<br>(75.4%) |
| **Admission diagnosis** |  |  |  |  |  |  |  |
| Non-communicable disease | 9102/19375 (47.0%) | 1138/2789 (40.8%) | 7964/16586<br>(48.0%) | 1176/2226 (52.8%) | 7867/17038 (46.2%) | 519/849 (61.1%) | 8550/18447<br>(46.3%) |
| Maternal health | 3723/19375 (19.2%) | 539/2789 (19.3%) | 3184/16586<br>(19.2%) | 199/2226 (8.9%) | 3508/17038 (20.6%) | 16/849 (1.9%) | 3697/18447<br>(20.0%) |
| Trauma | 3393/19375 (17.5%) | 690/2789 (24.7%) | 2703/16586<br>(16.3%) | 312/2226 (14.0%) | 3066/17038 (18.0%) | 79/849 (9.3%) | 3297/18447<br>(17.9%) |
| Infection | 3157/19375 (16.3%) | 422/2789 (15.1%) | 2735/16586<br>(16.5%) | 539/2226 (24.2%) | 2597/17038 (15.2%) | 235/849 (27.7%) | 2903/18447<br>(15.7%) |
| **Ward type** |  |  |  |  |  |  |  |
| Medical | 7179/19432 (36.9%) | 794/2794 (28.4%) | 6385/16638<br>(38.4%) | 1284/2230 (57.6%) | 5840/17084 (34.2%) | 537/853 (63.0%) | 6600/18495<br>(35.7%) |
| Surgical | 7341/19432 (37.8%) | 1191/2794 (42.6%) | 6150/16638<br>(37.0%) | 603/2230 (27.0%) | 6693/17084 (39.2%) | 228/853 (26.7%) | 7082/18495<br>(38.3%) |
| Maternal | 3655/19432 (18.8%) | 528/2794 (18.9%) | 3127/16638<br>(18.8%) | 178/2230 (8.0%) | 3462/17084 (20.3%) | 14/853 (1.6%) | 3630/18495<br>(19.6%) |
| Other | 1257/19432 (6.5%) | 281/2794 (10.1%) | 976/16638 (5.9%) | 165/2230 (7.4%) | 1089/17084 (6.4%) | 74/853 (8.7%) | 1183/18495<br>(6.4%) |
| **Ward level** |  |  |  |  |  |  |  |
| General ward | 17345/19387<br>(89.5%) | 2422/2789 (86.8%) | 14923/16598<br>(89.9%) | 1573/2230 (70.5%) | 15661/17044 (91.9%) | 566/851 (66.5%) | 16715/18466<br>(90.5%) |
| High care unit | 1341/19387 (6.9%) | 259/2789 (9.3%) | 1082/16598 (6.5%) | 318/2230 (14.3%) | 1022/17044 (6.0%) | 131/851 (15.4%) | 1206/18466<br>(6.5%) |
| Intensive care unit | 701/19387 (3.6) | 108/2789 (3.9%) | 593/16598 (3.6%) | 339/2230 (15.2%) | 361/17044 (2.1%) | 154/851 (18.1%) | 545/18466 (3.0%) |

Data are n/N (%) or median [IQR]. Denominators vary with the completeness of the data. COPD = chronic obstructive pulmonary disease; HIV = human immunodeficiency virus; AIDS = acquired immunodeficiency syndrome.

### 3.2 Missing data

No patient-level variable had a missingness rate over 0.6% (Supplementary Material S11).

### 3.3 Primary aims: Pain prevalence and severity

#### 3.3.1 Prevalence of any pain (i.e. ≥1 out of 10)

Of the 19 438 patients, the average prevalence [95%CI] of any pain (≥1/10) was 67.9% [67.2 – 68.5] (Table 2). Hospital-level prevalences of any pain ranged from 0% to 100%. Pain was more common among patients admitted for emergency/acute care than those admitted for elective care (68.9% vs 64.4; absolute difference +4.6 percentage points). Across the four categories of admission, which were admissions for non-communicable disease, maternal health, trauma, and infection, pain prevalence was highest among patients admitted for trauma (84.1%), exceeding the pain prevalence among patients admitted for non-communicable disease by 21.5 percentage points. Pain prevalence was also higher in surgical wards (77.3%) than in medical (58.6%; absolute difference between surgical and medical wards: +18.7 percentage points) or maternal wards (65.7%), and higher in high-care units (75.2%) than in general wards (67.5%; absolute difference between high-care and general wards: +7.8 percentage points) or intensive care units (62.3%). Details can be found in Table 2.

**Table 2:** Summary of the prevalence of any pain (≥1/10 and of severe pain (≥7/10).

| Category | n | Prevalence of any pain (≥1/10) with 95% CI | Prevalence of severe pain (≥7/10) with 95% CI |
| --- | --- | --- | --- |
| All patients | 19438 | 67.9% [67.2–68.5] | 14.4% [13.9–14.9] |
| **Age categories (years)** |  |  |  |
| 18 – 30 | 5510 | 68.4% [67.1–69.6] | 15.6% [14.7–16.6] |
| 31 – 40 | 4296 | 68.6% [67.1–69.9] | 15.2% [14.2–16.3] |
| 41 – 50 | 2779 | 69.5% [67.8–71.2] | 15.0% [13.8–16.4] |
| 51 – 60 | 2507 | 67.5% [65.6–69.3] | 14.0% [12.7–15.4] |
| 61 – 70 | 2262 | 65.0% [63.0–67.0] | 12.2% [10.9–13.6] |
| 71 – 80 | 1405 | 66.1% [63.6–68.5] | 10.9% [9.4–12.6] |
| 81 – 90 | 555 | 65.8% [61.7–69.6] | 11.7% [9.3–14.7] |
| >90 | 111 | 71.2% [62.1–78.8] | 14.4% [9.1–22.1] |
| Missing data | 13 | 69.2% [42.4–87.3] | 15.4% [4.3–42.2] |
| **Urgency of admission** |  |  |  |
| Elective | 4610 | 64.4% [63.0–65.7] | 12.2% [11.3–13.2] |
| Emergency/acute | 14740 | 68.9% [68.2–69.7] | 15.1% [14.5–15.6] |
| Missing data | 88 | 70.5% [60.2–79.0] | 13.6% [8.0–22.3] |
| **Admission diagnosis** |  |  |  |
| Non-communicable disease | 9102 | 62.6% [61.6–63.6] | 12.5% [11.8–13.2] |
| Maternal health | 3723 | 65.8% [64.2–67.3] | 14.5% [13.4–15.6] |
| Trauma | 3393 | 84.1% [82.8–85.2] | 20.3% [19.0–21.7] |
| Infection | 3157 | 68.3% [66.6–69.9] | 13.4% [12.2–14.6] |
| Missing data | 63 | 60.3% [48.0–71.5] | 9.5% [4.4–19.3] |
| **Ward type** |  |  |  |
| Maternal | 3655 | 65.7% [64.2–67.3] | 14.4% [13.3–15.6] |
| Medical | 7179 | 58.6% [57.4–59.7] | 11.1% [10.4–11.8] |
| Surgical | 7341 | 77.3% [76.3–78.2] | 16.2% [15.4–17.1] |
| Other | 1257 | 71.9% [69.4–74.3] | 22.4% [20.1–24.7] |
| Missing data | 6 | 83.3% [43.6–97.0] | 16.7% [3.0–56.4] |
| **Ward level** |  |  |  |
| General ward | 17345 | 67.5% [66.8–68.2] | 14.0% [13.5–14.5] |
| High care | 1341 | 75.2% [72.9–77.5] | 19.3% [17.3–21.5] |
| Intensive care | 701 | 62.3% [58.7–65.8] | 15.4% [12.9–18.3] |

| Category | n | Prevalence of any pain ($\geq 1/10$) with 95% CI | Prevalence of severe pain ($\geq 7/10$) with 95% CI |
| --- | --- | --- | --- |
| Missing data | 51 | 74.5% [61.1–84.5] | 11.8% [5.5–23.4] |
| **Critical illness status** |  |  |  |
| Critically illness | 2232 | 64.5% [62.5–66.5] | 17.0% [15.5–18.6] |
| Not critically ill | 17088 | 68.3% [67.6–69.0] | 14.0% [13.5–14.5] |
| Missing data | 118 | 66.1% [57.2–74.0] | 20.3% [14.1–28.5] |
| **Survival status** |  |  |  |
| Dead | 853 | 66.8% [63.6–69.9] | 17.7% [15.3–20.4] |
| Alive | 18500 | 67.8% [67.1–68.5] | 14.2% [13.7–14.7] |
| Missing data | 85 | 85.9% [76.9–91.7] | 12.9% [7.4–21.7] |

#### 3.3.2 Severity of pain

Across the full sample, the median [IQR] pain severity was 3 [0–5] and the mean ± SD pain severity was 3 ± 3 (Table 3). Approximately one third (32.1%) of patients reported no pain, 28% reported mild pain (1 – 3 out of 10), 25.4% reported moderate pain (4 – 6 out of 10), and 14.4% reported severe pain (≥7 out of 10) (Fig 2). Overall, there was no evidence of a meaningful difference in pain severity ratings between males and females (Table 3).

**Figure 2:**
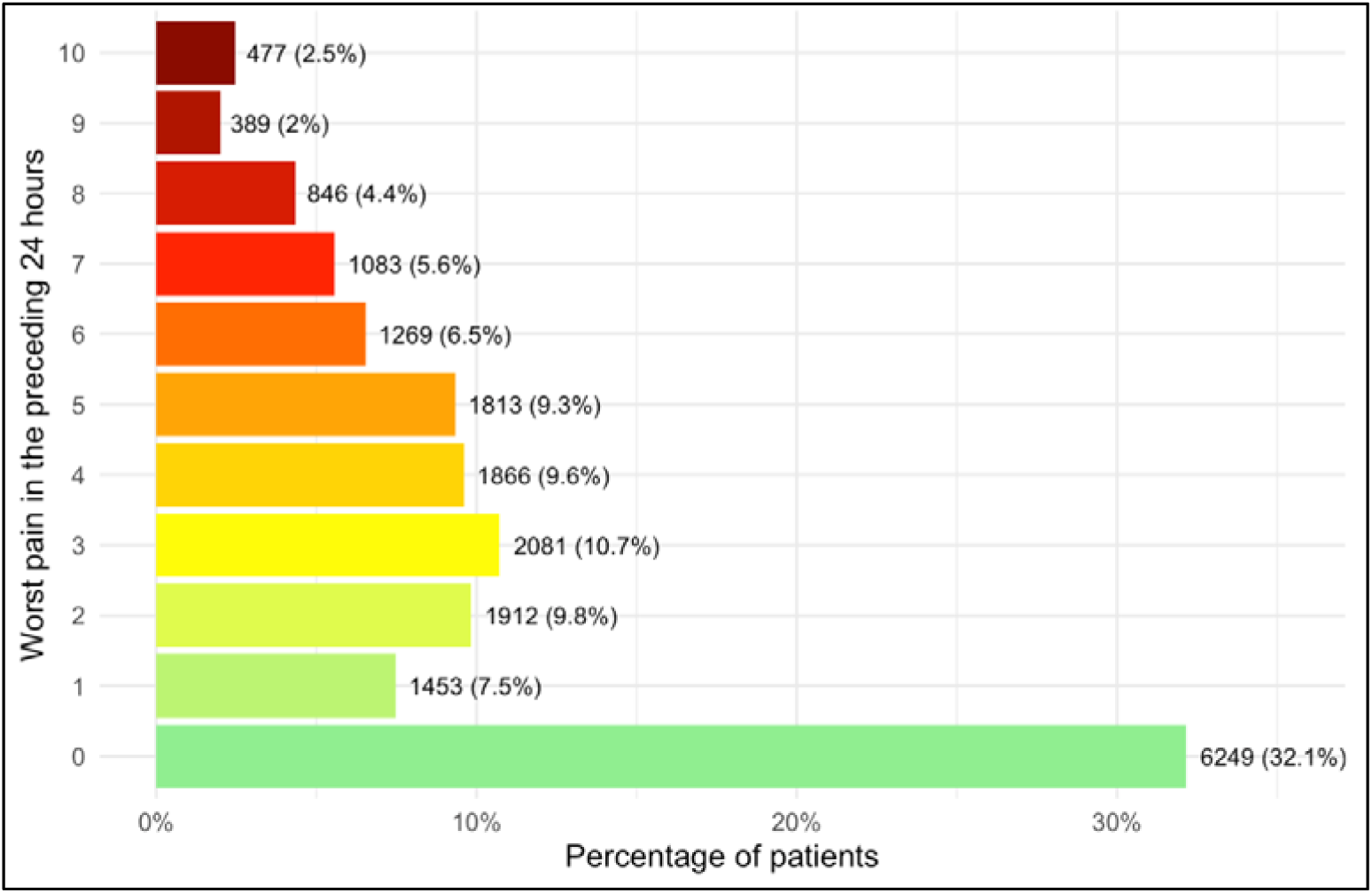
Proportion of patients (n=19438) reporting each level of self-reported pain severity.

**Table 3:**
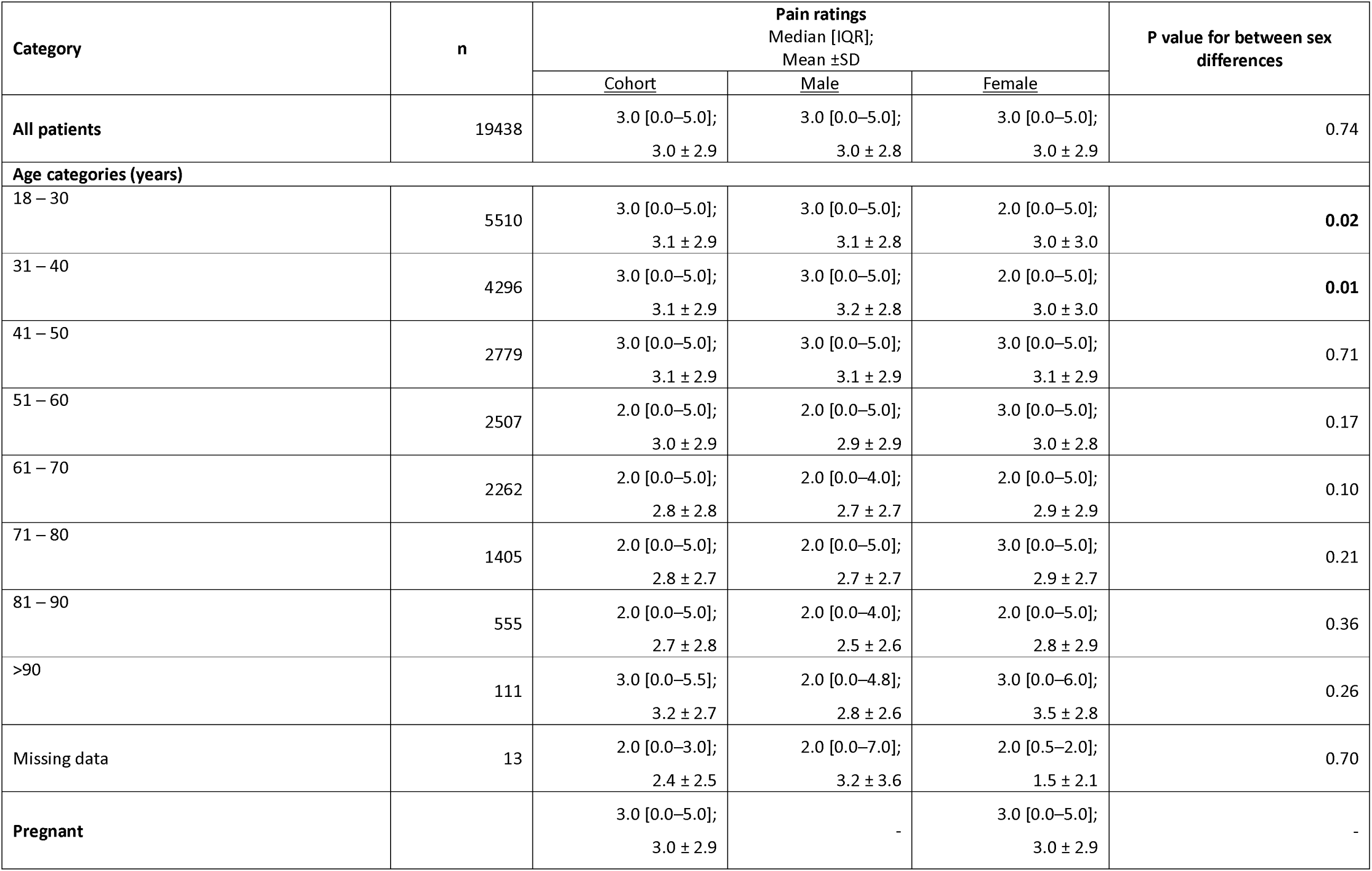
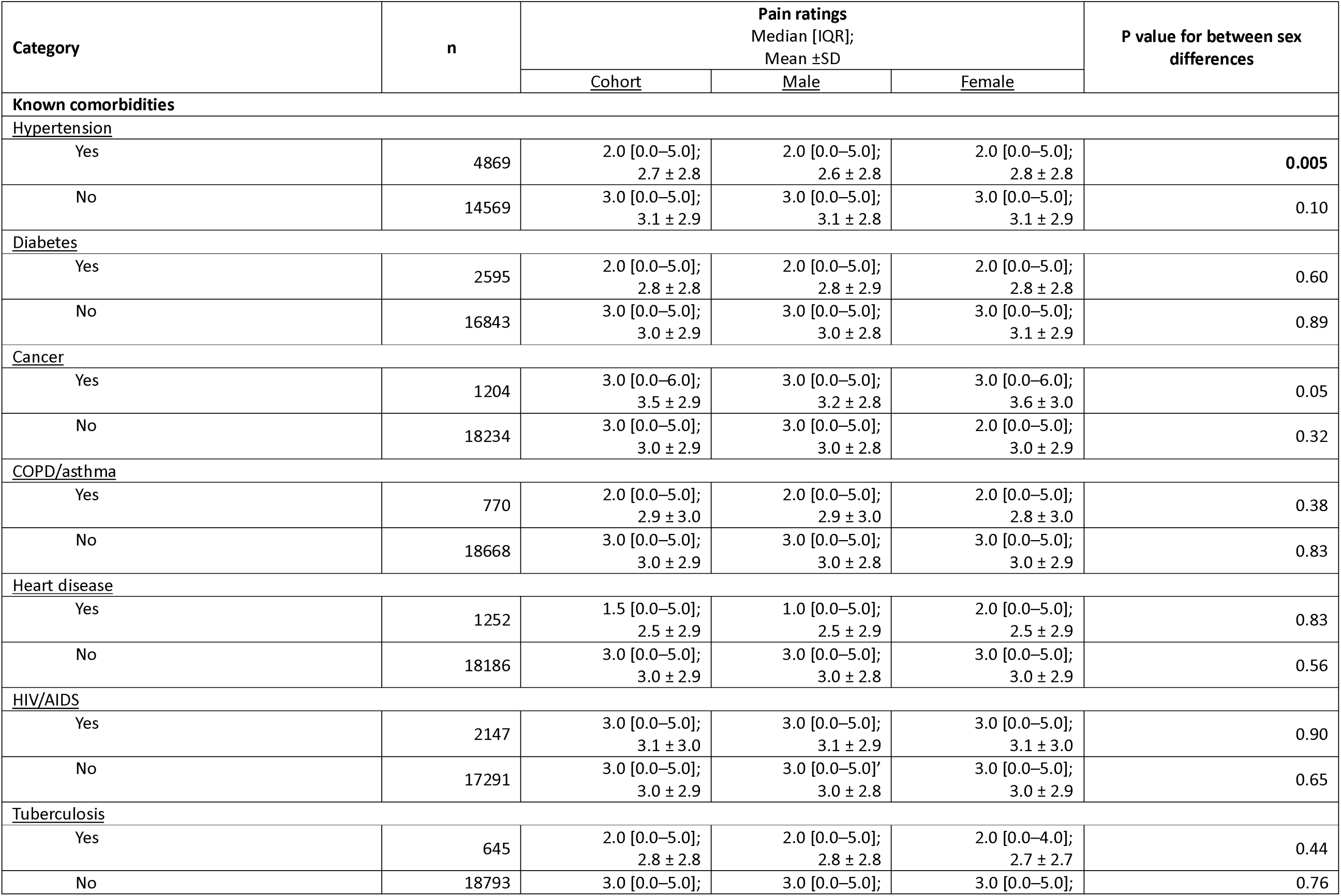
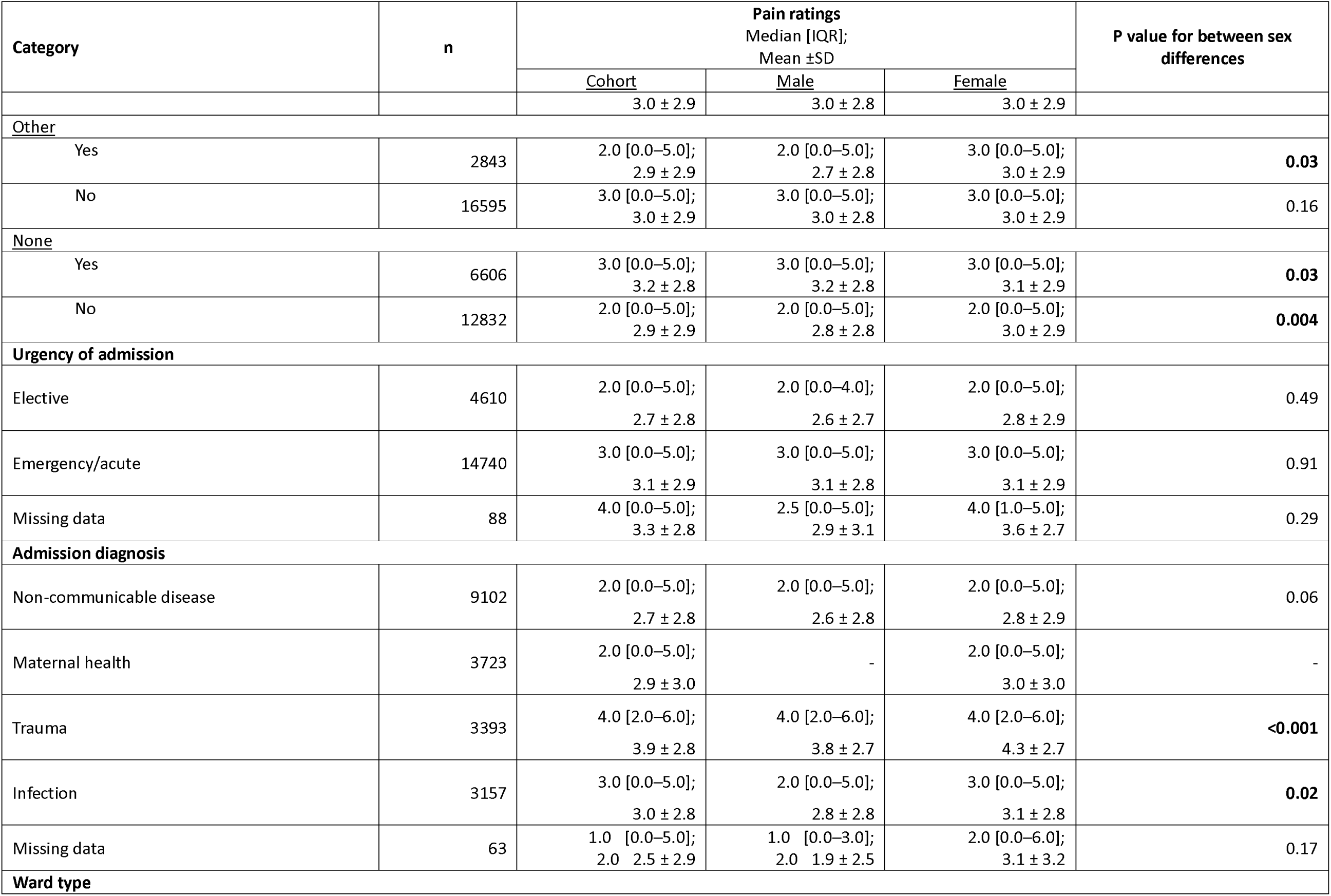
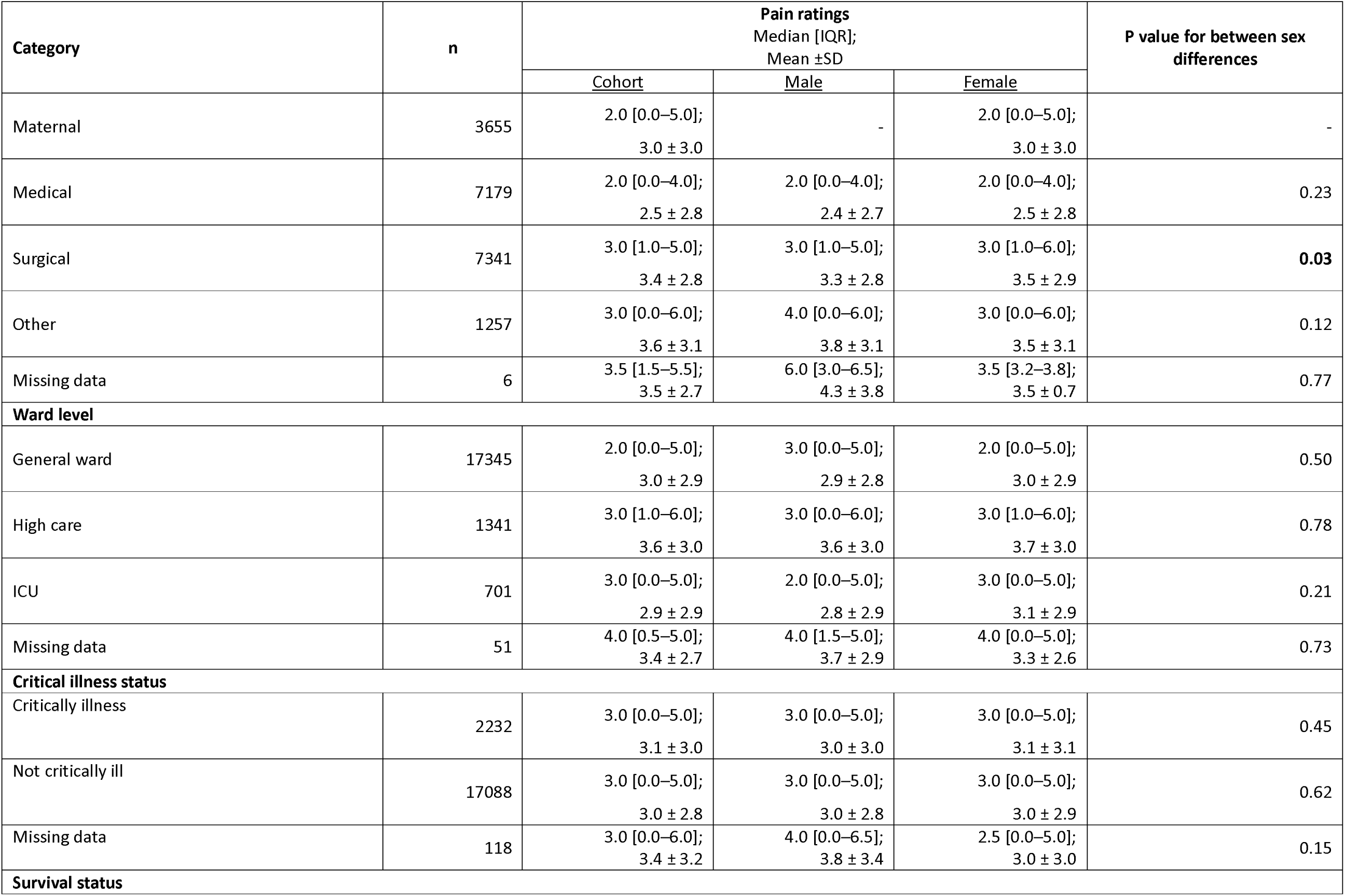
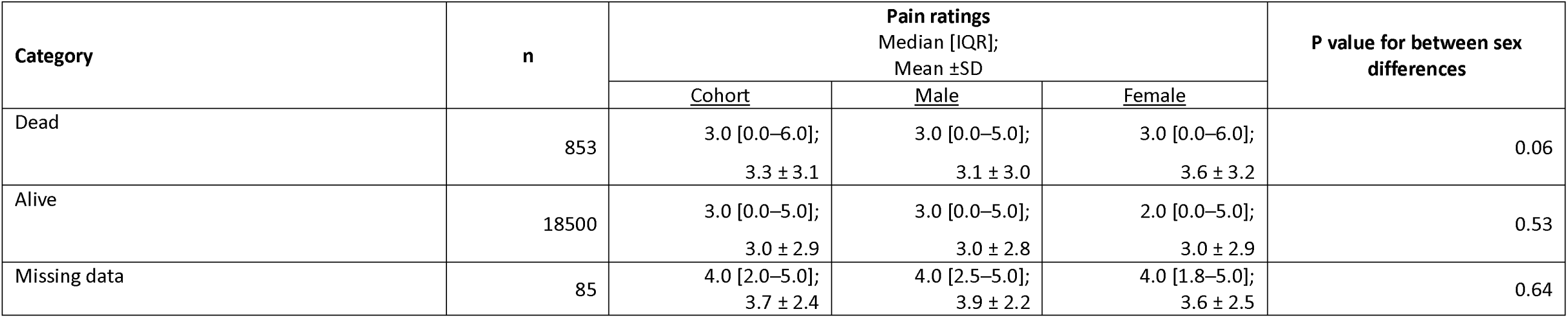
Summary of pain severity ratings between males and females. P-values are rounded to 2 significant figures.

| Category | n | Pain ratings<br>Median [IQR];<br>Mean ±SD | | | P value for between sex<br>differences |
| --- | --- | --- | --- | --- | --- |
|  |  | Cohort | Male | Female |  |
| All patients | 19438 | 3.0 [0.0–5.0];<br>3.0 ± 2.9 | 3.0 [0.0–5.0];<br>3.0 ± 2.8 | 3.0 [0.0–5.0];<br>3.0 ± 2.9 | 0.74 |
| **Age categories (years)** |  |  |  |  |  |
| 18 – 30 | 5510 | 3.0 [0.0–5.0];<br>3.1 ± 2.9 | 3.0 [0.0–5.0];<br>3.1 ± 2.8 | 2.0 [0.0–5.0];<br>3.0 ± 3.0 | 0.02 |
| 31 – 40 | 4296 | 3.0 [0.0–5.0];<br>3.1 ± 2.9 | 3.0 [0.0–5.0];<br>3.2 ± 2.8 | 2.0 [0.0–5.0];<br>3.0 ± 3.0 | 0.01 |
| 41 – 50 | 2779 | 3.0 [0.0–5.0];<br>3.1 ± 2.9 | 3.0 [0.0–5.0];<br>3.1 ± 2.9 | 3.0 [0.0–5.0];<br>3.1 ± 2.9 | 0.71 |
| 51 – 60 | 2507 | 2.0 [0.0–5.0];<br>3.0 ± 2.9 | 2.0 [0.0–5.0];<br>2.9 ± 2.9 | 3.0 [0.0–5.0];<br>3.0 ± 2.8 | 0.17 |
| 61 – 70 | 2262 | 2.0 [0.0–5.0];<br>2.8 ± 2.8 | 2.0 [0.0–4.0];<br>2.7 ± 2.7 | 2.0 [0.0–5.0];<br>2.9 ± 2.9 | 0.10 |
| 71 – 80 | 1405 | 2.0 [0.0–5.0];<br>2.8 ± 2.7 | 2.0 [0.0–5.0];<br>2.7 ± 2.7 | 3.0 [0.0–5.0];<br>2.9 ± 2.7 | 0.21 |
| 81 – 90 | 555 | 2.0 [0.0–5.0];<br>2.7 ± 2.8 | 2.0 [0.0–4.0];<br>2.5 ± 2.6 | 2.0 [0.0–5.0];<br>2.8 ± 2.9 | 0.36 |
| >90 | 111 | 3.0 [0.0–5.5];<br>3.2 ± 2.7 | 2.0 [0.0–4.8];<br>2.8 ± 2.6 | 3.0 [0.0–6.0];<br>3.5 ± 2.8 | 0.26 |
| Missing data | 13 | 2.0 [0.0–3.0];<br>2.4 ± 2.5 | 2.0 [0.0–7.0];<br>3.2 ± 3.6 | 2.0 [0.5–2.0];<br>1.5 ± 2.1 | 0.70 |
| Pregnant | | 3.0 [0.0–5.0];<br>3.0 ± 2.9 | - | 3.0 [0.0–5.0];<br>3.0 ± 2.9 | - |

| Category | n | Pain ratings<br>Median [IQR];<br>Mean ±SD |  |  | P value for between sex<br>differences |
| --- | --- | --- | --- | --- | --- |
|  |  | Cohort | Male | Female |  |
| Known comorbidities |  |  |  |  |  |
| Hypertension |  |  |  |  |  |
| Yes | 4869 | 2.0 [0.0–5.0];<br>2.7 ± 2.8 | 2.0 [0.0–5.0];<br>2.6 ± 2.8 | 2.0 [0.0–5.0];<br>2.8 ± 2.8 | 0.005 |
| No | 14569 | 3.0 [0.0–5.0];<br>3.1 ± 2.9 | 3.0 [0.0–5.0];<br>3.1 ± 2.8 | 3.0 [0.0–5.0];<br>3.1 ± 2.9 | 0.10 |
| Diabetes |  |  |  |  |  |
| Yes | 2595 | 2.0 [0.0–5.0];<br>2.8 ± 2.8 | 2.0 [0.0–5.0];<br>2.8 ± 2.9 | 2.0 [0.0–5.0];<br>2.8 ± 2.8 | 0.60 |
| No | 16843 | 3.0 [0.0–5.0];<br>3.0 ± 2.9 | 3.0 [0.0–5.0];<br>3.0 ± 2.8 | 3.0 [0.0–5.0];<br>3.1 ± 2.9 | 0.89 |
| Cancer |  |  |  |  |  |
| Yes | 1204 | 3.0 [0.0–6.0];<br>3.5 ± 2.9 | 3.0 [0.0–5.0];<br>3.2 ± 2.8 | 3.0 [0.0–6.0];<br>3.6 ± 3.0 | 0.05 |
| No | 18234 | 3.0 [0.0–5.0];<br>3.0 ± 2.9 | 3.0 [0.0–5.0];<br>3.0 ± 2.8 | 2.0 [0.0–5.0];<br>3.0 ± 2.9 | 0.32 |
| COPD/asthma |  |  |  |  |  |
| Yes | 770 | 2.0 [0.0–5.0];<br>2.9 ± 3.0 | 2.0 [0.0–5.0];<br>2.9 ± 3.0 | 2.0 [0.0–5.0];<br>2.8 ± 3.0 | 0.38 |
| No | 18668 | 3.0 [0.0–5.0];<br>3.0 ± 2.9 | 3.0 [0.0–5.0];<br>3.0 ± 2.8 | 3.0 [0.0–5.0];<br>3.0 ± 2.9 | 0.83 |
| Heart disease |  |  |  |  |  |
| Yes | 1252 | 1.5 [0.0–5.0];<br>2.5 ± 2.9 | 1.0 [0.0–5.0];<br>2.5 ± 2.9 | 2.0 [0.0–5.0];<br>2.5 ± 2.9 | 0.83 |
| No | 18186 | 3.0 [0.0–5.0];<br>3.0 ± 2.9 | 3.0 [0.0–5.0];<br>3.0 ± 2.8 | 3.0 [0.0–5.0];<br>3.0 ± 2.9 | 0.56 |
| HIV/AIDS |  |  |  |  |  |
| Yes | 2147 | 3.0 [0.0–5.0];<br>3.1 ± 3.0 | 3.0 [0.0–5.0];<br>3.1 ± 2.9 | 3.0 [0.0–5.0];<br>3.1 ± 3.0 | 0.90 |
| No | 17291 | 3.0 [0.0–5.0];<br>3.0 ± 2.9 | 3.0 [0.0–5.0];<br>3.0 ± 2.8 | 3.0 [0.0–5.0];<br>3.0 ± 2.9 | 0.65 |
| Tuberculosis |  |  |  |  |  |
| Yes | 645 | 2.0 [0.0–5.0];<br>2.8 ± 2.8 | 2.0 [0.0–5.0];<br>2.8 ± 2.8 | 2.0 [0.0–4.0];<br>2.7 ± 2.7 | 0.44 |
| No | 18793 | 3.0 [0.0–5.0]; | 3.0 [0.0–5.0]; | 3.0 [0.0–5.0]; | 0.76 |
|  |  | Cohort | Male | Female |  |
|  |  | 3.0 ± 2.9 | 3.0 ± 2.8 | 3.0 ± 2.9 |  |
| <u>Other</u> |  |  |  |  |  |
| Yes | 2843 | 2.0 [0.0–5.0];<br>2.9 ± 2.9 | 2.0 [0.0–5.0];<br>2.7 ± 2.8 | 3.0 [0.0–5.0];<br>3.0 ± 2.9 | **0.03** |
| No | 16595 | 3.0 [0.0–5.0];<br>3.0 ± 2.9 | 3.0 [0.0–5.0];<br>3.0 ± 2.8 | 3.0 [0.0–5.0];<br>3.0 ± 2.9 | 0.16 |
| <u>None</u> |  |  |  |  |  |
| Yes | 6606 | 3.0 [0.0–5.0];<br>3.2 ± 2.8 | 3.0 [0.0–5.0];<br>3.2 ± 2.8 | 3.0 [0.0–5.0];<br>3.1 ± 2.9 | **0.03** |
| No | 12832 | 2.0 [0.0–5.0];<br>2.9 ± 2.9 | 2.0 [0.0–5.0];<br>2.8 ± 2.8 | 2.0 [0.0–5.0];<br>3.0 ± 2.9 | **0.004** |
| **Urgency of admission** |  |  |  |  |  |
| Elective | 4610 | 2.0 [0.0–5.0];<br>2.7 ± 2.8 | 2.0 [0.0–4.0];<br>2.6 ± 2.7 | 2.0 [0.0–5.0];<br>2.8 ± 2.9 | 0.49 |
| Emergency/acute | 14740 | 3.0 [0.0–5.0];<br>3.1 ± 2.9 | 3.0 [0.0–5.0];<br>3.1 ± 2.8 | 3.0 [0.0–5.0];<br>3.1 ± 2.9 | 0.91 |
| Missing data | 88 | 4.0 [0.0–5.0];<br>3.3 ± 2.8 | 2.5 [0.0–5.0];<br>2.9 ± 3.1 | 4.0 [1.0–5.0];<br>3.6 ± 2.7 | 0.29 |
| **Admission diagnosis** |  |  |  |  |  |
| Non-communicable disease | 9102 | 2.0 [0.0–5.0];<br>2.7 ± 2.8 | 2.0 [0.0–5.0];<br>2.6 ± 2.8 | 2.0 [0.0–5.0];<br>2.8 ± 2.9 | 0.06 |
| Maternal health | 3723 | 2.0 [0.0–5.0];<br>2.9 ± 3.0 | - | 2.0 [0.0–5.0];<br>3.0 ± 3.0 | - |
| Trauma | 3393 | 4.0 [2.0–6.0];<br>3.9 ± 2.8 | 4.0 [2.0–6.0];<br>3.8 ± 2.7 | 4.0 [2.0–6.0];<br>4.3 ± 2.7 | **<0.001** |
| Infection | 3157 | 3.0 [0.0–5.0];<br>3.0 ± 2.8 | 2.0 [0.0–5.0];<br>2.8 ± 2.8 | 3.0 [0.0–5.0];<br>3.1 ± 2.8 | **0.02** |
| Missing data | 63 | 1.0 [0.0–5.0];<br>2.0 2.5 ± 2.9 | 1.0 [0.0–3.0];<br>2.0 1.9 ± 2.5 | 2.0 [0.0–6.0];<br>3.1 ± 3.2 | 0.17 |
| **Ward type** |  |  |  |  |  |
|  |  | Cohort | Male | Female |  |
| Maternal | 3655 | 2.0 [0.0–5.0];<br>3.0 ± 3.0 | - | 2.0 [0.0–5.0];<br>3.0 ± 3.0 | - |
| Medical | 7179 | 2.0 [0.0–4.0];<br>2.5 ± 2.8 | 2.0 [0.0–4.0];<br>2.4 ± 2.7 | 2.0 [0.0–4.0];<br>2.5 ± 2.8 | 0.23 |
| Surgical | 7341 | 3.0 [1.0–5.0];<br>3.4 ± 2.8 | 3.0 [1.0–5.0];<br>3.3 ± 2.8 | 3.0 [1.0–6.0];<br>3.5 ± 2.9 | **0.03** |
| Other | 1257 | 3.0 [0.0–6.0];<br>3.6 ± 3.1 | 4.0 [0.0–6.0];<br>3.8 ± 3.1 | 3.0 [0.0–6.0];<br>3.5 ± 3.1 | 0.12 |
| Missing data | 6 | 3.5 [1.5–5.5];<br>3.5 ± 2.7 | 6.0 [3.0–6.5];<br>4.3 ± 3.8 | 3.5 [3.2–3.8];<br>3.5 ± 0.7 | 0.77 |
| **Ward level** |  |  |  |  |  |
| General ward | 17345 | 2.0 [0.0–5.0];<br>3.0 ± 2.9 | 3.0 [0.0–5.0];<br>2.9 ± 2.8 | 2.0 [0.0–5.0];<br>3.0 ± 2.9 | 0.50 |
| High care | 1341 | 3.0 [1.0–6.0];<br>3.6 ± 3.0 | 3.0 [0.0–6.0];<br>3.6 ± 3.0 | 3.0 [1.0–6.0];<br>3.7 ± 3.0 | 0.78 |
| ICU | 701 | 3.0 [0.0–5.0];<br>2.9 ± 2.9 | 2.0 [0.0–5.0];<br>2.8 ± 2.9 | 3.0 [0.0–5.0];<br>3.1 ± 2.9 | 0.21 |
| Missing data | 51 | 4.0 [0.5–5.0];<br>3.4 ± 2.7 | 4.0 [1.5–5.0];<br>3.7 ± 2.9 | 4.0 [0.0–5.0];<br>3.3 ± 2.6 | 0.73 |
| **Critical illness status** |  |  |  |  |  |
| Critically illness | 2232 | 3.0 [0.0–5.0];<br>3.1 ± 3.0 | 3.0 [0.0–5.0];<br>3.0 ± 3.0 | 3.0 [0.0–5.0];<br>3.1 ± 3.1 | 0.45 |
| Not critically ill | 17088 | 3.0 [0.0–5.0];<br>3.0 ± 2.8 | 3.0 [0.0–5.0];<br>3.0 ± 2.8 | 3.0 [0.0–5.0];<br>3.0 ± 2.9 | 0.62 |
| Missing data | 118 | 3.0 [0.0–6.0];<br>3.4 ± 3.2 | 4.0 [0.0–6.5];<br>3.8 ± 3.4 | 2.5 [0.0–5.0];<br>3.0 ± 3.0 | 0.15 |
| **Survival status** |  |  |  |  |  |
|  |  | <u>Cohort</u> | <u>Male</u> | <u>Female</u> |  |
| Dead | 853 | 3.0 [0.0–6.0];<br>3.3 ± 3.1 | 3.0 [0.0–5.0];<br>3.1 ± 3.0 | 3.0 [0.0–6.0];<br>3.6 ± 3.2 | 0.06 |
| Alive | 18500 | 3.0 [0.0–5.0];<br>3.0 ± 2.9 | 3.0 [0.0–5.0];<br>3.0 ± 2.8 | 2.0 [0.0–5.0];<br>3.0 ± 2.9 | 0.53 |
| Missing data | 85 | 4.0 [2.0–5.0];<br>3.7 ± 2.4 | 4.0 [2.5–5.0];<br>3.9 ± 2.2 | 4.0 [1.8–5.0];<br>3.6 ± 2.5 | 0.64 |

Within the age categories of 18 to 30 years old and 31 to 40 years old, the distribution of pain severity ratings reflected that males reported slightly higher pain scores than females, on average (ages 18 – 30 p = 0.02; ages 31 – 40 p = 0.01). Among patients with hypertension, the distribution of pain severity ratings reflected that females reported slightly higher pain scores than males, on average (p = 0.005). Among the patients admitted for trauma or for infection, or those receiving care in the surgical wards, the distribution of pain severity ratings reflected that females reported slightly higher pain scores than males, on average (trauma p <0.001; infection p = 0.02; surgical ward p = 0.03). Notably, these statistical differences in pain severity between males and females were small in magnitude. With similar medians and largely overlapping distributions, suggesting that they are unlikely to be clinically meaningful. Details can be found in Table 3.

#### 3.3.3 Prevalence of severe pain (i.e. ≥7 out of 10)

Of the 19 438 patients, the average prevalence [95% CI] of severe pain (≥7/10) across participating hospitals was 14.4% [13.9–14.9] (Table 2). Hospital-level prevalences of severe pain ranged from 0% to 66.7%. Severe pain was slightly more common among patients admitted for emergency/acute care than those admitted for elective care (15.1% vs 12.2%; absolute difference +2.8 percentage points). Across the four categories of admission, severe pain prevalence was highest among patients admitted for trauma (20.3%; absolute difference between trauma and non-communicable diseases: +7.8 percentage points). Severe pain prevalence was also higher in surgical wards (16.2%) than in medical (11.1%; absolute difference between surgical and medical wards: +5.2 percentage points) or maternal wards (14.4%), and higher in high-care units (19.3%) than in general wards (14.0%; absolute difference between high-care and general wards: +5.4 percentage points) or intensive care units (15.4%). Details can be found in Table 2.

### 3.4 Secondary aims

#### 3.4.1 Association between pain rating (0 – 10) and critical illness status

The multivariable logistic regression model demonstrated no evidence of an association between pain severity ratings and critical illness status (OR 1.01, 95%CI 0.99–1.03, p = 0.19) (Table 4; model diagnostics and fit are presented in Supplementary Material S12).

**Table 4:** Adjusted three-level (nested) generalised logistic mixed model of the relationship between pain rating (0 – 10) and (i) critical illness status and (ii) in-hospital seven-day mortality. P-values are rounded to 2 significant figures.

|  | Outcome: critical illness |  |  | Outcome: mortality |  |  |
| --- | --- | --- | --- | --- | --- | --- |
|  | Odds ratio | 95% confidence interval | p value | Odds ratio | 95% confidence interval | p value |
| Intercept (odds) | 0.02 | 0.02 – 0.03 | <0.0001 | 0.0006 | 0.0003 – 0.0011 | <0.0001 |
| Pain severity (0 – 10) | 1.01 | 0.99 – 1.03 | 0.19 | 1.05 | 1.02 – 1.07 | 0.0009 |
| Age per 10 years | 1.07 | 1.04 – 1.11 | <0.0001 | 1.23 | 1.18 – 1.29 | <0.0001 |
| Pregnant | 0.87 | 0.66 – 1.14 | 0.30 | - | - | - |
| Known comorbidities |  |  |  |  |  |  |
| Hypertension | 1.00 | 0.89 – 1.13 | 0.98 | - | - | - |
| Diabetes | 0.95 | 0.82 – 1.09 | 0.47 | - | - | - |
| Cancer | 1.26 | 1.05 – 1.52 | 0.016 | 2.53 | 2.00 – 3.20 | <0.0001 |
| COPD/ Asthma | 2.13 | 1.77 – 2.56 | <0.0001 | - | - | - |
| Heart disease | 1.60 | 1.36 – 1.88 | <0.0001 | - | - | - |
| HIV/AIDS | 1.31 | 1.12 – 1.54 | 0.0007 | 1.39 | 1.09 – 1.76 | 0.0070 |
| Tuberculosis | 2.06 | 1.66 – 2.55 | <0.0001 | - | - | - |
| Other | 1.20 | 1.06 – 1.36 | 0.0048 | - | - | - |
| Urgency of admission |  |  |  |  |  |  |
| Emergency/ acute | 2.44 | 2.12 – 2.82 | <0.0001 | 3.25 | 2.47 – 4.28 | <0.0001 |
| Elective | Reference |  |  |  |  |  |
| Main category for admission |  |  |  |  |  |  |
| Infection | 1.32 | 1.02 – 1.71 | 0.034 | 3.30 | 1.91 – 5.72 | <0.0001 |
| Non-communicable disease | 1.83 | 1.44 – 2.32 | <0.0001 | 6.38 | 3.82 – 10.66 | <0.0001 |
| Trauma | 2.14 | 1.66 – 2.75 | <0.0001 | 7.88 | 4.67 – 13.28 | <0.0001 |
| Maternal health | Reference |  |  |  |  |  |
| Critical illness | - | - | - | 6.95 | 5.95 – 8.13 | <0.0001 |

#### 3.4.2 Association between severe rating (0 – 10) and in-hospital seven-day mortality

The multivariable logistic regression model demonstrated that increasing pain severity was associated with increased odds of dying within seven days (OR 1.05, 95%CI 1.02–1.07, p = 0.0009) (Table 4; model diagnostics and fit are presented in Supplementary Material S13).

##### Kaplan-Meier survival plot comparing in-hospital mortality over seven days for patients with and without severe pain

The Kaplan-Meier plot included 2 603 (95.17%) of 2 735 patients with severe pain (1 964 discharged, and 639 in hospital at day seven), and 5 666 (89.95%) of 6 299 patients without severe pain (1 722 discharged, and 3 944 in hospital at day seven). The Kaplan-Meier Plot (Supplementary Material S14) suggested that patients with severe pain (≥7/10) were significantly more likely to die within seven days than patients with non-severe pain (<7/10) (p = 0.023).

### 3.5 Sensitivity analysis

Two countries (Nigeria and South Africa) each contributed more than 10% of the total sample. The findings of excluding these two countries from the multivariable logistic regression analyses for the secondary aim were consistent with the main findings: there was no evidence of an association between pain severity ratings and critical illness status (OR: 1.02, 95%CI 0.99-1.04, p = 0.14), and increasing pain severity rating was associated with increased odds of dying within seven days (OR: 1.04, 95%CI: 1.01-1.08, p = 0.01) (Supplementary Material S15).

### 3.6 Exploratory analyses

#### 3.6.1 Odds of severe pain by sex and age

There was no evidence of an association between sex and the odds of severe pain (OR 0.97, 95%CI 0.89 – 1.06; p = 0.47; Supplementary Material S16). However, increasing age was significantly associated with lower odds of having severe pain. A one-standard-deviation increase in age was associated with an 11% reduction in the odds of having severe pain (OR 0.89, 95% CI 0.84–0.95; p = 0.0001). There was no evidence of an interaction between sex and age (p = 0.20), suggesting that the association between age and severe pain status did not differ by sex.

#### 3.6.2 Association between severe pain status and critical illness

Severe pain status (≥7/10) was associated with critical illness status (x^2^ (1) = 14.5, p = 0.0001). The multivariable logistic regression model demonstrated that patients with severe pain had 30% greater odds of being classified as critically ill (OR 1.31, 95% CI 1.15–1.48, p < 0.0001) (Supplementary Material S17).

#### 3.6.3 Association between severe pain status and in-hospital seven-day mortality

Severe pain status was associated with in-hospital seven-day mortality (x^2^ (1) = 7.7, p = 0.0055). The multivariable logistic regression model demonstrated that patients with severe pain had 27% greater odds of dying within seven days (OR 1.27, 95% CI 1.04–1.56, p = 0.020) (Supplementary Material S18).

## 4. Discussion

This study provides, to the best of our knowledge, the first multi-centre estimates of the prevalence and severity of acute pain among hospitalised adults across 22 African countries or territories. In this large sample of 19 438 hospitalised patients, two-thirds reported pain and one in seven reported severe pain (≥7/10). Pain was more common among patients admitted for emergency or trauma care, and in surgical and high-care wards. Pain severity did not differ meaningfully between sexes overall, although small differences were observed within specific subgroups. There was no evidence of an association between increasing pain severity and critical illness; however, increasing pain severity was associated with a higher likelihood of in-hospital mortality within seven days.

The prevalence and severity of pain observed in this study are broadly consistent with reports from high-income settings, where estimates of pain prevalence range from 38% to 84% and of severe pain from 9% to 36% among hospitalised patients [23]. In our cohort, 67.9% of patients reported pain and 14.4% reported severe pain. This similarity is notable given substantial differences in healthcare resources between high-income countries and many hospitals in Africa, including fewer healthcare workers, limited access to analgesics, and reduced availability of specialised acute pain services [3, 12, 33, 56].

The finding that one in seven hospitalised patients in Africa report severe pain is clinically important. Severe acute pain has been associated with delayed recovery and adverse clinical outcomes, including increased risk of developing persistent pain [15, 16, 18, 22, 41, 47]. It likely also contributes to psychological distress and reduced quality of life. At a systems level, poorly controlled pain has been linked to longer hospital stays, higher readmission rates, and increased healthcare costs [16, 22, 27]. Perhaps, especially in healthcare systems with strained resources, treating severe pain is an opportunity to considerably improve the quality of patient care while potentially also reducing system burden.

Improving acute pain management in hospitalised patients in Africa is likely to require coordinated clinical, research, and policy efforts. A recent international Delphi study identified key research priorities for postoperative pain in African settings, including routine pain management practices, predictors of acute and chronic postsurgical pain, facilitators and barriers to effective care, patients’ satisfaction with pain management, and low-cost, low-resource strategies for improving pain management [3]. Although these priorities focus specifically on postoperative pain, they broadly reflect that pain in hospitalised patients is also likely shaped by a complex interplay of clinical, system-level, and sociocultural factors. Barriers to optimal pain management in African settings include limited access to medications (especially opioids), limited access to the equipment and skills required for regional nerve blocks, and workforce shortages [3]. In parallel, pain expression and reporting are influenced by social norms, expectations of care, and patient–provider dynamics [35, 38, 40]. How these factors interact or potentially oppose each other to influence pain reporting and pain severity in African settings in not yet known. We would therefore add to the list of research priorities exploration of the full range of context-, population-, and person-specific factors that influence acute pain and its management in African hospitals. In addition, more detailed characterisation of pain, including the type, location, and causes of pain, access to and use of analgesia, and the functional and psychological impact of pain, would guide the development of targeted and context-appropriate interventions. Therefore, while the current study provides a much-needed foundation of multi-country data on the burden of acute pain among hospitalised patients in Africa, much context-specific data is still required to support better clinical pain care, resource allocation, and pain policies on the continent. In particular, we advocate for research on pain characterisation, the availability, acceptability, use, and effectiveness of pharmacological and behavioural treatments for pain, and system-level barriers to pain care.

Although the current study observed an association between increasing pain severity and in-hospital seven-day mortality, we believe this finding should be interpreted with caution. The observational design precludes causal inference, and pain severity may reflect underlying disease severity rather than directly influencing mortality. Although the association remained after adjustment for independent predictors of mortality and in the sensitivity analysis, residual confounding, such as access to timely treatment or underlying disease processes, is likely. Previous work from high-income settings has also reported associations between poorly controlled pain and mortality [1], but the direction and mechanisms of these relationships remain unclear. Further research is needed to determine whether pain is a marker of risk, a contributor to adverse outcomes, or both.

This study had several strengths. It included a large cohort of hospitalised patients across 22 African countries and territories with minimal missing data. Of the 19 438 patients included, no patient-level variable had a missingness rate over 0.6%. Pain was assessed using standardised questions, with pre-specified definitions and analysis plans, aligning with principles of transparent and reproducible research. Therefore, this study provides a robust and generalisable dataset to inform ongoing research and improvements to pain management and health policy in Africa.

This study has four key limitations. First, the visual-numerical scale for pain severity did not address pain-related function, nor the affective component of pain, although some work suggests that pain is a unitary experience [44] and that even numerical pain ratings contain complex information that extends beyond just intensity [14]. More comprehensive characterisation of pain should consider the location, anatomical extent, functional, psychological, and cognitive effects of pain. The scale anchor labels were translated ad hoc, leaving some uncertainty about the consistency of anchor meaning across languages. Second, this study collected no data on pain treatments administered to patients. Therefore, it cannot speak to the effectiveness of analgesic treatments in this cohort nor to how such treatments may have influenced pain prevalence and severity and its relationship to critical illness or seven-day mortality. This limits interpretation and should be considered when drawing conclusions from these findings. Third, a pragmatic selection of confounders included in the study could be considered a limitation, although the decision was based on prior epidemiological evidence supporting the association of many of the confounders with critical illness and mortality [6]. Finally, the pragmatic and intentionally limited nature of the dataset restrict our ability to meaningfully explore potential explanations for the observed lower odds of severe pain with increasing age. This finding should therefore be interpreted cautiously and warrants further investigation in studies specifically designed to interrogate age-related differences in pain experience and reporting among hospitalised patients.

## Conclusion

Acute pain is highly prevalent among hospitalised patients across African settings, with one in seven patients experiencing severe pain. These findings are comparable to data from in high-income countries, despite substantial differences in healthcare resources, suggesting that acute pain remains a persistent and under-addressed problem globally. The observed association between acute pain and in-hospital seven-day mortality should be interpreted cautiously and warrants further investigation. This study provides a foundation for future research aimed at comprehensively characterising pain, including context-specific factors that influence pain severity and reporting, availability, use, and effectiveness of pharmacological and behavioural treatments for pain, and system-level barriers to pain care in African settings.

## Data sharing statement

Data sharing requests are welcome from bona fide researchers, and will be considered by the ACIOS Steering Committee.

## Conflicts of interest

GJB is supported by a Carnegie DEAL 3 Postdoctoral Fellowship (G-21-58838), and receives speakers’ fees for talks on pain and rehabilitation.

RMP has received research grants and/or honoraria from Edwards Lifesciences and Intersurgical UK.

RP receives speakers’ fees for talks on pain and rehabilitation, is a director of the not-for-profit organisation, Train Pain Academy, and serves as a councillor for the International Association for the Study of Pain.

VJM has received speakers’ fees for talks on pain and rehabilitation and is an associate director of the not-for-profit organisation, Train Pain Academy.

TB declares technical consultancies with UNICEF, the World Bank, USAID, and PATH, all outside the submitted work.

