## Supplementary material for "One in seven has severe pain: A point-prevalence study of hospitalised patients across Africa"

1. Gillian J Bedwell, MSc, Department of Anaesthesia and Perioperative Medicine, Groote Schuur Hospital, Faculty of Health Sciences, University of Cape Town, Cape Town, South Africa.
2. Professor Romy Parker, PhD, Professor, Department of Anaesthesia and Perioperative Medicine, Groote Schuur Hospital, Faculty of Health Sciences, University of Cape Town, Cape Town, South Africa.
3. Victoria J Madden, PhD, Associate Professor, Department of Anaesthesia and Perioperative Medicine, Groote Schuur Hospital, Faculty of Health Sciences, University of Cape Town, Cape Town, South Africa.
4. Juan Scribante, PhD, Associate Professor, Surgeons for Little Lives, Department of Paediatric Surgery, University of the Witwatersrand, Johannesburg, South Africa.
5. Muhammed Elhadi, MBBCh, Faculty of Medicine, University of Tripoli, Tripoli, Libya.
6. Professor Adesoji O Ademuyiwa, FACS, Professor of Surgery (Paediatric and Surgical Epidemiology), Department of Surgery, College of Medicine, University of Lagos and Lagos University Teaching Hospital, Lagos, Nigeria.
7. Professor Babatunde Osinaike, FMCA, Professor of Anaesthesia and Intensive Care, University of Ibadan/University College Hospital, Ibadan, Oyo State, Nigeria.
8. Christian Owoo, MV, MBChB, FGCS, MPH, Senior lecturer, University of Ghana Medical School; Director of the Medical Training and Simulation Centre, University of Ghana Medical Centre; and Consultant Anaesthesiologist & Head of ICU, Korle Bu Teaching Hospital, Accra, Ghana.
9. Daniel Sottie, MBChB, DA (WACS), FGCS (Anaesthesia), Korle-Bu Teaching Hospital, Accra, Ghana.
10. Karima Khalid, MD, MMed, Department of Anaesthesia and Intensive Care, Muhimbili University of Health and Allied Sciences, Dar es Salaam, Tanzania.
11. Adam Hewitt-Smith, MBBS, Lecturer, Faculty of Health Sciences, Busitema University, Busitema, Uganda, Queen Mary University of London, United Kingdom, EC1M 6BQ.
12. Arthur Kwizera, MBChB, MMed, Makere University, Kampala, Uganda.
13. Fitsum Kifle Belachew MSc Med, Network for Perioperative and Critical Care, Debre Berhan University Asrat Woldeyes Health Sciences Campus, Debre Berhan, Ethiopia and Global Surgery Division, Department of Surgery, University of Cape Town, Cape Town, South Africa.
14. Degisew Dersso Mengistu, MPH, National Emergency, Injury and Critical Care Program Coordinator, Ministry of Health, Ethiopia
15. Yared Boru Firissa, MD, ALERT Comprehensive Specialized Hospital, Addis Ababa, Ethiopia.
16. Tirunesh Busha Gemechu, MD, Honorary staff, Black Lion Hospital, Addis Ababa, Ethiopia.
17. Gaudencia Dausab, MMed, Ministry of health and social services Intermediate Hospital Katutura Consultant, Windhoek, Namibia.
18. Unotjari Kauta, MBBS, Ministry of Health and Social Services, Senior Medical Officer in the Medical Intensive Care Unit Intermediate Hospital Katutura, Windhoek, Namibia.
19. Kaveto Sikuvi, FCEM (SA), Ministry of Health Namibia, Lecturer, University of Namibia School of Medicine, Windhoek, Namibia.
20. Nahla Kechiche, MD, Associate Professor, Department of Pediatric Surgery, University Hospital of Monastir, Monastir, Tunisia.
21. Kelan Bertille Ki, MMed, Associate Professor, Joseph Ki-Zerbo University, CHU Pédiatrique Charles de Gaulle, Ouagadougou, Burkina Faso.
22. Martin Mukenga, MD, Specialist in Anesthesiology, University Hospital of Kinshasa, Kinshasa, Democratic Republic of Congo.
23. Dolly Munlemvo, MD, Specialist in Anesthesiology, University Hospital of Kinshasa, Kinshasa, Democratic Republic of Congo.
24. Mustapha Bittaye, FWACS, University of The Gambia, SerreKunda. The Gambia.
25. Abubacarr Jagne, MBchB, Fellow WACP and ISN fellow of Nephrology, Deputy Chief Medical Director and Chair of Medical Advisory Committee EFSTH, and lecturer, University of The Gambia, SerreKunda, The Gambia.
26. Mohamed Abdinor Omar, MBBS, Global Surgery Emergency Department Federal Ministry of health and Human Services Somalia, Mogadishu, Somalia.
27. Hassan Ali Daoud, MD, Amoud University, Geneva Graduate Institute, Somaliland.
28. Mohamed Faisal, MD, College of Medicine and Health Science, University of Hargeisa. Somaliland Emergency Medicine Association, Hargeisa, Somaliland.
29. Professor Mahmoud Elfiky MD, Professor, Cairo University, Cairo, Egypt.
30. Mpho Seleke, MBBCh, District Medical Officer, Ministry of Health, Lesotho.
31. Tarig Fadalla, MBBS, Ribat University Hospital, The National Ribat University, Khartoum, Sudan.
32. Alshaima Koko, MBBS, Pediatric Surgery Center, Ribat University Hospital Khartoum, Sudan.
33. Alemayehu G Bedada, MD, Associate Professor, University of Botswana, Gaborone, Botswana.
34. Gilles Niengo Outsouta, MD, Hospital Practitioner in Polyvalent ICU at CHU Brazzaville, Brazzaville, Congo.
35. Marie Elombila, Professeur Agrégé, Université Marien Ngouabi CHU de Brazzaville, Brazzaville, Congo.
36. Professor Ahmed Rhassane El Adib, MD, Professor and Head of Anesthesia and Critical Care in Gynecology-Obstetrics Department, Mohammed VI University Hospital, Cadi Ayyad University, Marrakesh. Dean of Faculty of Medicine, Mohammed VI University of Sciences and Health, Casablanca, Morocco.
37. Meryem Essafti, MD, Assistant professor, Anesthesia and Critical Care in Gynecology-Obstetrics Department, Mother and Child Hospital, Mohammed VI University Hospital, Faculty of Medicine and Pharmacy of Marrakech, Cadi Ayyad University of Morocco.
38. Dino Lopes, MD, Maputo Central Hospital, Tutor of the Emergency Medicine residency, Maputo, Mozambique.
39. Atilio Morais, PhD, Director de Departamento de Cirurgias, Hospital Central de Maputo, Faculdade de Medicina, Universidade Eduardo Mondlane, Maputo, Mozambique
40. Pisirai Ndarukwa, PhD, Bindura University of Science Education, Bindura, Zimbabwe and University of KwaZulu Natal, KwaZulu Natal, South Africa.
41. Newten Handireketi, MSc, Bindura University of Science Education and National Institute of Health Research, Ministry of Health and Child Care-Zimbabwe, Harare, Zimbabwe.
42. Fred Bulamba, MMed, Department of Anaesthesia and Critical Care, Faculty of Health Sciences, Busitema University, Tororo, Uganda, William Harvey Research institute, Queen Mary University of London, London, United Kingdom.
43. Busisiwe Mrara, MMed, Department of Anaesthesiology, Nelson Mandela Academic Hospital, Walter Sisulu University (WSU), Mthatha, South Africa
44. Professor Hyla-Louise Kluyts, MMed (Anaes), PhD, Department of Anaesthesiology, Sefako Makgatho Health Sciences University, Ga-Rankuwa, South Africa.
45. Marian Kinnes, Department of Anaesthesia and Perioperative Medicine, Groote Schuur Hospital, Faculty of Health Sciences, University of Cape Town, Cape Town, South Africa.
46. Abdullahi Said Hashi,^,^ MD, Department of Anesthesiology and Reanimation, Mogadishu Somali-Türkiye Recep Tayyip Erdoğan Training and Research Hospital, Mogadishu, Somalia.
47. Abigail Kusi Amponsah, PhD, Senior Lecturer, School of Nursing and Midwifery, Kwame Nkrumah University of Science and Technology, Kumasi, Ghana.
48. Professor Aderonke Omobonike Akinpelu, PhD, Professor, Department of Physiotherapy, Faculty of Clinical Sciences, College of Medicine, University of Ibadan, Ibadan, Nigeria.
49. Andrew Amata, FMCA, Consultant Anaesthesiologist, Beit-CURE International Hospital, Lusaka, Zambia.
50. Aunel Mallier Peter, DNB, lecturer, Department of Anaesthesia, University of the Witwatersrand, Johannesburg, South Africa
51. Professor Elizabeth Nwasor, MBBS, DA (WACS), FWACS, MPH, PFDA (WFSA), Professor, Department of Anaesthesia, Ahmadu Bello University Teaching Hospital, Zaria, Nigeria.
52. Jeremie Kalambay, MD, Specialist in Anaesthesiology, Department of Anaesthesia, University Hospital of Kinshasa, Kinshasa, Democratic Republic of Congo.
53. Hellen N Kariuki, PhD, senior lecture, Department of Medical Physiology, University of Nairobi, Nairobi, Kenya.
54. Nana Fening, MMed (Wits), Anaesthesiologist, Department of Anaesthesiology, Faculty of Health Sciences, University of the Witwatersrand, Johannesburg, South Africa.
55. Nomaqhawe Moyo, MSc, senior lecturer, Department of Anaesthesia and Critical Care, University of Zimbabwe, Harare, Zimbabwe.
56. Patrick Mukuna, MD, Specialist in Anaesthesiology, Department of Anaesthesia, University Hospital of Kinshasa, Kinshasa, Democratic Republic of Congo.
57. Sean Chetty, PhD, Executive Head of Department, Anaesthesiology & Critical Care, Faculty of Medicine and Health Sciences, Stellenbosch University, Cape Town, South Africa.
58. Sudarshanie Bechan, FCA, Head Clinical Unit, Inkosi Albert Luthuli Central Hospital and University of KwaZulu-Natal, Durban, South Africa.
59. Tania Pretorius, FCA(SA) MMed Anaesthesia, Honorary lecturer, Department of Anaesthesia and Perioperative Medicine, University of Cape Town, Cape Town, South Africa.
60. Professor Bamidele Victor Owoyele, PhD, Professor, Department of Physiology, Faculty of Basic Medical Sciences, University of Ilorin, Ilorin, Nigeria.
61. Luyanduthando Mqadi, MSc, Research assistant, African Pain Research Initiative, Department of Anaesthesia and Perioperative Medicine, Neuroscience Institute, University of Cape Town, Cape Town, South Africa
62. Hanél Duvenage, MSc Molecular Medicine and Bioinformatics, MSc Nutrition, Research and Development Manager, Safe Surgery South Africa.
63. Gwendoline Arendse, MPH, research assistant, Department of Anaesthesia and Perioperative Medicine, Groote Schuur Hospital, Faculty of Health Sciences, University of Cape Town, Cape Town, South Africa.
64. Luke Hannan, MPH, statistical consultant, Division of Epidemiology and Biostatistics, School of Public Health, Faculty of Health Sciences, University of Cape Town, South Africa.
65. Professor Landon Myer, PhD, Professor, statistical consultant, Division of Epidemiology and Biostatistics, School of Public Health, Faculty of Health Sciences, University of Cape Town, South Africa.
66. Carl Otto Schell, PhD, Department of Global Public Health, Karolinska Institutet, Stockholm, Sweden, Centre for Clinical Research Sörmland, Uppsala University, Eskilstuna, Sweden, Department of Medicine, Nyköping Hospital, Sörmland Region, Nyköping, Sweden.
67. Tim Baker, PhD, Department of Emergency Medicine Muhimbili University of Health and Allied Sciences, Dar es Salaam, Tanzania; and Department of Global Public Health, Karolinska Institutet, Stockholm, Sweden; and Queen Mary University of London, United Kingdom and Clinical Research Department, London School of Hygiene and Tropical Medicine, United Kingdom.
68. Professor Rupert M Pearse, MD(Res), Professor of Intensive Care Medicine, Queen Mary University of London, United Kingdom, EC1M 6BQ.
69. Professor Bruce M Biccard, PhD, Nuffield Professor of Anaesthetic Science, Nuffield Department of Clinical Neurosciences, University of Oxford.

### **Steering Committee:**

**NIHR Global Health Group on Perioperative and Critical Care, co-Directors and Senior Investigators:** Rupert M Pearse, Bruce M Biccard

**Chief Investigator:** ACIOS - Tim Baker

**Statistical analysis:** Gillian J Bedwell, Bruce M Biccard, Gwendoline Arendse, Luke Hannan, Landon Myers

**Data administrators:** Hanél Duvenage

**APPRISE (A**frican **P**artnership for **P**erioperative and C**R**it**I**cal Care Re**SE**arch**) Project Management (NIHR133850):**

Mohammed Adem, Elvis Amunyo, Gwen Arendse, Tim Baker, Upendo Bandeke, Godfrey Barabona, Gillian J Bedwell, Tizeta Belachew, Ermiyas Belay, Aneel Bhangu, Bruce Biccard, David Bishop, Happines Biyengo, Yared Boru, Fred Bulamba, Turunesh Busha, Marcelle Crowther, Lucy Cunnama, Alma O Damasy, Selam Daniel, Justine Davies, Abiy Dawit, Brook Demissie, Degsew Dersso Mengistu, Yamkela Desemela, Kokeb Desta, Kelem Desta, Priyanthi Dias, Hanél Duvenage, Rowan Duys, Robert Dyer, Ryan Ellis, Charles Fadipe, Margot Flint, Alexander Fowler, Aphelele Futshane, Desta Galcha, Richard Gamubaka, Nuroadis Guchima, Simphiwe Gumede, Thomas Hamborg, Anneli Hardy, Adam Hewitt-Smith, Sabra Hussein, Anna Hvarfner, Anab F Issa, Kasandji Kabambi, Aneth Kaliza, Assen Kamwesigwe, Muzamiru Kawiso, Peniel Kenna, Karima Khalid, Justine Khanyalano, James Kidulu, Fitsum Kifle, Tewodros Kifleyohanes, Marian Kinnes, Juma Kitwara, Herbert Kiwalya, Hyla-Louise Kluyts, Charles Machumu, Edson Mafana, Peter Magala, Salome Maswime, Rev. Mzubanzi Mdunyelwa, Borislava Mihaylova, Elibariki Mkumbo, Linda Mlunde, Catherine Mokotla, Steve Molaoa, Lebogang Moloi, Jolene Moore, Busisiwe Mrara, Lwandile Mtshabe, Betelehem Mulye, Joanitah Nakibuule, Joan Naluyima, Hasifah Namutebi, Rose Nandutu, Juliana Nanimambi, Ezile Ninise, David Okanya, Rupert Pearse, Linda Pohl, Rosalind Prinsloo, Rafael S Shayo, Timothy Stephens, Nicola Vickery, Cecilia Vindrola, John Walutsyo, Bezaye Zemdkun.

**ACIOS Investigators:**

Hospital Lead Investigators*

**Botswana**

National leader: Alemayehu Ginbo Bedada

*Princess Marina Hospital*

- Shingirai Muzondiwa*

- Kagiso Wadikonyana

- Tariro Mubika

- Wabotlhe Ntwayapelo

**Burkina Faso**

National Leader: Kelan Bertille Ki

*Centre Hospitalier Universitaire Yalgado Ouédraogo, Ouagadougou (CHUYO)*

   - Abdramane Ouattara*

- Papougnezambo Bonkoungou

- Lankoundé Martin

*Centre Hospitalier Universitaire de Tengandogo, Ouagadougou (CHUT)*

    - Salam Savadogo*

- Nathanael Bamogo

- Bayé Nébié

*Centre Hospitalier Universitaire Sourô Sanou (CHUSS)*

    - Ismael Guibla*

    - Daouda Barro

    - Latifa Aida Sawadogo

- Guy Thierry Ki

- Bego-wene Isai Bado

- Ibrahim Alain Traore

*Centre Hospitalier Universitaire de Bogodogo*

- Salah Idriss Séif Traore*	- Wendelamita Lydie Rosine Kaboré

- Salifou Napon

- Arouna Louré

- Marame Diop

**Congo**

National Leader: Gilles Niengo Outsouta, Marie Elombila

*CHU de Brazzaville*

- Marie Elombila*

- Otiobanda Gilbert Fabrice

- Niengo Outsouta Gilles

- Mpoy Emy Monkessa Christ Mayick

- Iwoba Rebet Sarrah Armanda

- Koumou Banga Fernand Régis

**Democratic Republic of Congo**

National Leader: Dolly Munlemvo, Martin Mukenga

*Biamba Marie Mutombo Dikembe*

- Jean Jacques Kalongo*

*Centre Hospitalier Initiative Plus (CHIP)*

- Patrick Mukuna*

- Frank Nguvulu*

*Cliniques Universitaires de Kinshasa*

- Berthe Barhayiga*

- Patrick Mukuna

- Martin Mukenga

*Clinique Kinoise*

- Jeremie Kalambayi*

*El Rapha Clinic*

- Ted Likongo*

**Egypt**

National Leader: Mahmoud Elfiky

*Aswan University Hospital*

    - Sarah Magdy Abdelmohsen*

    - Mohie El-Din Mostafa Madany

*Giza International Hospital*

    - Ahmed Saber Abdelrahman*

    - Sarah Mansour

**Ethiopia**

National Leaders: Tirunesh Busha Gemechu, Yared Boru Firissa, Fitsum Kefle, Degisew Mengistu

*AaBET Hospital*

    - Ayalew Zewdie Tadesse*

    - Daniel Adugna Dashura

- Alemayehu Beharu

- Seble Ashagre Awoke

*Alert Hospital*

- Tsegay Gebreanenia Hagos*

- Eyuel Teshome

- Amare Yohans Mekonen

- Abnet Ewnetie Emirie

Arbaminch University Hospital

*- Desta* Galcha Gerbu*

- Weldemichael Yigebahal Belay

- Aregahegn Mulgeta Munka

- Begashaw Melesse Dicha

- Addisu Tesfaye Koster

- Moges Tessema Hesbeto

*Dilla University Teaching Hospital*

- Zerihun Tesfaye*

- Wondimu Dori

- Roba Kebede

- Mihret Kaleb

- Selamawit Yizelkal

- AbrahamT/markos

*Haramaya University Hiwot Fana Comprehensive Specialized Hospital*

- Melaku Getachew*

- Fanta Wondimneh

- Tilahun Teshager

- Gamachu Bekuma

*Hawassa Hospital*

- Emnet Tesfaye Shimber*

- Kindie Nigatu Woubshet

- Kibru Kifle Karicha

- Melaku Teshale

*Jimma University Medical Center*

- Gemechis Melkamu Fufa*

- Addis Ketemaoboro

- Mastewal Yiseni Yimer

- Tilahun Mebre Defersha

- Tewodros Assefa Gossa

*Wolkite University Hospital*

- Dereje Zewdu*

- Tilahun Temesgen

- Khedir Fikadu

- Shimelis Getu

- Sime Assefa

- Feleke Habte

**The Gambia**

National Leaders: Mustapha Bittaye, Abubacarr Jagne

*Bundung Maternal and Child health Hospital (BMCH)*

- Dado Jabbie*

- Marie Saal Mendy

- Isatou Bobo Jallow

- Fanta Fofana

*Farafenni General*

- Lamin FTT Fadera *

- Bubacarr Jabbie

- Karimadi Fofana

- Divine Nko-Onye Moseri

*Edward Francis Small Teaching Hospital (EFSTH):*

- Sheikh Omar Bittaye*

- Fatou Jatta

- Anna Donkor

- Awa Mboob

- Abubacarr Jagne

- Fatou Jallow

*Kanifing General Hospital*

- Kajali Camara*

- Majula Ceesay

- Lamin Sanyang

- Kawsu Fatty

- Muhammed Touray

- Lamin O Beyai

- Ebrima A Jallow

- Modou Lamin Conteh

- Bubacarr Sowe

- Roheyatou Jawara

- Ndumbeh Manneh

**Ghana**

National Leaders: Christian Owoo, Daniel Sottie

*Cape Coast Teaching Hospital, Cape Coast*

- Oluwayemisi Esther Ekor*
- Emmanuel Owusu Ofori*
- Patience Koggoh
- Luke Adagrah Aniakwo
- Evans Kofi Agbeno
- Ganiyu Adebisi Rahman
- Kwasi Agyen-Mensah
- Thomas Agyen
- Martin Tangnaa Morna
- Michael Nortey
- Makafui Yigah
- Ansumana Bockarie
- Temabore Victoria Daboner
- Yvonne Ayerki Nartey
- Abigail Serwaa Boateng
- Prince Arthur
- Quainoo Arnold Kofi
- Papa Kojo Mbroh
- Edwina Okaikai Obodai

*Dunkwa Municipal Hospital (DMH)*

- John Benjamin Annan*
- Kwaku Nti Acquah
- Kwadwo Amoah
- Gloria Ababio
- David Sackey
- Emmanuel Tagoe
- Jonathan Kinphul

*Eastern Regional Hospital, Koforidua (ERHK)*

- Arko Akoto Ampaw*
- Foster Amponsah-Manu
- Philip Lartey
- Gabriel Attipoe-Djagmah
- Ijeoma Aja
- Francis Wuobar
- Audrey Addy
- Kwesi Edwin-Tenkorang
- Ransford Aduah
- Denutsui Etornam Rhoda
- Ekuamoah Violet
- Esther Adjei-Acquah
- Agnes Avorwulanu
- Ruth Dzifa Ntumi
- Charles Edem Tsivanyo
- Tetteh A. Kwame
- Pinto Henry Kabukie
- Andrea Asante

*Effiduase Government Hospital (EfGH)*

- Kingsley Damilare Adeoye*
- David Antwi-Agyei
- David Abass
- Mercy Asare
- Kwame Danquah Diaw

*Ejisu Government Hospital (EjGH)*

- Isaac Asenso Brobbey*
- Rose Mensah
- Dominic Awuah
- Afia Kyere

*Ga East Municipal Hospital (GEMH), Accra*

- Ebenezer Oduro-Mensah*
- Roxana Segborwotso
- Marion Amo-Mensah
- Reuben Grumah
- Alberta Twumasi-Boakye
- Aubrey Tigwii
- Selywn Agyemang
- Mercy Akoto
- Agnes Amonoo-Mends
- Martha Dsane-Lamptey

*Greater Accra Regional Hospital (GARH), Accra*

- Ambe Obbeng*
- Henrietta Nana Serwaa Fiscian
- Simeon Biney
- Dorcas Osei-Poku
- Nana Ama Christian
- Frederickson Pobee
- Dorinda Norbetha Lee
- Magdalene Boamah
- Ayishetu Ndeogo
- Mamm-Mavies Tsotsoo Tetteh
- Adu-Brempong Carl
- Zelda Robertson
- Linda Addai
- Gideon Appiah-Boadu
- Al-Hassan Dasana Andani
- Salamatu Nantongma
- Joseph Oliver-Commey
- Lawrence Ofori-Boadu

*Komfo Anokye Teaching Hospital (KATH), Kumasi*

- Irene Bandoh*
- Akwasi Antwi-Kusi
- Moses Siaw-Frimpong
- Eunice Aboagye
- Sophia Naa Ayikailey Ankrah
- Stephen Opoku Ofori
- Pearl Nyarko Korang
- Samuel Nana Prempeh Agyeman-Gyebi
- John Adabie Appiah
- Yaw Ampem Amoako
- Felix Owusu Osae
- Stephen Sarfo
- Kwaku Gyasi Danso
- Oppong Philip Peprah
- Martha Poku
- Maame Temah Appiah-Berko
- Ebenezer Akomea Agyin
- Anita Eseenam Agbeko
- Paa Ekow Hoyte-Williams
- Emmanuel Osei Kankam
- Joseph Bonney
- Shiela Afua Kedze
- Dennis Dwomoh Nkrumah
- Eva Adu-Boakye
- Yaw Larbi
- Kwabena Addow Opare-Addo
- Amanda Badjo Amankwata

*Korle-Bu Teaching Hospital (KBTH), Accra*

- Christian Owoo*
- Kwadwo Opoku-Darko*
- Daniel A. Sottie
- Pokua Sarpong
- Grace-Imelda Obeng-Adjei
- Lorraine Baffour-Awuah
- Ernest Aniteye
- Desmond Seshie
- Simitsewa Amoo-Aidoo
- Adjoa Ofei
- Supriya Wassiamal
- Anita Ohenewa Yawson
- Nancy Asiedua Larbi
- Kwame Afriyie Gyamera
- Kelvin Asamoah-Agyepong
- Kwaku Obeng
- Naa Martekuor Vanderpuye
- Cornelia Quarcoopome
- Khadijah Bandawu
- Aisha Sumaila
- Nuhaila Alhassan
- Lydia Apraku-Peprah
- Leslie I. Adam-Zakariah
- Kwadwo Darko
- Bless-Michael Bonney
- Raphaela Agyarko
- Felicia Birch Freeman
- Henry Kwesi Bulley
- Nana Serwaa Agyeman Quao
- Steve Blankson
- Aba Yorke
- Priscilla Allotey
- Theophyllus Addo
- Gifty Addo
- Theodore Boafor
- Maryanne Zuolo
- Kofi Adzi Gudugbe
- Henry Kumi
- Ken Atobrah-Apraku
- Eric Tetteh
- Efua Thompson
- Jerry Coleman
- William Klah
- Michael Ntumy
- Kwaku Asah-Opoku
- Gordon Amoh
- Nicholas Siklere
- Precious Owoo
- Eunice Nyankah
- Faisal Adjei
- Elvis Ohemeng-Mensah
- Jefferson Owusu-Adae
- Prince Larvie
- Clarence Abeareba Basogloyele
- Latif Saiba
- Dela Dinku
- Ernest Yorke
- Yaw Ofori-Adjei
- Jane Afriyie-Mensah
- Fiifi Duodu
- Beatrice Baaye
- Woedem Tettey
- Akosua Agyen Frimpong
- Vishnu Abayateye
- Kwame Darko
- Michael Kwapong-Nyarko
- Solomon Atindama
- Joachim Amoako
- Kenneth Baidoo
- Estella Bilson-Amoah
- Eugene Owusu-Achaw
- Evans Sefa Asiedu
- Daasebre Ahensan
- Kow Entua Mensah
- Innocent Adzamli
- Charles Kofi Amoah
- Desrie Gyan
- Matthew Owusu Boamah
- Paa Kwesi Blankson
- Prudence Nutsuklo
- Daniel Baddoo
- Isaac Asiedu
- Dela Fiagbe
- Esinu Akosua Agbeli
- Kingsley Abankwa
- Patrick Bankah
- Omane Acheamfour Okrah
- James Evans Mensah
- George Darko Brown
- Doreen Anderson
- Elikem Ametepe
- Geoffrey Birikorang
- Priscilla Vandyke
- George Nketiah
- Alfred Seedah
- Adejoke Aiyenigba
- Barbara Boi
- Serwah Amoah
- Comfort Gaituah
- Edith Ntumy
- Imoro Braimah Zeiba
- Rachel Kumi Adamson
- Nana Yaa Asuama Afful
- Emily Martha Nortey
- Freda Amoateng
- Senyo Fumador
- Priscilla Agyemang-Duah
- Esther Brobbey
- Samuel Antwi Oppong
- Alfred Edwin Yawson

*Kumasi South Hospital*

- Kwame Ofori Boadu*
- Rita Larsen-Reindorf*
- Emmanuel Sam Kwabena Baffoe
- Fredrick Gyamfi Apraku
- Nana Takyiaw Borki Owusu
- Margie Kyei
- Yaw Akyina

*Kumawu Government Polyclinic*

- Alex Agbanu*
- Joshua Dennis Krah
- Osei Amponsah-Kwatiah

*Manhyia District Hospital*

- Kwame Danso*
- Kamarudeen Korku Hussein
- Appiah Minka
- Rosemond Boah
- Louis Osei
- Hajara Alhassan
- Sarah Dwomoh
- Ernestina Sarfo
- Rita Amponsah

*Maternal and Child Health Hospital, Kumasi*

- Prosper Kwaku Gbekor*
- Laila Adutwum
- Yaa Bema
- Mabel Osei-Tutu

*Nkawie Toase Government Hospital (NTGH)*

- Eric Kwame Detoh*
- Seth Obiri
- Michael Gyamfi
- Paul Afriyie
- Seth Asante Egyin

*SDA Hospital, Kwadaso*

- Randolph Baah Adu*
- Victor Vandel Adjadeh
- Albert Amissah Asiedu

*Swedru Municipal Hospital (SMH), Swedru*

- Julius Abuku*
- Joycelyn Maame Esi Darkwah
- Richard Dankwah
- Edmund Oko Codjoe
- Dorcas Rockson
- Bernard Nyamekye Antwi
- Gifty Obo-Ninsin Baidoo

*Trauma and Specialist Hospital, Winneba (TSHW)*

- George Kwame Prah*

- Petrina Bingab
- Emmanuel Koomson
- Emmanuel Ghartey
- Frederick Duah Yao
- Panyin Benyimah Avemee
- Anyemedu Asare Fredovich
- Isaac Korku Akotoye
- Josephine Okine
- Agnes A. Anane
- Akosua Owusu Sarpong
- Marion Okoh-Owusu

*University of Ghana Medical Centre (UGMC), Accra*

- Amoako Duah*
- Kwame Ekremet*
- Nana Akosua Oppong-Nkrumah
- Joseph Tanlongo
- Dela Andrea Nutsugah
- Kordai Mould
- Susan Siabi
- Kwame Anim-Boamah

**Libya**

National Leader: Muhammed Elhadi

*Abosleem Tripoli Hospital*

- Mohamed Fathi Khmera*

- Aya Rasem Khmir

- Abdulrahman Mohammed Alghziwi

*Abusitta Hospital for Respiratory Diseases*

- Khayri Karban*

- Areej Dakshi

*Al-Afia Clinic (AAC)*

- Mohamed Saleh Addalla*

- Taha Khaled Elfaituri

- Taha Alkabat

- Ahmed Muhsin Alatiweel

- Mohammed Salaheddin Dabaie

- Abdulhamid Shaban

*Aldiaa Hospital*

- Ali Abdulnasir Kredan*

- Abdulalim Ramadan Kuridan

- Abdurrahman Abdussalam Haddud

*Alfardous Clinic*

- Ahmed Eldeeb*

- Ahmed Jreibi

*Alfath Medical Center*

- Nafati Taher Alnafati*

- Nabeel Ateeyah Faraj Atiyah

*Aljala Hospital*

- Mohamed Naser Lawgali*

- Aml Emran Khalleefah Alwirfili

- Mustafa Khalil Abdallah Elrgeig

- Rawan Othman

- Fatma Ali IKday

*Aljalaa Maternity Hospital (AMH)*

- Marwa I M Shoukrie*

- Msara Jamal Haider

- Majdi Ehmeda S Kamil

- Ameerah Ali Hassan Rahoumah

*Al-Khoms Teaching Hospital*

- Kamla Ali*

- Honayda Almuakkif

- Shahed Alaref

- Aya Haddad

- Amal Nasser

- Maram Abdulgani

*Almarj Teaching Hospital*

- Alsnosy Abdullah Khalefa Mohammed*

- Osama Alemenefie

*Alshahid Attia Alkasah General Hospital*

- Ahlaam M. Ali Ayad*

*Alshaimaa Clinic*

- Ahmed Elfaituri*

- Anwar Salah Hussain Mohamed

- Reem Abdullah Salim Alkikli

- Alaedden Abdalla Akhmag

- Saleh Abdulla Hadia

*Al-Zawia Teaching Hospital*

- Salim Almaqtouf*

- Mohamed Alsori Alharari

- Ali Alsouri Alharari

- Rawia Adel Mohamed Draa

- Abdulmunem Mustafa Olu

- Marwa Almabrouk Alazomi

- Randah Dhu Omar Aldeeb

*Benghazi Medical Center*

- Arwi Kara*

- Ayoub Akwaisah

- Mohamed Al Gharyani

- Mostafa El Awami

- Aya Essa

- Abtisam Alharam

- Aeshah Aboukaleesh

- Fatimah Aboukaleesh

- Noura Kareem

- Narjis Husayn

- Mohamed Adel

- Fatimah Mohammed Alenani

- Iesra Eldagheili

- Abdul-muhaymin Almuquryaf

- Rabab Alkurghali

- Sara Alsaeiti

- Hadeil Abd Elaziz

- Salma Akhlaif

- Fawzia Alferjani

- Wijdan Sayfulnasr

- Dalal Salah

- Muna Denini

- Ahmed Ahmayda

- Saja Almugla

- Salwa Ali Awidat

- Abdulsalam Albadri

- Fatima Elfeituri

- Arwi Adrees

- Mohammed Abosedra

- Ameerah Abraheem

- Fathiah Elferjani

- Malak Barghathi

- Yasmin Elmgawob

- Amani Zoubi

- Nourfan Altarhouni

- Asia Bolifa

- Malak Areibe

- Ehab Othman

- Suhaib Issa

- Tasnim Hasan

- Salem Senussi

- Marwah Aleidhah

- Fadelalla Elmozoghi

- Nisren Alsalme

- Fatma Elashhab

- Almotasem Bellah Elsharif

- Abdulmuez Abdulmalik

- Rana Shembesh

- Eman Bureziza

- Sara Abdelmaged

- Hana Faraj

- Najla Alaguri

- Mohammed Salem

- Mohamed Alamrony

- Mustafa Amraja

- Mohammed Elferjani

- Mohammed Alabeedi

*Brak General Hospital*

- Mohammad Yahmad

- Mubarka Alzarouq

*Crown Healthcare Clinical (CHC)*

- Hamida El Magrahi*

- Abir Ben Ashur

- Salem Ali

*Gharyan Central Hospital (GCH)*

- Hibah Bileid Bakeer*

- Akram Alkaseek

- Haitam Shames

- Aya Alqaarh

- Hashim Aborkhis

- Widad Nouralddeen Faraj Amhimmid

- Hala Misbah Ali

- Taha Husayn Alhadi Alfeeras

- Sundes Daba

- Ahmed Abdurrahman Algeblawi

*Misrata Medical Center*

- Abdulwahab Alzeldeen Abdalei

- Fatima Asedeq Abdulali

- Ahmed Ismael Saleh

- Abdulhamid Mohamed Alailesh

- Mohamed Moftah Assalhi

- Mohanad Taher Bintaher

- Boshra Basher Hashim

- Weam Mohammed Drah

- Abdelaziz Mahjub Gobbi

- Eman Mohammed Abdalhafit Alabani

- Hamdan Bashir Hilan

- Omar Mohamed Ertaiba

- Ahmed Muftah Taweel

*National Heart Centre, Benghazi*

- Wesal Tarik Yahya Hamad *

- Hadia Mohamed Omar Aldilfaq

- Aya Adam Hassan Mousa

*Preventative Medicine Hospital*

- Abdurraouf Abusalama*

- Kusay Ayad

- Abdurrahim Elzoubi

- Mohammed Altarabulsi

- Alabas Almigheerbi

- Ahmad Alfayad

- Hatim Elgbaili

- Abdulmuhaymen Elmeshrgie

- Mohammed Albashri

- Mouna Ahmed A Abuhshaima

*Sabha Medical Center*

- Khadeja Mohammed Mohammed Alawal

- Essraa Ali Alhudhiry

- Anwaar Ali Othman Eshnaf

- Safia Adem Abdulla

- Firdous Assadeq Mllie

- Mona Abdu Alsalam

- Etidal Ali Abuanniran

- Bushra Qezo

*Sabratah Teaching Hospital*

- Sara Egreara*

- Nabila Abdalkader

- Huda Abdelmajed

- Hajer Abdulhamid

- Ghazala Abouklaish

- Ahmed Alajeeli

- Khawla Alalage

- Ibtehaj Albaewi

- Malak Albusayifi

- Azhaar Aldeeb

- Ali Algarradhi

- Najwa Alkowash

- Safa Almaakef

- Mohammed Almahjoubi

- Hana Altam

- Masarah Ateeyah

- Sahar Azzaz

- Hajir Bahroun

- Doha Blaou

- Safa Barayik

- Bilqays Habhab

- Mohamed Hassan

- Saja Khalifa

- Aya Maiw

- Yasmeen Meelad

- Safiya Mohamed

- Ali Omar

- Ali Tirihbat

*Tabarak Private Hospital*

*-* Saifaleslam Elsahli*

- Abdulwahhab Fakroun

*Tobruk Medical Center*

- Manal A. Mohammed Zaglam

- Areej Alhassan Abdullah Benghuzi

- Shaymaa Moftah Salem

- Sari Sulayman Abdulhafith

- Hana Adrees Hasan

- Najwa Ibrahim

- Mohammed M. M. Abdaljalil

- Eman Moftah Faraj

- Amani Mousay Abdulmawlay Mohammed

- Malak Mashery

- Eman Ahmed Moftah

- Halima Soliman Abdualnabi

- Eman Gaith Ahwaia

- Fatama Mohamed Salem Selaman

- Esra Abdulaziz Hamad Albarani

- Dua Aljali

- Aminah Eid Abd Alsameea

- Morad G Rahel

- Murajia Mahmoud

- Aisha Mabrouk

- Marzouga Mabrouk

- Rehab Rejab

- Ayyah Emran

*Tripoli Central Hospital*

- Anas Mohammed Aboutartour*

- Batool Ahmed Abdulkarim

- Buthuynah Mohammed Alsaeh Alqahwash

- Laila Ramadan Askar

- Nada Ali Omran Dhem

- Amer Mohamed Abdulaziz Mosbah

- Khalid Asad Bin Qanad

- Abdurraouf Musbah Khalifa Said

- Qabs Madi

- Amal Ibrahim Furjane Khalifa

- Mohamed Hamed Said

- Abdulrhim Omar Mohammed Almeshri

- Marwa Muftah Daloub

- Yara Talal Mohammed Bariun

- Ahmed Nureddin Ben Shaban

- Dareen Ahmed Elmahdi

- Dawoud Amhimmid Saeid

- Firas Nureddin Hammas

- Mahad Mahmoud Qaeim

- Feras Mohamed Shneib

- Nouri Khalefah Afheej

*Tripoli University Hospital*

- Muad Fathi Abuhallalah*

- Abdalla Mustafa Abdalla Hdidan

- Abdulmalik Ibrikat

- Aisha Abdulfatah Gehimi

- Aisha Alwarfally

- Randah Dhu Omar Aldeeb

- Mawada Mohammed Elgeriane

- Rabeeah Abd Aldaem AbuAlneeran

- Abdulhamid Mohammed Alsagheer

- Safa Zaydan Ammar

- Arwa Tawfeeq Abdulnabi

- Bassam Erhoma

- Mohamed Mustafa Elghazal

- Mohamed Abdusalam Elhaderi

- Mohammmed Yousef Elimselati

- Esra Abdulhafed Abuaen

- Esra Ben Zahra

- Abduladim Omran Ezaddin

- Hidaya Badri Dozan

- Wadad Mohammed Jomaa

- Lobna Shawesh

- Mosab Emhemmed Matous

- Ghaida Abdalla Naana

- Nada Abdulmonam Elhoush

- Ola Abdulqadir Alsharif

- Rawia Adel Mohamed Draa

- Ritaj Mahmoud Ghasem Agha

- Reyam Abdalla Naana

- Abdulsabur Mohamed Salih

- Sanabil Mansour Abdullah

- Serag Lameen Almzainy

- Tasneem Musbah Fara

*Zliten Medical Center (ZMC)*

- Najat Ben Hasan*

- Abdulmalik Abeed

- Hajer Abusnina

- Majdolin Miloud Almahjoub

- Radhwan Alsaedi

- Lubnah Alsunousi Mohammed Alokshi

- Manal Almaqrahi

- Hudi Dalaf

*Zwara Maritime Hospital*

- Fahed Gareb*

- Sehar Naji Ashini

- Mwada Tarek Mehdi

**Lesotho**

National Leader: Mpho Seleke

*Mafeteng Government Hospital*

- Malefane Moysamai*

- Mpata Seleso

- Mpine Maqolo

*Paray Mission Hospital*

- Maphiri Ramafikeng*

- Tina Ntee Ntsane

- Thato Molahlehi

- Amohelang Moreki

- Lerato Mohlalisi

- Molahlehi Makhalanyane

- Noi Khoeli

- Lebohang Lebina

**Morocco**

National Leader: Ahmed Rhassane El Adib, Meryem Essafti

*Hopital Arrazi, CHU Mohammed VI*

- Samia Errami*

- Soulaimane Laaziri

- Ayoub Bousselham

- Iltimass Gouazar

- Mohamed-Sami Melouane

- Hind Essalim

- Rim Belarbi

- Rihab Belfouzy

- Hasna Laghmami

- Oumama Boujidi

- Salma Amahmid

- Riyad Ghailan

- Said Errahouy

- Adam Mohamed Aajly

- Oussama Ennamra

- Nouha Smily

- Raouia Fares

- Abdellah Agnaou

**Mozambique**

National Leader: Dino Lopes, Atílio Morais

*Hospital Central de Maputo (HCM)*

- Dino Lopes*

- Elka Nhaduco

- Francisco Taimo

- Nadia Estafeira

- Onésia Lucia Sérgio Chitsembe Mombassa

- Sebastião Moisés

- Mouzinho Saide

- Cesaltina Lorenzoni

- Luis Gonçalves Ferrão

**Namibia**

National Leader: Kaveto Sikuvi, Unotjari Kauta, Gaudencia Dausab

*Intermediate Katutura Hospital*

- Gaudencia Dausab

- Nasjtasha Pieterse

- Anna Hangula

- Fhenny Moongo

- Natasha Nghitukwa

- Rejoice Makongwa

- Dhruvin Das

- Njohela Mwandemele

- Christian Ndambi

- Larisa Jafta

- Hilma Uugwanga

- Maria Sibolile

- Seuna Karuaihe

*Katima Mulilo State Hospital*

- Bitoma Thotho Amisi*

- Munikasu Christopher

- Sinvula Lutombi
 - Masongo Annie
 - Matomola Chuma
 - Chipman Kamilla
 - Nzwile Alphoniso
 - Sisamu Mwiza Daphine
 - Kawana Concilia
 - Estha Masasa
 - Lilungwe Sonnety
 - Njahi Kaiba

*Keetmanshoop District Hospital*

- Evans Sanga*

- Rebecka Martin

- Hilma Namuhuya

- Vute Nekwaya

- Loide Johannes

*St Mary's Hospital*

- Goodman Uushona*

- Daylight Manyere

- Elina Muulu

- Lindah Kabende

- Joseph Chinanga

*Walvis Bay District Hospital*

- Chloe Paulse*

- Natasha !Gontes

- Hambeleleni Kamati

*Windhoek Central Hospital*

- Frans Nambinga

- Loide Namwandi

- Nasjtasha Pieterse

- Anna Hangula

- Fhenny Moongo

- Natasha Nghitukwa

- Faith Dzenga

- Dhruvin Das

- Njohela Mwandemele

- Christian Ndambi

- Ndapunikwa Nghihalwa

- Justina Shikongo

- Tracy Mweti

- Larisa Jafta

- Hilma Uugwanga

- Melissa Spiegel

- Maria Sibolile

- Seuna Karuaihe

**Nigeria**

National Leader: Adesoji O. Ademuyiwa, Babatunde Osinaike.

*Abubakar Tafawa Balewa University Teaching Hospital*

- Ibrahim Salim Abdullahi*

- Auwal Adamu

- Rabiu Mohammed Bashir

- Abubakar Muhammad Ballah

- Isa Bashiru Ibrahim

- Yusuf Raiyanu Jasawa

- Aliyu Bashir Adamu

*Ahmadu Bello University Teaching Hospital*

- Saidu Yusuf Yakubu

- Bilkisu Adamu

- Oluseyi Oyebode Ogunsua

- Shafaatu Ismail Sada

- Tunde Talib Sholadoye

- Alfa Yakubu

- Halima Olufunmilola Abdulsalam

- Fomete Benjamin

- Samuel Isa Gana

- Abdulkadir Muhammad Kabiru

- Hamisu Yakubu

- Rabiu Isah Mohammed

- Sufyan Ibrahim

- Umma Suleiman Bawa

- Ganiyat Ronke Olagunju

- Babangida Salahu Mohammed

- Mohammad EL-Amin Idris

- Michael Odigbo

- Fatima Mahmud-Ajeigbe

- Aghadi Ifeanyi Kene

- Tasiu Saadu

- Sheidu Owuda Abdullahi

- Rabi'at Muhammad Aliyu

- Mudi Awaisu

- Chitumu Dotiro

- Stephen G. Gana

- Lucy. E. Okwajebi

- Muhammad Daniyan

- Abdulghaffar Adeniyi Yunus

- Olutayo A. Gana

- Oguntayo Olanrewaju Adekunle

- Muhammad Lawal Abubakar

- Ibrahim Ibrahim Lawal

- Atiku Likunga Atiku

- Ukwubile Linus

- Ahmad Bello

- Ahmad Tijjani Lawal

- Aminu Muhammad Balarabe

- Anisah Yahya

- Shehu Toro Muhammad

- Emmanuel Raphael Abah

- Fakuta Tathiya Naiwa

- Hussaini Yusuf Maitama

- Nwoye Ugo Daniel

- Elizabeth Ogboli-Nwasor

- Abdullahi Sudi

- Muhammad Raji Mahmud

- Ugwu Euphemia Mgbosoro

- Igele Agom Cletus

- Abass Oluwaseyi Ajayi

- Emmanuel Paktama Bwala

- George Duke Mukoro

- Habibu Balarabe

- Sani A. Abubakar

- Bilqis O. Muhammad

- Ademola O. Adeleye

- Sunday O Ajike

*Alex Ekwueme Federal University Teaching Hospital*

- Donatus O. Egwu*

- Richard L. Ewah

- Promise O. Ubanatu

- Cletus U. Onwe

- Nathan O. Mbawike

- Joshua A. Adebayo

- Chijioke Udu

- Nneka N. Chiege

- Stephen O. Nwafor

- Stephen C. Eke

- Pearl C. Eke

- Chinyere S. Onoka

- Juliana Chi-Nwogo

- Mary Okoyari

- Oluchi F. Ogah

- Chinenye T. Agbo

- Obiageli A Obi

- Mercy N. Nwangwu

- Loveth N. Ugochukwu

- Kelechi Adindu

- Stella A Ugochukwu

- Obiora C. Bernard

- Sunday V. Nwali

*Aminu Kano Teaching Hospital*

- Abubakar Bala Muhammad*
- Lofty-John Anyanwu
- Datti Alhassan Muhammad
- Salahu Dalhat
- Bashir Yunusa
- Mustapha Ibrahim Usman
- Abdulrahman Muhammad
- Abdulrazak Ajiya
- Ademola Babatunde
- Ramalan Mansur  Aliyu
- Aisha Nalado
- Alfa Mika'il Abdullahi
- Aliyu Ibrahim
- Amal Galadanci
- Aminu Abba
- Mamuda Atiku
- Atiku Jibrilla
- Bello Muideen Abodunde
- Hamza Muhammad
- Ibrahim Musa Idris
- Isma'il Jibrin
- Baba Ahmad
- Kabir Adamu Musa
- Shehu Kana
- Mahmoud Kawu Magashi
- Mohammad Aminu Mohammad
- Musa Zango
- Mustapha Miko Abdullahi
- Muzzammil Abdullahi
- Usman Abubakar Nagoma
- Nasiru Ishaq
- Oseni Ganiyu
- Saminu Muhammad
- Shamsudeen Muhammad
- Sulaiman Daneji
- Idris Usman Takai
- Zynat Sani Alhassan
- Tijjani Nasiru Nagwamutse
- Misbahu Haruna
- Abdulrahman Abba Sheshe
- Garba Ilyasu
- Musa Babashani
- Suleiman Abdulrashid
- Musa Baba Maiyaki

*Babcock University Teaching Hospital*

- Omotayo Felicia Salami*

- Clifford Imonitie

- Usman Kolawole Ajayi

- Ayodeji Emmanuel Babalola

*Federal Teaching Hospital Iddo-Ekiti*

*-* Abiodun Idowu Okunlola*

- Cecilia Kehinde Okunlola

- Segun Alex Atolani

- Olumuyiwa Ariyo

- Tesleem Olayinka Orewole

- Olakunle Fatai Babalola

- Adedayo Idris Alawu

- Paul Olukayode Abiola

- Omagbeitse Henry Abiyere

- Augustine Adebayo Adeniyi

- Adewumi Bakare

- Ajayi Adeleke Ibijola

*Federal Teaching Hospital Katsina*

- Ibrahim Salisu*

- Auwal Mohammed Abdullahi

- Naziru Garba Shuaibu

- Ibrahim O. Shuaibu

- Akeem Ibiyemi

- Jafar Halliru

- Sanusi Bala

- Sani Kasim Inuwa

- Gamaraddeen Abdullahi Muhammed

- Auwal Haruna

- Amina Umar Akeel

- Yahaya Lawal Kankara

*Irrua Specialist Teaching Hospital*

- Irene Irenosen Akhideno*

- Kelvin Salami

- Peter Itua

- Patrick Iniaghe

- Rachael Izekor

- Christian Uanzekin

- Ehizojie Fidelis

*Jos University Teaching Hospital*

- Samuel Isaiah Nuhu*

- Henry Yammoh Embu

- Mangai Audu Ngeh

- Kefas Thomas Malau

- Musa Abdullahi Aliyu

- Husseina Amina Aliyu

- Udoka Okorie

*Lagos State University Teaching Hospital*

- Oluwaseun David Oladokun*

- Obashina Ayodele Ogunbiyi

- Oluwayemisi Bamidele Oluwadun

- Oluwafunmilayo Aderemi Ikotun

- Akintayo Olugbemi Ogunjuboun

- Yetunde Adebimpe Oyeyode

- Nasiru Akobe Suleiman

- Folayinka Ayofunke Ogunmuyiwa

- Ilochi Uchechukwu Nnaji

- Ibrahim Olajide Dada

- Fatima Ajuma Ojabo

*Lagos University Teaching Hospital*

- Muyiwa Rotimi*

- Adesoji Ademuyiwa

- Damilola Awotunde

- Chisom Agwu

- Oluwajuwon Afolayan

- Jennifer Okei

- Abidat Ashimi

- Oluwadarasimi Adeboyeku

- Abimbola Ogunbadejo

- Daniel Lawal

- Shakirat Adejumo

- Goodness Donye

- Tobiloba Lawal

- Gideon Osadare

- Titilola Awosika

- Jared Oseghale

- Danielle Obiwulu

- Christianah Otegbola

- Emmanuel Williams

*Memfys Hospital*

- Maduabuchi Paul Ufoegbunam*

- Chika A. Ndubuisi

- Friday G Okonna

- Obioma Richards Akwada

- Donald E. Ogolo

*Olabisi Onabanjo University Teaching Hospital, Sagamu*

- Fatungase Oluwabunmi Motunrayo*

- Shoyemi Ramotalai Oluwatoyin

- Shotayo Oluwakemi Adenike

- Adefuye Bolanle Olufunlola.

- Ogunjimi Luqman Opeoluwa

- Nwokoro Chigbundu Collins

- Ogundele Ibukunolu Olufemi

- Amosun Lukmon

*University College Hospital, Ibadan (UCHI)*

- Olusola Kayode Idowu*

- Babatunde B Osinaike

- Afolabi Adebayo Oladeji

- Oluwaseun Kehinde Adebayo

- Mutiu A Jimoh

- David A Aderinto

- Yemi Raheem Raji

- I Adeola Fowotade

- Ajibola Oladiran

- Oluwakemi Badejo

- Kehinde Abraham Ojifinni

- Adigun Tonitomi

- Mosimabale J. Balogun

- Taiwo A Lawal

- Oyeyemi E. Dada

- Yewande Olaoye Babalola

- Oluwasanmi Adekunle Ajagbe

- Olurotimi Olaolu Akinola

- Oluseun O. Saanu

- Olatunji Okikiola Lawal

- Olayinka Ramotu Eyelade

- Adegbolahan Jacob Fakoya

- Olukemi A Adekanmbi

- Mukaila O Akinwale

- Arinola Adeyoola Sanusi

- Foluke Oladele Sarimiye

- James Ayokunle Balogun

- Matias Ogbonia Orji

- Afieharo I. Michael

- Rukiyat Adeola Abdus-Salam

- Dare Isaac Olulana

- Omobolaji Oladayo Ayandipo

- Oludolapo Olawunmi Afuwape

- Sikiru Adekola Adebayo

- Tarela Frederick Sarimiye

- Oluseyi O. Agboola

- Thankgod C Okonkwo

- Augustine Oghenewveyin Takure

- Adekunle Daniel

- Douglas Efe

*University Of Benin Teaching Hospital (UBTH)*

- Akpabio Uforo Ezekiel

- Imuetinyan Rashida Edeki

- Aluya Eseosa Faith

- Ehiorobo Samson Edohen

- Omorogiuwa Idemudia Oduware

- Aighobahi George Akpede

- Peter Ikponmwosa Agbonrofo

- Michael Ediale

- Joel Enaholo

- Otasowie Osagie

- Oduware Emmanuel Ehigiegba

- Noruwa Patience Ekhator

- Omorogbe Scott Osahon

- Osaheni Osayomwanbo*

*University Of Calabar Teaching Hospital (UCTH)*

- Stella A. Eguma

- Sunday O. Sangolade

- Chinedu J. Anachunam

- Nkoyo E. Enyenihi

- Victoria E. Ndoma

- Augustine O. Odemwingie

*University Of Ilorin Teaching Hospital*

- Olanrewaju Olubukola Oyedepo

- Benjamin Olusomi Bolaji

- Christianah Iyabo Oyewopo

- Abdulrasheed Adegoke Nasir

- Olayide Sulaiman Agodirin

- Lookman Oluwatosin. Lawal

- Anne Oluwabunmi Mokuolu

*University Of Maiduguri Teaching Hospital, Maiduguri (UMTH)*

- Audu Idrisa*

- Ahmed Hamman Gabdo

- Nuhu Ali

- Jamila Audu Idrisa

- Mohammed A Ahmed

- Mohammed A S Abdullahi

- Ali Mohammed Ramat

- Ibrahim Musa Kida

- Babagana Bako

- Hassan M Dogo

- Babagana Usman

- Jibril Khalil

- Hamman Ibrahim Garandawa

- Sulaiman M Maina

- Olayinka Adewunmi

- Maryam Usman Kashim

*University Of Port Harcourt Teaching Hospital (UPHTH)*

- Job Otokwala*

- Vernatious Aniobi

- Busola Alagbe-Briggs

- John Udo

- Ageh Hannah

**Somalia**

National Leader: Mohamed Abdinor Omar

*Madina Hospital*

- Hilal Mohamed Nor*

- Abdishakor Mohamud Ahmed

- Mohamed Abdi Ahmed

*Mogadishu Somalia - Turkey Recep Tayyıp Erdoğan Training and Research Hospital (TRTETRH)*

- Abdullahi Said Hashi*

- Mohamed Sheikh Hassan

- Mohamed Farah Yusuf Mohamud

- Nasra Mohamud Hilowle

- Marian Muse Osman

*Kalkaal Hospital*

- Sakariye Abdullahi Hassan*

- Suleyman Abdullahi Mohamed

- Timothy Kimutai

- Abdullahi Mohamed Mohamud

**Somaliland**

National Leader: Hassan Ali Daoud, Mohamed Faisal

*Borama Regional Hospital*

- Mouna Ahmed Abdillahi*

- Fatima Eid Ibrahim

*Burao General Hospital*

- Omar Aden Yusuf*

- Mohamed Said Awil

- Hamse Awil Mohamoud

*Hargeisa Group Hospital*

- Ayoub Mohamed Suleiman

- Mohamoud Abdullahi Ismail*

- Samira Maxamed Macalin

- Abdirahman Abdirisak Ahmed

**South Africa**

National Leader: Juan Scribante

Provincial Leaders: Busiswe Mrara, Sade Hendricks, Nondwe Mgoqo, Jenny Nash, Fathima Paruk, Zainub Jooma, Zane Farina, Marie Oosthuizen, Estie Cloete

*Addington Hospital*

- Sebenzile Sikhakhane*

- Usha Singh

- Jayd Kanjee

- Nishen Gokal

- Nqobile Zulu

- Sabihah Murchie

- Mausum Beeput

- Nokukhanya Shange

- Sarisha Haripersad

- Leona Ravinath

- Kerissa Naidoo

- Thabang Kolanyane

- Njabulo Ntuli

- Lusanda Magwenyane

- Fezile Mkhize

- Thubelihle Nsele

- Lethuxolo Shange

*Adelaide Hospital*

- Angela Mary Thain Hartwig*

- Relebohile Joyce Khoabane

- Mfundo Gubhela

*Butterworth Hospital*

- Bongeka Mfecane*

- Musa Sarile

- Thobile Dlalisa

- Nashlen Naidoo

*Charles Johnson Memorial Hospital*

- Shepherd Nzenza*

- Jacob Myeni

- Kwanele P Majozi

- Mlungisi P Shange

- Zakhele Nxumalo

- Ziphokuhle Khumalo

- Mujinga Sylvie Biaya

*Church of Scotland Hospital*

- Siphelele Kubheka*

- Rodney Magwenya

- Thuba Mazibuko

- Nkanzimulo Dlamini

- Khayelihle Jobe

- Mondli Mfeka

- Lindokuhle Sangweni

- Snegugu Zondi

- Mbongeleni Zuma

- Siphesihle Njoko

- Mounir Raddadi

- Lindokuhle Zungu

*Cloete Joubert Hospital*

- Judy-Liesel Hardie*

- Marion Irene Frost

- Mandisa Sfundo Ziqubu

*Cradock Provincial Hospital*

- Elsje Bester*

- Anli van Niekerk

- Heinri Edwards

- Joy Awokiyesi

- Sinazo Ngini

- Carli Whitehead

*Dr Pixley Ka Isaka Seme Memorial Hospital*

- Tessa Korda*

- Theroshnie Kisten

- Halalisiwe Khanyi

- Shakeel Kader

- Matthew Jones

- N Buthelezi

- Chris Hooper

- Urekha Ballasur

- Susan Brown

- Alicia Ackerman

- Ahmad Asmal

- Muhammad Asmall

- Nico Hudson

- Rene Kruger

- Kedibone Mbanga

- Vuyisa Mdingi

- Molifi Moshoadiba

- Zanine Moyce

- Fezile Mthimkhulu

- Akhie Narain

- Marsha Ramburuth

- Tapiwa Sibanda

- Sivenderen Thaver

- Cherade Wilson

- Nondumiso Zondi

- Mkholisi Gama

- Genevie Borrageiro

*Eerste River Hospital*

- Martha Elizabeth Bronkhorst*

- Danielle Charlotte Hendricks

*Emmaus Hospital*

- Mampho Mochaoa*

- Yonela Tsewu

- Mbuso Nkala

- Lwanzo Mathe

- Deliwe Nkosi

*General Justice Gizenga Mpanza Regional Hospital*

- Roel Matos-Puig*

- Ernest Muragijeyesu

- Ria Devi Naidoo

- Nompumelelo Precious Sibiya

- Nthabiseng Precious Molokwane

- Shilendra Harripersad

- Dyavan Singh

- Vajra Chandrasekhar

*Grey’s Hospital*

- Arisha Ramkillawan*

- Nomandla Tsibiyane

- Nandini Gramoney

- Thubelihle Jali

- Michelle Terry Dolores Smith

- Sinovuyo Madikane

- Ayanda Ntinga

- Sinenhlanhla Mkhize

- Zukiswa Cabangana

- Sakhisosenkosi Zamimpilo Hlela

- Parvania Munthree

- Takudzwa Shava

- Lieze Geldenhuys

- Dilshad Shaik

- Jithin Mohan

- Mihir Patel

- Sabeeha Khan

- Fathima-Zahra Rahiman

- Richard Tatenda Chikosi

- Alexandra Bench

- Marcel Simpson

- Natalie Hendricks

- Freda-Heléne Leuvennink

- Ingebor Jäger

*Groote Schuur Hospital*

- Rowan Duys*

- Margot Flint

- Simphiwe Gumede

- Abhilash Nair

- Aleta Sibi

- Aliya Bhorat

- Ayabonga Yedwa

- Chloe Ash

- Danika Govender

- Darren Piaray

- Dineo Kotu

- Jacob Blou

- Jani Gerber

- Jenna Piercy

- Johann Fourie

- Joseph Ruiz von Walter

- Joshua Louis

- Khelan R Dheda

- Khulile Singata

- Mahdiya Bhayat

- Mayilan Chetty

- Nabiha Ebrahim

- Naho Khorombi

- Nina Mabusela

- Olachi Emeruem

- Phi Nguyen

- Rahul Joseph

- Robyn Brown

- Samiya Bhorat

- Suvina Chanerika

- Tiashan Moodley

- Zaeem Ebrahim

- Zander Skye Isaac

*Harry Gwana Hospital*

- Mariette Grobbelaar*

- Dave Bishop

- Marzanne Nel

- Leesa Bishop

- Sanele Magagula

- Kirsten Marais

- Victor Ruhinda

- Suhail Sayad

- Nerlicia Sewnarain

- Aphiwe Mtshengu

- Tsakani Mabunda

- Sisanda Tiya

- Dipo Adeyemi

- Muhummed Uwais Moosa

- Lwazi Goso

- Anika Lucas

- Emma Scheepbouwer

- Nabeela Motala

- Glenda Watt

- Nok’zotha Njoko

- Gaby Nel

- Bonga Mthembu

- Melissa Purdon

- Previn Johnson

- Humeshan Naidoo

- Jan Kriel

- Howard Wain

- Buhlebonke Skhakhane

- Asheeq Soobader

- Abdur Razaaq Lamera

- Melanie Lesch

- Cara Dunn

- Hilde Turkstra

- Marcello Leita

- Tajil Manmohan

- Natanael Janse van Rensburg

- Nkosi Nxumalo

- Thandiwe Mchunu

- Jisheel Kanaye

- Tamsin Singh

- Bonga Khoza

- Melissa Yolanda Hippolite

- Nonhlanhla Zimase

- Nonhlanhla Madlala

- Stephanie Scriba

*Isimelela Hospital*

- Graeme Hofmeyer*

- Richard Southey

- Michaela Peters

- Muetu Kapena

*King Edward Hospital*

- Kasandri Govender*

- Alishka Naidoo

- Fiona Lourens

- Andisiwe Ngcobo

- David Govender

- Yasheen Maharaj

- Lwazi Mntungwa

- Dhivendra Singh

- Suman Mewa Kinoo

- Ruvashni Naidoo

- Kishan Anand Naidu

- Nhlakanipho Ngubane

- Maaneka Ramadhin

- Randolph Green-Thompson

- Nyameka  Maluleke

- Karthik Naidoo

- Sne F Mkhize

- Patrick Kanana

- Brian Ubisi

- Mfundo Ndlela

- P Hiralal

- S Lakhani

- Balungile Dzingwe

- Andile Mfene

- Tumisho Bokgobelo

- Nithin Khoon Khoon

- Aadi Modi

- Chanel Dalais

- Zubair Omar

- Meegashen Narainsamy

- Nombuso Mlawu

- Saba Sivuyisiwe

- Taskeen Mather

*Ladysmith Hospital*

- Nozipho P. Mabaso-Langa*

- Brain Sibusiso Blakie Mbhele

- Sihlengiwe Zandisile Nyawose

- Jabulani Agrippa Nzimande

- Pholoso Prince Moele

- Mxolisi Brian Ndimande

- Nonhlanhla Zulu

*Madadeni Hospital*

- Marcin Kopieniak*

- Christian Kyanda-Kaboza

- Farrah Khan

- Etienne Venter

- Nondumiso Masondo

- Bonginkosi Nene

- Abiodun Lamina

- Nkosi Sibahle

- Samukelesiwe Kubheka

- Tilou Thobane

- Mzamo Mnguni

*Madwaleni Hospital*

- Andrew Miller*

- Steyn Botha

- Melandie Fourie

- Reinhard Schmidt

- Chris Westwood

*Mahatma Gandhi Memorial Hospital*

- Devandiran Harriraman Rungan*

- Devarani Naidoo

- Adushan Govender

- Zizipho Noluthando Madikizela

- Siddharth Mohan

- Bavna Hira

- Keshree Naidoo

- Valsura Ramsundar

- Brendan Moonsamy

*Manguzi Hospital*

*-* Mlibo Mthembu*

- Mark Blaylock

- Eddie Tembe

*Mthatha Regional Hospital*

- Busisiwe Cawe*

- Michael Ojonimi Ameh

- Osahon Daniel Erebor

- Sijabulile Cassius Sosibo

- Sikelela Siyibane

- Charles Choto

- Mlamli Dotye

- Zolelwa Zibuye Nandi

- Sanelisiwe Sivuyile Ngceba

- Thulisa Anela Qaziyana

- Chwayita Katshwa

- Unathi Simamkele Ntshongwana

- Sivuyise Gwazela

- Lerato Pakade

- Siphosihle Msutu

- Singleton Luxolo Sandla

- Thobeka Portia Ngcobo

- Sandiso Yekani

- Vuyolwethu Sotashe

- Khanya Galela

*Nelson Mandela Academic Hospital*

- Abongile Sukwana*

- Busisiwe Mrara	(Provincial Leader)

- Lindubuhle Beba

- Mawande Mayibenye

- Samuel Alomatu

- Siqhamo Magadla

- Gloria Hyera

- Siphesihle Khoza

- Lelethu Jwambi

- Mzomhle Kiza

- Athandiwe Qhonono

- Lungisa Petse

- Nomvuzo Saqu

- Lwandile Mtshabe

- Sibi Joseph

- Yondela Sidoyi

- Banele Kokose

- Anele Dayimani

- Chwayita Makrexeni

- Emma Muendo Loko

- Apelele Futshane

- Sinesipho Ndedwa

- Nosiphiwo Semane

- Aphiwe Mqhayi

*New Somerset Hospital*

- Jacob Blou*

- Rahul Joseph

- Joseph Ruiz von Walter

- Phi Nguyen

- Joshua Louis

- Chris Pearce

- Robin Veitch

- Samiya Bhorat

- Aliya Bhorat

- Dineo Kotu

- Margot Flint

- Nina Mabusela

*Newcastle Hospital*

- Sandile Dube*

- Makura Ndhlovu

- Siphiwe Ngema

- Kaajal Brijlall

- Thabani Nyathikazi

- Kashal Ramsaroop

- Ameer Beeharry

*Ngwelezana Hospital*

- Stefan van der Walt*

- Chiara Anne Baars

- Karishma Heeramun

- Sabrina Pillay

- Xander Botha

- Christopher Brits

- Chad Young

- Andrew Monahan

- Mikaeel Abdool

- Brendon Chetty

- Anele Thabiso Ndlangisa

- Asanda Marawu

- Phumzile Khuboni

- Philiswa Nkondlo

- Siphe Xabendlini

- Abu Bakr Arbee

- Michael Robert Cooke

- Hsin-Nua Wang

- Jeandré Malherbe

- Craig Sydney Scholtz

- Michael Santana

- Jo Shadwell

- Julia Tooke

- Paul Martin Pretorius

- Zamancwango Mncwango

- Luke Miguel Fourie

- Daniel McElhenny

- Riza Ehlers

- Andile Twele

- Julia Kenmuir

- Rory Livanos

- Gregory Wiid

- Lwande Gongota

- Anele Thabiso

- Zaza Shange

*Northdale Hospital*

- Dela Maiwald*

- Henri van der Merwe

- Reena Panicker

- Saidur Molla

- Sarel Kruger

- Sanam Chuturgoon

- Laila Choonara

- Werner Erasmus

- Aashiqui Ramith

- Alana Williams

- Anathi Sokanyile

- Dane Rampini

- Emma Howes

- Gabriella Kwant

- Heilisha Dehaloo

- Kaylin Adams

- Seelo Mvelase

- Mthokozisi Nsele

- Ridwa Hajee

- Saahil Bhanial

- Tajil Manmohan

- Sajit Alladeen

*Port Shepstone Hospital*

- Robert Stevenson*

- Preston Moodley

- Shirish Sewpersad

- Spheh Ndlovu

- Marli Ellis

*Prince Mshiyeni Memorial Hospital*

- Rajesh Ramjee*

- Myint Aung

- Prashant Gokal

- Omishka Hirachund

- Sizwe Zungu

- Allison Smith

- Wayne Rees

- Nishan Pillay

- A Hirjee

- Khaya Matanzima

- Pavan Kishendutt

*Queen Nandi Hospital*

- Ravi Mishra*

- Yoshua Bwambale

- Mphathiseni Dlamini

- Dedi Mutondo

- Mbayo Yuma

- Chane Retief

- Wandile Shandu

- Precious Mzobe

- Tando Mpotulo

- Emily du Plessis

*Robert Mangaliso Sobukwe Hospital*

- Judith Maria Oosthuizen*

- Nicquin Nervin Adams

- Andrea Snyman

- Sunnet Ellis

- Jacobus Rademan

- Lisa Combrink

- Herman Hendrik van der Linden

- Pieter Gerhardus Marais

- Matshidisho Nodoba

- Thazriq Eksteen

- Yusuf Ameen

- Sidharth Singh

- Tamilla Nieuwoudt

- Michaela Bredenkamp

- Nitesh Naranbhai

- Anke van der Linden

*St Mary's Hospital*

- Adam Asghar*

*-* Nazreen Ahmed

- Lungile Mthethwa

- Zein Elabideen Darwish

- Aavishkar Rooplall

- Kamintha Govender

- Kershan Munien

- Mohamed Nadeem Ali

- Zahraa Vahed

- Thembelihle Pretty Hloni

- Siyabonga Ximba

- Phumelele Ntinga

- Leeyen Singh

- Sunhera Sukdeo

- Jabulilie Solomon

- Nikeziwe Ncwane

- Nokwanda Mabaso

- Pumla Zondi

*Steve Biko Academic Hospital*

- Nhlakanipho Shabalala*

- Tiffany Pratt

- Mmapali Mokapela

- Chuene Hlahla

- Nkateko Chauke

- Petrus Jansen van Vuuren

- Chloe Howell

- Sanjula Pillay

- Dineo Mashigwane

- Fred Mulunda

*Uitenhage Provincial Hospital*

- Jannes Moolman*

- André Oosthuizen

- Allison Muller

- Maryll Stuurman

*Victoria Hospital*

- Zahnne Fullerton*

- Ziyaad Limalia

- Abigail Davies

- Kelsey Bester

- Margot Flint

- Simphiwe Gumede

*Wentworth Hospital*

- Mergan Naidoo*

- Dylan Barnard

- Jadon Haridutt

- Leigh Coetzee

- Shanaaz Fortune

- Deveena Jasmin Maharaj

- Muhammad Yusuf Sayed Essop

*Zithulele Hospital*

- Nicholas Fine*

**Sudan**

National Leader: Tarig Fadalla, Alshaima Koko

*Merowe Daman Hospital*

- Mazin Mahir*

- Alhussein Hamid

- Abdelfatah Abdelmageed

*Wadi-Halfa Hospital*

- Sara Abdalla*

- Shadan Elsayied

- Moaj Ahmed

- Alamin Ali

**Tanzania**

National Leader: Tim Baker, Karima Khalid, Bernard Mbwele

*Muhimbili University of Health and Allied Sciences*

- Aneth C Kaliza

- Linda Mlunde

- Elibariki Mkumbo

- Rafael S Shayo

- Godfrey Barabona

- Happines Biyengo

- Alma O Damasy

- Anna Hvarfner

- Sabra Hussein

- Anab F Issa

- Charles Machumu

*Bahi District Hospital*

- Edson Gaston Luvakubusa*

- Atupyane Mati

- Simforosa A. Atanas

- Monica N. Lucas

- Michael J. Munuo

- Zulfa S. Matola

*Chemba District Hospital*

- Greyson Victor Joseph*

- Asadulillah Kidusi

- Scholastica B. Bitesigirwe

- Dionisia A. Mkonga

*Ilsofu District Hospital Sumbawanga*

- Jenitha Joseph Cheru*

- Petronela Malema

- Shija Mathias

*Kilimanjaro Christian Medical Center*

- Maynard Samuel Makere*

- Chrystal Tarah Munyanyi

- Kelvin Fidelis Rutahoile

- Linnah Abraham Njau

- Rutendo Gina Mutimukulu

- Precious Nicole Gunha

*Mawenzi Regional Refferal Hospital*

- Leonard Joseph Mrosso*

- Serena Silayo

- Flora Mbuyu

- Joyce Materu

- Mary John Chuwa

*Mbalizi Designated Regional Hospital*

- Mathayo Thomas*

- Grace Charles

- Dina J. Lihangaka

- Herieth Mwenda

- Bebatus Nyamahanga

*Mbeya Regional Referral Hospital*

- Cholela Braison Augustino*

- Pascal Robert Dalama*

- Remigius Richard Apolinary

- Edward Mathias Nsemwa

- Magreth Gitige Mangi

- Catherine Gatwa

- Geofrey Oscary Kayombo

- Winfrida Yena Ndebele

- Keneth Mashauri Kyando

- Stella Maneno Jacob

*Mbeya Zonal Referral Hospital (MZRH)*

- Sixtus Ruyumbu Safari*
- Filbert Francis Ilaza

*Muhimbili Orthopedic Institute*

- Shafi Hamis*

- Monica Peter

- Rehema Mushi

- Sajda Ally

- Heneriko Benedicto

- Gustavu Mbunda

- Reuben Andeshi

- Abel Elias

- Willium Anstone

- Fransesco Aron

- Saguda Matondo

- Suzana Constantine

*Mwalimu Nyerere Regional Hospital*

- Ezekiel Komanya*

- Samwel Asantaeli

- Francis Joseph Gwejo

- James Gambaseni

*Mwananyamara*

- Hubert August Ngowi*

- Lucky Harran Mkocha

- Sesilia Michael

- Onesmo Seth Laizer

- Dede Dotto Chapajuja

*Rubya Catholic District Hospital*

- Arserius Rutaiwa*

- Jackline P. Kokuhirwa

- Efgenia M. Bombo

- Domina J. Muikila

- Leah D. Mwelinde

*St. Joseph Designated District Hospital*

- Deusdedith William*

- Liberatus Mushi

- Bonphace B. Msimbano

- Clement J. Mchomvu

*Sumve Designated District Hospital*

- Charles Charles Petro*

- Pendo N. Yombo

- Tekla Timotheo

- Christian L. Kusekwa

- Servat Ikigijo

- Patricia Osward

*Utete Hospital Rufiji*

- Ephrem Tahhani

- Stamili Mkumba

- Senso William

- Hadija Wanguvu

*Uwata Hospital*

- Edwin Edward Ernest*

- Edwin Edward

- Eliab J. Daud

- Innocent B. Sanga

- Anna J. Kalinga

**Tunisia**

National Leader: Nahla Kechiche

*Habib Bourguiba University Hospital*

- Rania Ammar*

- Mounir Bouaziz

- Fatma Kolsi

- Rahma Daoued

- Mouna Jerbi

- Noureddine Rekik

- Amel Bouzid

*Hedi Chaker Hospital of Sfax*

- Hayet Zitouni*

- Sahar Elleuch

- Salma Toumi

- Fatma Khanfir

- Sana Omri

- Kamel Kolsi

- Mohamed Ben Hmida

- Moamed Maalej

- Kais Chaabane

- Riadh Mhiri

*Hospital of Gabes*

- Samar Bellil*

- Houda Belmabrouk

*University Hospital of Mahdia*

- Oussama Jaoued*

- Imen Bannour*

*University Hospital of Monastir*

- Sawsen Chakroun

- Nessrine Ben Saad

- Marwen Baccar

- Lassaad Sahnoun

**Uganda**

National Leaders: Adam Hewitt-Smith, Arthur Kwizera

*Budaka HCIV*

- John Kiboma Wogabaga*

- Mercy Logose

- Yaya Miriam Jackline

*Busiu HCIV*

- Roggers Odongo *

- Aruho Moses

*Busolwe Regional Hospital*

- Innocent Musiime*

- Joan Naluyima

- Peter Magala

*Buwasa HCIV*

- Moses Tommy Wobudubire*

- Mary Nambuba

- Gerald Waniala

- Joram Esemu

- Getrude Nakiria

- Emily Chesang

- Zamu Logose

- Salim Nabila

*Hoima Regional Hospital*

- Sophie Namasopo*

- John Kalungi

- Isooba Safiyu Ayub

- Amon Asindu

- Elizabeth Namuyala

- Isaac Muyanja

- Ivan Mwebesa

*Jinja RRH*

- Ruth Muhindo*

- Anthorny Wasukira

- Mary Innocent Waswa

- Ronald Aruho

- Moses Mwebaza Kakooza

- Grace Nambuya

- Brenda Balungi

*Katakwi General Hospital*

- Joseph Emuron*

- Clare Ameri

- Roggers Kedi

- Constance Amulen

*Lubaga Hospital*

- Gladys Nabukenya*

*Mbale RRH*

- John Paul Ochieng*

- Joan Nachuka

- Priscilla Mercy Adikin

- Peace Draleru

- Naume Etoko Akello

- Federeth Nabisubi

- Hasifah Namutebi

- Glades Isina

- Jennifer Kipwola

- Derrick Muhwana

- Fred Salya

- Harriet Patricia Asekenye

- Christine Tino

- Doreck Nahurira

- Assen Kamwesigye

- Jude Mulowoza

- Ronald Aridriga

- Sam Orech

- Brenda Namugga

- Richard Gamubaka

- Fred Maiso

- Joshua Orikiriza

- Andrew Lemu

- Jena Amos Chebet

- Moses Chelangat

- Bruno Onen Chan

- Zainab Kabasemeza

*Mulago National Referral Hospital*

- Jane Nakibuuka*

- Paul Omagor

- Lynn Martha Nattabi

- Vanessa Nantale Lubulwa

- Benard Oyang

- Moses Arinaitwe

- Eddy Cantong

- Vanessa Nanono

- Dramanigo Kodjo

- Maria Assiimwe

- Bright Nagaba

- Ceasor Julius Owor

- Nicholas Kabugo

- Emmanuel Eemu

- Dickson Kamoga

- Charles Emuduko

- Sheif Semakula

- Cornelius Sendagire

*Nsambya Hospital*

- Tomanya Kakuru Kenneth*

- Susan Nabunya

- Joyce Wamala N.

- Claire Namuwaya

- Chelsea Edwards Sanyu

*St Mary’s Hospital Lacor*

- Kenneth Innocent Nyeko*

- Mwesigwa Lukwago Seezi

- Allan Phillip Barigye

- Herman Tabula Mpumbu

- Derrick Mukurasi

- Ronnie Omoro

- Maurine Lenia

*Uganda Heart Institute*

- Catherine Namutebi*

- Sarah Namatovu

**Zimbabwe**

National Leader: Pisirai Ndarukwa, Newten Handireketi

*Marondera Hospital*

- Ndaiziwei Masukhume*

- Samson Pomo

### Supplementary Material S1: Results of a systematic search of publications on the prevalence of acute pain and its relationship with critical illness and mortality.

*Search strategy*

((Adult[Title/Abstract] OR adults[Title/Abstract]) AND (Inhospital[Title/Abstract] OR hospitalised[Title/Abstract] OR hospitalized[Title/Abstract] OR inpatient[Title/Abstract] OR inpatients[Title/Abstract] OR Hospitalization[Title/Abstract] OR Hospitalisation[Title/Abstract])) AND (Pain[Title/Abstract])

*Databases used in the literature search*

The following electronic databases were searched with a strategy that spanned the time from their inception to the date of the search:

- PubMed (includes MEDLINE),
- Scopus,
- PsycArticles,
- Cochrane Library,
- Web of Science Core (use to search and then use menu on left to filter for Core option and BIOSIS), and
- BIOSIS (via Web of Science).

*Search date*

13 October 2024

#### PRISMA diagram for systematic review on pain prevalence, and its association with critical illness and mortality in hospitalised patients.

Records identified through database search conducted on 13 October 2024

n = 8 977

PubMed: 2 983 records

Scopus: 2 824 records

PsycArticles: 6 records

Cochrane library: 78

Web of Science core: 2 370

Biosis: 716

**Identification**

Duplicates removed

n = 5 270

**Screening**

Titles/abstracts screened

n = 3 707

Excluded records

n = 3 670

**Eligibility**

Full-test records assessed for eligibility

n = 31

Records included in review

n = 10

Excluded records n = 20

- Pain severity not assessed/not reported: n = 8

- Wrong study design: n = 6

- Full text not available: n = 4

- Selected cohort: n = 1

- Cohort <200: n = 1

- Cohort <18 years old n = 1

**Included**

#### Table summary for systematic review on pain prevalence, and its association with critical illness and mortality in hospitalised patients.

| **Study ID** | **Date of data collection** | **Study design** | **Country** | **World bank income classification at the time of data collection** | **Healthcare facility level** | **Sample size** | **Age (years)**  Mean ±SD, median (range) or [IQR] | **Sex**  **Male: female** | **Pain rating scale** | **Prevalence of pain**  [95%CI] | **Severity of *current* pain** | **Severity of *worst* pain in the preceding 24 hours** | **Severity of *average* pain in the preceding 24 hours** | **Severity of *least* pain in the preceding 24 hours** |
| --- | --- | --- | --- | --- | --- | --- | --- | --- | --- | --- | --- | --- | --- | --- |
| Allegue, Velasco [1] | October – December 2017 | Cross-sectional | Spain | High-income | Tertiary | 611 | 71 ± 16 (18 – 101) | 319:292 | 10cm VAS  mild pain: 1 - 3; moderate pain: 4 – 7; severe pain: 8 - 10 | 36.7%  Moderate to severe pain: 20.4% | 4 [2 – 5] |  |  |  |
| Becerra-Bolaños, Armas-Domínguez [2] | 8 June – 9 July 2021 | Cross-sectional | Spain | High-income | Tertiary | 274 | 66.17 ±14.8 | 148:126 | 10cm VAS  0: no pain; 10: maximum pain | 52.9% | 5 ± 2.8 |  |  |  |
| Beck, Dunton [3] | Not reported | Cross-sectional | USA | High-income | 326 facilities participated | 21 268 | 59 ± 18.15 (19 - +90) | 4970:7258  Missing data: 9040 | 0 – 10 anchor labels not reported | 72% |  |  | 6.03 ± 2.45 |  |
| Das, Dhar [5] | June – August 2018 | Cross-sectional | India | Lower-middle income | Tertiary | 847 | 46.8 ± 15.8 (18 – 89) | 412:435 | 0 – 10 VAS  0: no pain; 1 – 3: mild; VAS 4 – 6: moderate; and VAS 7 – 10: severe pain | 70.6%  Mild pain: 9.4%  Moderate pain: 41.1%  Severe pain: 49.5% | 6.27 ± 1.97 |  |  |  |
| Erazo-Muñoz and Colmenares-Mejía [7] | May – September 2015 | Cross-sectional | Colombia | Upper-middle income | Tertiary | 338 | 61^*^ [47 – 73] | 125:213 | 0 – 10 VAS  0: no pain; 10: worst pain you can imagine | 43.3%  Mild pain: 53.3%  Moderate pain: 31.0%  Severe pain: 15.2% | 4^*^ [1 – 6] | 8^*^ [6– 10] | 4^*^ [1 – 6] | 3^*^ [1 – 5] |
| Hajjioui, Fourtassi [8] | 29 March 2012 | Cross-sectional | Morrocco | Lower-middle income | Tertiary | 411 | 47.75 ± 17.34 (18 – 106) | 175:236 | 0 – 10 VAS anchor labels not reported | 41.6% | 4.70 ±2.01 |  | - | - |
| Jaksch, Neuwersch [9] | Not reported | Cross-sectional | Austria | High-income | 3 facilities | 1089 | 66.4 ± 17.9 | 838:784 | 0 – 10 NRS anchor labels not reported  Mild pain: 1 – 3; moderate pain: 4 – 6; severe pain: 7 – 10 | 40.0%  Mild: 48.3%  Moderate: 37.1%  Severe pain: 14.6% | 3.8 ± 2.3 | 6.0 ± 2.6 |  |  |
| Vallano, Malouf [13] | 01 January 2002 – 31 September 2003 | Cross-sectional | Spain | High-income | 15 facilities | 1675 | 58.4 ± 18.3 (18 – 98) | 864:811 | 0 – 100mm VAS  Severe pain: >60mm VAS | 48.5% [46.1 – 50.9]  Severe pain 19.7 [17 – 22.4] | 40 (10 – 100) | - |  |  |
| Van Hecke, Van Lancker [14] | October 2012 – April 2013 | Cross-sectional | Belgium | High-income | 2 facilities (1 teaching and 1 general) | 351 | 61.7 ± 18.5 | 161:190 | 0 – 10 NRS  (0: no pain; 10: worst pain imaginable) | 64.4%  Mild: 36.7%  Moderate 19.1%  Severe: 8.0% | 2.2 ± 3.6 |  |  |  |
| Visentin, Zanolin [15] | October 2000 | Cross-sectional | Italy | High-income | 20 facilities | 3931 | 67 [ 52 – 76] | 1917:1930 | 0 – 10 NRS anchor labels not reported  Mild pain: 1 – 3; moderate pain: 4 – 7; severe pain: 8 – 10 | 91.2 % [95%CI 90.3 – 92.1%]  Mild pain: 21.7%  Moderate pain: 22.9%  Severe pain: 46.6% | 7 [3 – 10] |  |  |  |

Pain severity data are presented as Mean ±SD, median (range) or [IQR], unless specified with ‘*’ for mean.

#### References for systematic review on pain prevalence, and its association with critical illness and mortality in hospitalised patients.

### Supplementary Material S2: Ethical approval processes for the African Critical Illness Outcomes Study.

| **Country** | **National ethics committee approval** | **National ethics number** | **University ethics approvals** | **Hospital ethics approvals** |
| --- | --- | --- | --- | --- |
| Botswana |  |  |  | 1 |
| Burkina Faso | Ministerie de la Sante et de L'Hygiene Publique | No number. 2 August 2023 |  | 1 |
| Congo | Ministerie l'Enseignement Superieure de la Recherche Scientifique et de l'Innovation Technologique | 053-40/MESRIT/DGRST/CERSSA/-23 |  |  |
| DRC |  |  | 1 |  |
| Egypt |  |  | 1 | 2 |
| Ethiopia | AHRI/ALERT | PQ49/23 |  |  |
| Gambia |  |  |  | 1 |
| Ghana |  |  | 1 | 2 |
| Lesotho | Ministry of Health | Ref: ID 211-2023 |  |  |
| Libya | Ministry of Health | NBC:002.H-23.10 |  |  |
| Morocco |  |  |  | 1 |
| Mozambique |  |  |  | 1 |
| Namibia | Ministry of Health and Social Services | Ref: 22/3/1/1 |  |  |
| Nigeria | National Health Research Committee of Nigeria | NHREC/01/01/2007-10/08/2023 | 1 | 14 |
| Somalia | Ministry of Health and Human Services | Ref/MOHHS/DGO/0697/Sep2023 |  |  |
| Somaliland | Ministry of Health Development | Ref: MOHD/VM:3 |  |  |
| South Africa | South African National Clinical Trial Registry | DOH-27-072023-9030 | 3 | 33 |
| Sudan |  |  |  | 2 |
| Tanzania | National Institute for Medical Research | NIMR/HQ/R.8a/Vol.IX/4473 |  |  |
| Tunisia |  |  | 1 | 4 |
| Uganda | Mbale Regional Referral Hospital Research & Ethics Committee and Uganda National Council for Science & Technology | MRRH-2023-304  HS3338ES |  | 1 |
| Zimbabwe | Medical Research Council of Zimbabwe | MRCZ/E/346 |  |  |

### Supplementary Material S3: Broadcasting document

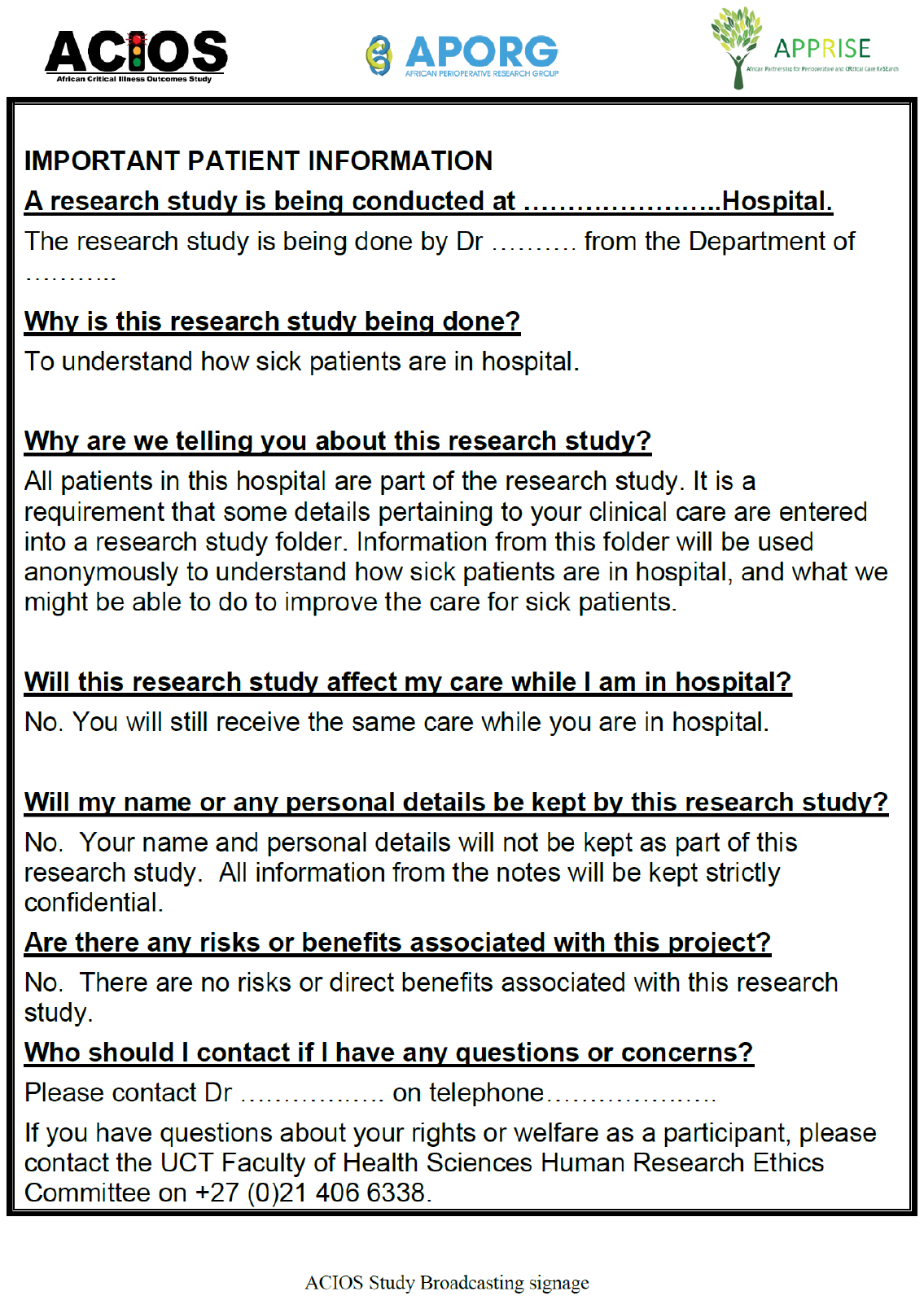

### Supplementary Material S4. Severity of pain in hospitalised patients in Africa: Statistical Analysis Plan

**The African Critical Illness Outcomes Study (ACIOS)**

A prospective, multi-country, multi-centre, observational study to determine the hospital point-prevalence and mortality rates of adult patients with critical illness in acute hospitals in Africa.

Sub-study: Severity of pain in hospitalised patients in Africa

**Statistical Analysis Plan (SAP)**

Version 1.0

Date: 18/03/2024

Registration ClinicalTrials.gov - NCT06051526

Based on “ACIOS protocol version 2.0 HREC approved”

| **Persons contributing to the analysis plan** | |
| --- | --- |
| **Names and positions** | Assoc Professor Tim Baker, MUHAS  Professor Bruce Biccard, UCT  Professor Rupert Pearse, QMUL  Dr Carl Otto Schell, KI  Anneli Hardy (Statistician)  Gillian J Bedwell, UCT  Assoc Professor Victoria J Madden, UCT  Professor Romy Parker, UCT |
| **Authorisation** | |
| **Position** | **Chief Investigator** |
| **Name** | Assoc Professor Tim Baker, MUHAS |
| **Date** | 13^th^ February 2024 |
| **Position** | **Trial Statistician** |
| **Name** | Anneli Hardy |
| **Date** | 2024-02-13 |

**Remit of the SAP**

The purpose of this document is to provide details of the statistical analyses and presentation of results to be reported within the sub-study – “Severity of pain in hospitalised patients” – of the ACIOS study. It is important to set these out and to agree them in advance of inspecting the outcome data for the study, so that data derived decisions in the analysis are avoided. Any exploratory, post hoc, or unplanned analysis will be clearly identified as such in the study analysis report.

**Timing of the SAP**

The SAP version 1.0 was written prior to the investigators having access to the data.

1. Study Summary

| **Short title** | ACIOS – pain severity |
| --- | --- |
| **Methodology** | A prospective, international, multi-centre, observational study. |
| **Research sites** | Acute hospitals in African countries. |
| **Objective** | To determine the severity of self-reported pain in hospitalised patients across Africa. |
| **Number of patients** | Not specified. All eligible patients in participating hospitals. |
| **Inclusion criteria** | All in-hospital patients aged 18 years or older in all departments and wards in participating hospitals in Africa. |
| **Exclusion criteria** | None |
| **Patient follow-up** | Until hospital discharge or death, censored at 7 days after inclusion. |
| **Primary outcomes** | 1. To determine patients’ self-reported severity of pain in the preceding 24 hours on a scale of 0 – 10. 2. To determine the association between severity of pain and critical illness in adult patients admitted to hospitals across Africa. 3. To determine the association between severity of pain and mortality in adult patients admitted to hospitals across Africa. |
| **Data collection duration** | One day in each hospital in September-December 2023 plus 7 days follow-up in each hospital |
| **Proposed start date** | 7^th^ September 2023 |
| **Proposed end date** | 27^th^ December 2023 |

**Introduction**

Pain is poorly managed in hospitalised patients ^1^. Uncontrolled pain in hospitalised patients is strongly associated with both short- and long-term complications. Patients are more likely to develop secondary complications such as sepsis, bleeding, and infarctions if their pain is not adequately managed ^2,3^. Further, pain of >7 on a visual analogue scale is strongly associated with an increased risk of persistent pain ^4^. In high-income countries, up to 86% of hospitalised patients report having poorly managed pain ^1,5^. However, there is a paucity of the literature on the severity of acute pain in hospitalised patients in Africa. Therefore, this sub-study aims to determine the severity of self-reported pain in hospitalised patients across Africa; and the relationship between severity of pain and (1) critical illness and (2) mortality. This study falls under the ACIOS – African Critical Illness Outcome Study – parent study.

**Statistical Analysis Plan**

*General analysis principles*

Data will be presented at a continental African level. All institutional and national level data will be anonymised prior to publication. Categorical variables will be described as proportions and will be compared using chi-square tests. Continuous variables will be described as mean and standard deviation if normally distributed or median and inter-quartile range (IQR) if not normally distributed. No comparisons between groups will be performed at a univariate level.

For the analysis of the objectives, we will present the following information:

- The number of patients included in each analysis.
- Summary statistics of the outcome (e.g. median (IQR), mean (SD), number (%), range).
- A point estimate, odds ratio or hazard ratio with 95% confidence intervals.
- A two-sided p-value with a significance level of <0.05 will be used where relevant.

For data that are not necessary for each objective, imputation of missing observations will not be made and will be reported descriptively.

Statistical analyses will be performed using the Statistical Package for the Social Sciences (SPSS) version 28.0.1.1 (SPSS Inc., Chicago, IL, USA) and R (version 4.2.1) in RStudio ^6^.

**Sample Size / Recruitment**

As many sites as possible will be recruited in participating countries. All adult patients will be eligible for inclusion in the sites. A sensitivity analysis will be done for each objective including only data from hospitals that recruited >90% of eligible patients. We do not have a specific sample size and statistical models will be adapted to the event rates provided by the sample recruited. Participation in the study, and completeness of follow-up will be illustrated by a STROBE flow diagram.

Patient recruitment and description will be presented as follows:

- STROBE flow diagram including i) countries, ii) number of eligible patients, iii) patients included and excluded.
- The number of participating hospitals, hospital characteristics and patients at each hospital level will be reported in a table. Detailed hospital characteristics will be provided in a Supplementary Table.
- The patient characteristics of the cohort will be presented in summary tables.

**Objectives**

1. To determine patients’ self-reported severity of pain in the preceding 24 hours on a scale of 0 – 10.
2. To determine the association between severity of pain and critical illnesses in adult patients admitted to hospitals across Africa.
3. To determine the association between severity of pain and mortality in adult patients admitted to hospitals across Africa.

**Statistical analysis plan for Objective 1 “*self-reported pain severity*”**

Using the appropriate summary statistics, we will report patients’ self-reported report pain severity in the preceding 24 hours from data collection on a scale of 0 – 10, where 0 is ‘no pain’ and 10 is ‘the worst pain you can imagine’. We will report pain severity across all participants as well as for the following groups: patients with critical illness; patients without critical illness; patients who died; and patients who survived.

Some patients were unable to give pain ratings (e.g. patients who were unconscious at the time of data collection). Therefore, we will also present the number of patients who provided pain reports.

Dummy Table 1. Self-reported pain severity of the African Critical Illness Outcomes Study (ACIOS) patient cohort

|  | **All patients (n=?)** | **Patients with critical illness (n=?)** | **Patients without critical illness (n=?)** | **Patients who died (n=?)** | **Patients who survived (n=?)** |
| --- | --- | --- | --- | --- | --- |
| **Pain severity (0 – 10)**  *[presented using the appropriate summary statistics]* |  |  |  |  |  |

**Statistical analysis plan for Objective 2 “association between severity of pain and critical illness”**

Data on ‘critical illness’ will be extrapolated from the parent study ACIOS.

We will conduct univariate and multivariate logistic regression models to determine the association between severity of pain (continuous variable) and critical illness (binary variable). Severity of pain will be the independent variable and critical illness will be the dependent variable. The independent patient associated with critical illness identified in the primary paper will be included in the multivariate model. We will follow best practice by using both visual data analysis and formal modelling to investigate this relationship. The specifics of the models will be determined by the data features to achieve the best fitting model that is interpretable.

We will use a three-level generalized mixed model, with patients being at the first level, hospital at the second and country at the third level, to account for the expected correlation in outcomes within hospitals and countries.

**Statistical analysis plan for Objective 3 “association between severity of pain and mortality”**

We will present the number and proportion of critically ill and non-critically ill patients who die in hospital within the 7 days of data collection. The defined time for the outcomes is from the point of inclusion of the patient into the study to hospital discharge or death, censored at 7 days. Patients discharged alive are not followed up at home. Patients still in hospital receiving therapy at 7 days will be regarded as “alive” and included in the study.

We will conduct univariate and multivariable logistic regression models to determine the association between severity of pain (continuous variable) and mortality (binary variable). Severity of pain will be the independent variable and mortality will be the dependent variable. Additional patient factors identified to be independently associated with mortality will be included in the multivariate model. We will follow best practice by using both visual data analysis and formal modelling to investigate this relationship. The specifics of the models will be determined by the data features to achieve the best fitting model that is interpretable.

We will use a three-level generalized mixed model, with patients being at the first level, hospital at the second and country at the third level, to account for the expected correlation in outcomes within hospitals and countries.

A Kaplan-Meier graph will be constructed of the in-hospital mortality from Day 0 to Day 7 for patients with severe (>7 on VAS) pain and patients *without* severe pain (<7 on VAS). Time will be counted from recruitment to the study until discharge, death or censored. The graph will visualise how mortality risk changes over time. A log-rank test for equality of the survival functions will be performed if the assumptions necessary for using the test hold.^7^

*Missing data:* data from patients lost to follow up (missing outcome data) will be included in the statistical analysis without imputation and reported descriptively. Data from these patients will not be included in the mortality analysis (i.e. objective 3), but will be included in other analyses (i.e. objectives 1 and 2).

### Supplementary Material S5: Statistical analysis plan amendment

**The African Critical Illness Outcomes Study (ACIOS)**

Parent study: A prospective, multi-country, multi-centre, observational study to determine the hospital point-prevalence and mortality rates of adult patients with critical illness in acute hospitals in Africa.

Sub-study: Prevalence and severity of pain in hospitalised patients in Africa

**Statistical Analysis Plan: amendment**

This document is an amendment to the original statistical analysis plan: version 1, date: 18/03/2024, protocol registration ClinicalTrials.gov - NCT06051526.

The original statistical analysis plan included 3 objectives, for which the analyses have already been conducted at the time of preparing and locking this document. These objectives were:

1. To determine patients’ self-reported severity of pain in the 24 hours before assessment, on a scale of 0 – 10.
2. To determine the association between severity of pain and critical illness status in adult patients admitted to hospitals across Africa.
3. To determine the association between severity of pain and mortality in adult patients admitted to hospitals across Africa.

Severe pain (pain of ≥7 out of 10 on a 0 – 10 scale) is associated with adverse clinical outcomes, including the development of chronic pain^1, 2, 3^. Given the clinical significance of severe pain, we have opted to amend our statistical analysis plan to assess the potential clinical significance of severe pain in this dataset. Therefore, we will now include analyses of “severe pain status” as an independent variable predicting 1) critical illness and 2) mortality, alongside analyses of whether the prevalence of “severe pain status” 3) differs by patient sex or 4) is predicted by age. Additionally, we will include a model investigating whether age interacts with sex in predicting the prevalence of severe pain. Table 1 summarises the SAP amendments.

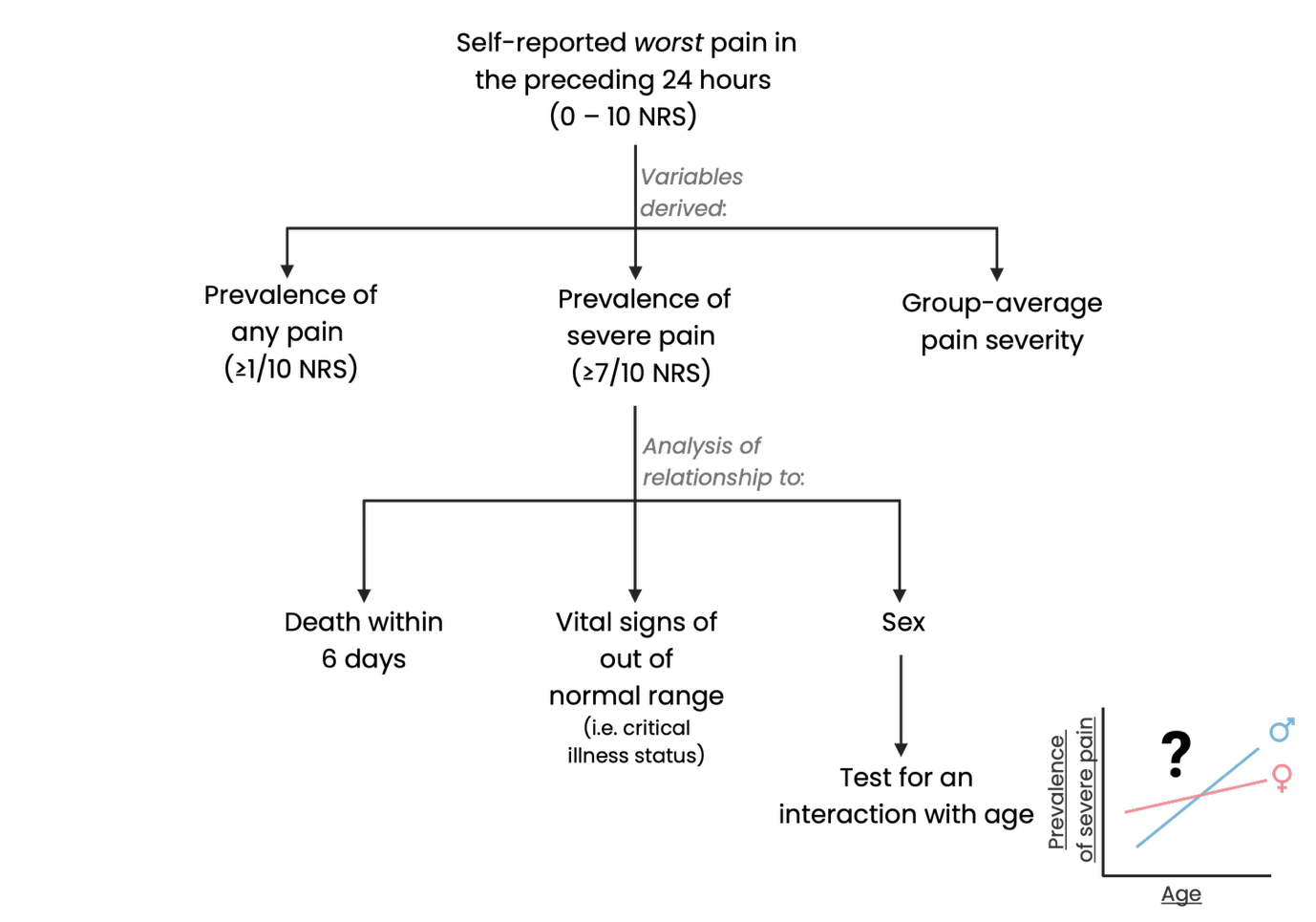

Figure 1: Flow diagram illustrating the analytical decision-making process

Table 1: Summary of the amendments to the statistical analysis plan. Rows coloured in green indicate that the analyses have already been conducted at the time of preparing and locking this document. Rows coloured in orange indicate that the analyses have not yet been conducted at the time of preparing and locking this document.

| **Research question** | **Descriptive statistics** | | | **Model structure/Approach** | **Notes** |
| --- | --- | --- | --- | --- | --- |
| What is the prevalence and severity of pain in hospitalised patients across Africa? | 1. Prevalence of any pain (≥1/10) (95%CI) [range] | 2. Average severity as median [IQR] | 3. Prevalence of severe pain (≥7/10) (95%CI) [range] | 1. Calculate prevalence within each hospital 2. Calculate and report prevalence (and variance) across hospitals. | In acknowledgement of the importance of understanding variability in pain management as a clinical outcome, the prevalence of pain and of severe pain will each be calculated for each hospital and then pooled to provide a sample estimate, allowing insight to the hospital-to-hospital variance in prevalence. |
| Does the prevalence of severe pain differ by sex and age? | - | - | Tables summarising the prevalence of severe pain for males and for females.  Plot displaying the interaction of age and sex on the prevalence of severe pain. | Step 1: Prev_severe_pain ~ sex + (1\|hospital:country)  Step 2: Prev_severe_pain ~ sex*age + (1\|hospital:country)  Dependent variable: prevalence of severe pain within each hospital  Independent variable: age (continuous); sex (binary) | Both sex and age shape behaviours such as pain reporting and care delivery. Therefore, we will model prevalence of severe pain as a function of sex to estimate the average difference in severe pain prevalence between males and females. We will then extend this to a sex*age interaction model to determine whether those sex differences are modified by age. |
| Is critical illness associated with severe pain? | - | - | - | Critical_illness_status ~ Severe_pain_status + covariates + (1\|hospital:country)  Dependent variable: critical illness status (binary)  Independent variable: severe pain status (binary)  Covariates:   - age, - sex, - urgency of admission (elective vs emergency/acute, - main category of admission (non-communicable disease, trauma, infection, or maternal health), - chronic diseases (hypertension, diabetes, cancer, chronic obstructive pulmonary disease or asthma, heart disease, HIV/AIDS, tuberculosis, or other), - pregnancy | These covariates were pragmatically determined; we collected data only on patient factors we considered clinically important for in-hospital critical illness and mortality and that would be readily available from hospital records. |
| Is 7-day mortality associated with severe pain? | - | - | - | Mortality_status ~ Severe_pain_status + covariates + (1\|hospital:country)  Dependent variable: mortality status (binary)  Independent variable: severe pain status (binary)  Covariates:   - age, - urgency of admission (elective vs emergency/acute, - main category of admission (non-communicable disease, trauma, infection), - chronic diseases (cancer, HIV/AIDS), - critical illness | The primary ACIOS study ^4^ reported that the following eight variables were independently associated with mortality: age, cancer, HIV infection, emergency surgery, critical illness, and admission for infection, non-communicable disease, and trauma. Therefore, we included only these eight confounders in the multivariable regression model investigating the relationship between severe pain and mortality. |
| Is critical illness associated with pain severity? | - | - | - | Critical_illness_status ~ pain_severity + covariates + (1\|hospital:country)  Dependent variable: critical illness status (binary)  Independent variable: pain severity (continuous)  Covariates:   - age, - sex, - urgency of admission (elective vs emergency/acute, - main category of admission (non-communicable disease, trauma, infection, or maternal health), - chronic diseases (hypertension, diabetes, cancer, chronic obstructive pulmonary disease or asthma, heart disease, HIV/AIDS, tuberculosis, or other),   pregnancy | This analysis has already been conducted and will be included in the supplementary files. |
| Is 7-day mortality associated with pain severity? | - | - | - | Mortality_status ~ pain_severity + covariates + (1\|hospital:country)  Dependent variable: mortality status (binary)  Independent variable: pain severity (continuous)  Covariates:   - age, - urgency of admission (elective vs emergency/acute, - main category of admission (non-communicable disease, trauma, infection), - chronic diseases (cancer, HIV/AIDS), - critical illness | This analysis has already been conducted and will be included in the supplementary files. |

### Supplementary Material S6. Hospital data record form

**African Critical Illness Outcomes Study (ACIOS) – Hospital CRF**

**Section 1: Hospital characteristics**

1. Language preference:	⬜ English	⬜ French	⬜ Arabic ⬜ Portuguese ⬜ Other …………………………
2. Hospital name:	………………………………………………………
3. Country:	………………………………………………………
4. Level of hospital:

⬜ First-level (e.g. district)	⬜ Second-level (e.g. Regional)	⬜ Third-level (e.g. University/Central/National)

5. Type of hospital:	⬜ Government	⬜ Private	⬜ Charitable
6. Total number of hospital beds:	Total
7. Number of beds in High Care Units:	Total
8. Number of beds in ICUs:	Total
9. Population served (catchment) of the hospital:

### Supplementary Material S7. Patient data record form

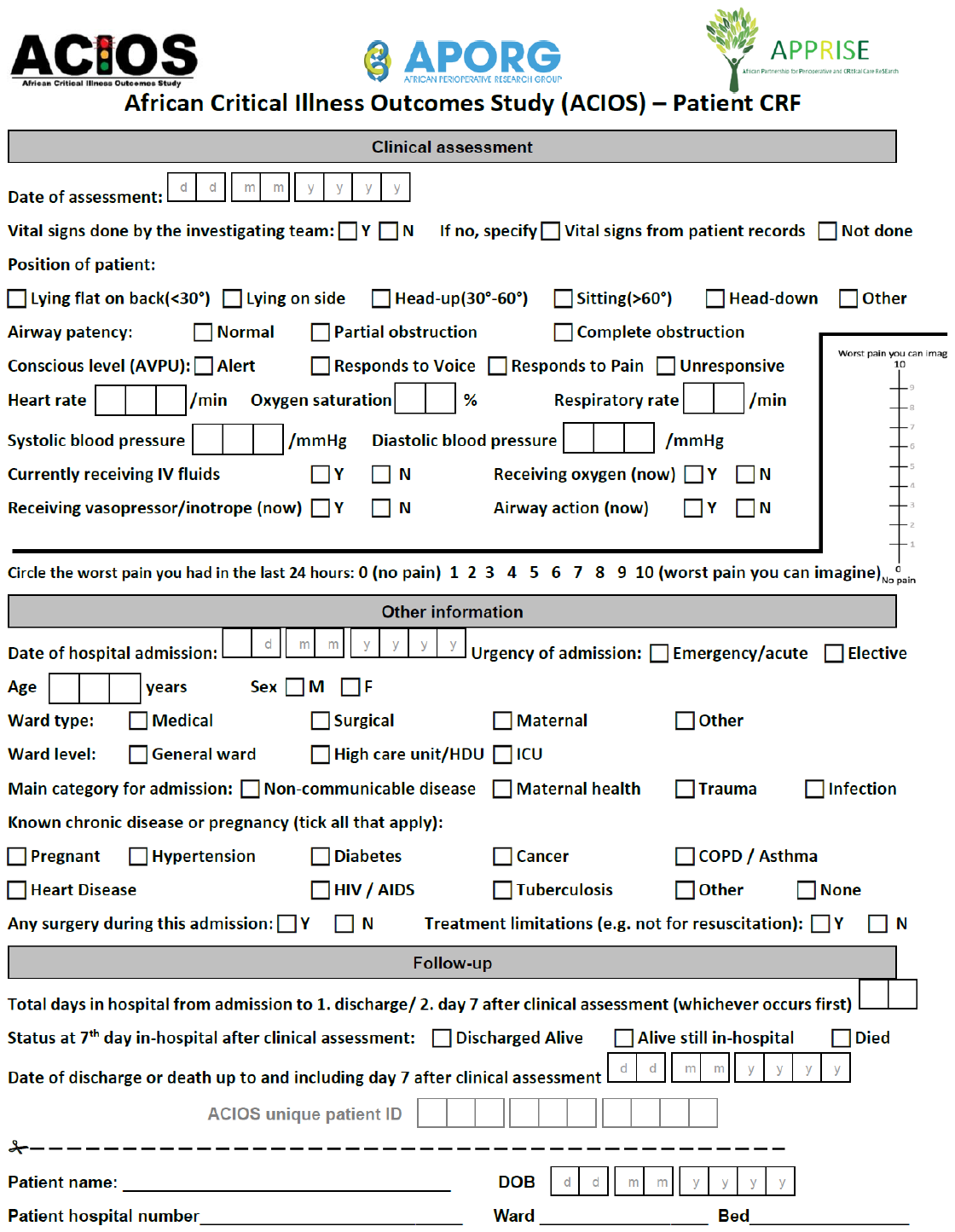

### Supplementary Material S8. African Critical Illness Outcomes Study (ACIOS) definitions

**Definitions:**

**Position of patient:** The position the patient is in the bed/chair when the investigating team arrive at the bedside. The number of degrees refers to the angle of the head and body compared to the legs.

**Airway patency:** Normal is an unobstructed airway. Partial obstruction may be indicated by stridor, secretions in the airway identified by gurgling, or snoring. Complete obstruction is evident by a see-saw chest movement (chest down and abdomen up with attempted breathing against a closed glottis). Complete obstruction is an airway emergency and requires calling the attending clinical team immediately.

**Conscious level AVPU:** Measurement of conscious level. Is the patient Alert = A. If they are not alert but they respond to your voice = V. If they don’t respond to voice but respond to a painful stimulus = P. If they remain unresponsive even with a painful stimulus = U (unresponsive).

**Currently receiving IV fluids**: At the time of clinical assessment, the patient is receiving IV fluids if IV fluids are hanging at the bedside, and currently dripping into an intravenous cannula, or the patient has been receiving fluids as described within the previous few minutes but is now finished and a new fluid is being prepared to be administered.

**Receiving oxygen (now):** At the time of clinical assessment, the patient is receiving oxygen if supplementary oxygen is currently flowing into a nasal cannula, face mask or other delivery device that is correctly fitted so that oxygen is entering the patient’s lungs.

**Receiving vasopressor/inotrope (now):** Ongoing care with a vasopressor or inotrope infusion – for example noradrenaline, adrenaline, dopamine or dobutamine.

**Airway action (now):** An action to open the airway or maintain a free airway. For example: chin lift, jaw thrust, oro-pharyngeal airway, naso-pharyngeal airway, intubated patient.

Receiving Essential Emergency and Critical Care (EECC): Patients will be deemed to be receiving EECC if they are:

• critically ill due to the conscious level criterion and:

o	are lying in the lateral position or

o	have an oro-pharyngeal or naso-pharyngeal airway inserted in their pharynx or

o	have an ongoing chin-life or jaw-thrust or

o	have other airway protection.

• critically ill due to a respiratory criterion and:

o	are receiving oxygen.

• critically ill due to a circulatory criterion and:

o	are receiving intravenous fluids or

o	are receiving a vasopressor or inotrope.

**High care unit/HDU:** A unit or ward or part of a ward which is dedicated to providing an increased level of care when compared to a general ward. High care units often have increased nurse:patient ratios, more equipment and more advanced care such as oxygen, CPAP, vasopressors etc. This does *not* include units with mechanical ventilation, as that is an ICU. Includes recovery rooms providing an increased level of care.

**ICU:** A unit or ward which is dedicated to providing an increased level of care when compared to a general ward or high care unit including mechanical ventilation.

**Main category for admission:** The main diagnosis or reason that the patient is being treated in hospital.

**Treatment limitations:** A patient has a treatment limitation if the clinical team have made the clinical judgement that some treatments would not be in the patient’s best interest. For example “DNR” (do not resuscitate in the event of a cardiac arrest), or “Not for ICU” in the event of deterioration.

**Days in hospital:** Total number of days in hospital from admission.

**Status at hospital discharge or 7^th^ day in-hospital after clinical assessment:** The survival status of the patient at hospital discharge, or at the 7th day after clinical assessment (if the patient had not yet been discharged). The study is censored at the 7^th^ day after clinical assessment.

### Supplementary Material S9. Distribution of pain ratings.

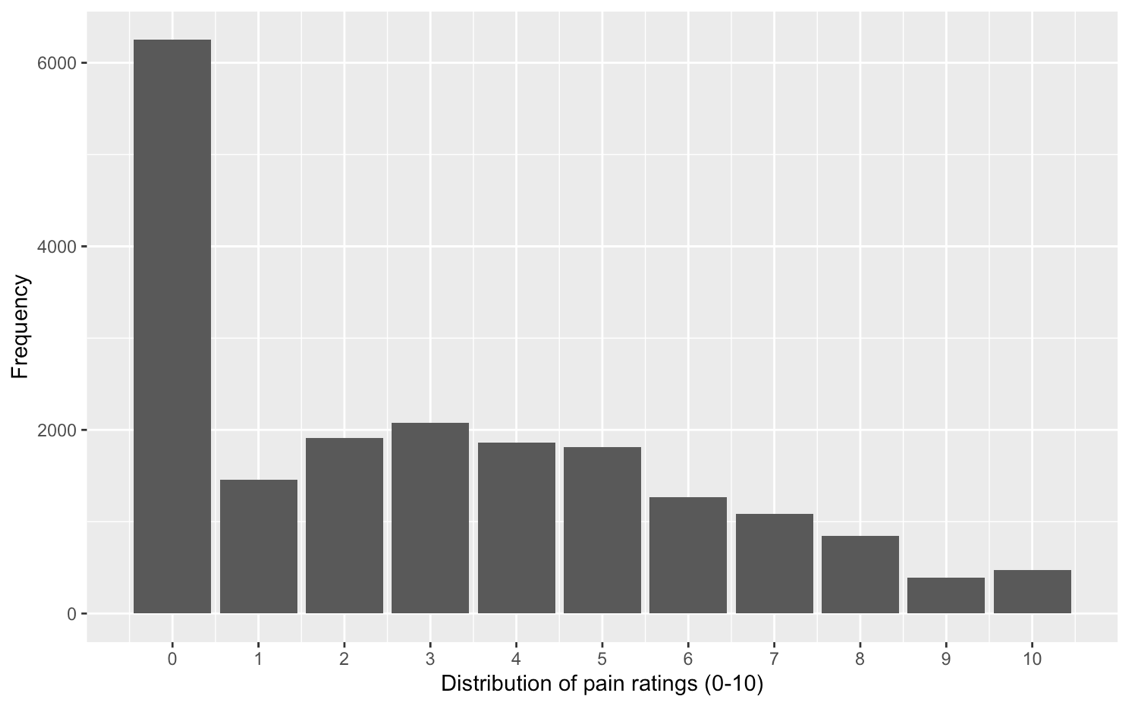

### Supplementary Material S10. Participating countries, hospitals and participants recruited

| **Country** | **Hospitals** | **Participants** | **Proportion of participants** |
| --- | --- | --- | --- |
| Botswana | 1/180 | 354/19872 | 1.8% |
| Burkina Faso | 4/180 | 1226/19872 | 6.2% |
| Congo | 1/180 | 82/19872 | 0.4% |
| DRC | 4/180 | 42/19872 | 0.2% |
| Egypt | 2/180 | 265/19872 | 1.3% |
| Ethiopia | 8/180 | 1041/19872 | 5.2% |
| Gambia | 4/180 | 266/19872 | 1.3% |
| Ghana | 18/180 | 1933/19872 | 9.7% |
| Lesotho | 2/180 | 30/19872 | 0.2% |
| Libya | 28/180 | 1613/19872 | 8.1% |
| Morocco | 1/180 | 246/19872 | 1.2% |
| Mozambique | 1/180 | 363/19872 | 1.8% |
| Namibia | 7/180 | 677/19872 | 3.4% |
| Nigeria | 19/180 | 2286/19872 | 11.5% |
| Somalia | 3/180 | 224/19872 | 1.1% |
| Somaliland | 3/180 | 74/19872 | 0.4% |
| South Africa | 37/180 | 6167/19872 | 31.0% |
| Sudan | 2/180 | 39/19872 | 0.2% |
| Tanzania | 16/180 | 1338/19872 | 6.7% |
| Tunisia | 5/180 | 512/19872 | 2.6% |
| Uganda | 13/180 | 1055/19872 | 5.3% |
| Zimbabwe | 1/180 | 39/19872 | 0.2% |
| **DCP-3 level of hospital*** | |  |  |
| Level 1 | 56/173 | 2657/19220 | 13.8% |
| Level 2 | 38/173 | 3846/19220 | 20.0% |
| Level 3 | 79/173 | 12717/19220 | 66.2% |
| **Type of hospital** | | | |
| Government | 152/173 | 18472/19477 | 94.8% |
| Private | 19/173 | 704/19477 | 3.6% |
| Charitable | 5/173 | 301/19477 | 1.5% |
| **Number of beds**  Median (IQR) | | | |
| Total hospital beds | 265 (122 – 519) | | |
| Beds in high care units* | 7 (3 – 16) | | |
| Beds in intensive care units** | 7 (2 – 12) | | |
| Population served (catchment) of the hospital | 350 000 (91 715 – 2 077 500) | | |

Data are n/N (%). Denominators vary according to completeness of data

* 8/180 hospitals did not report the number of beds in high care units. 36/172 hospitals (20.9%) reported no high care beds.

**7/180 hospitals did not report the number of beds in intensive care units. 42/173 (24.9%) reported no intensive care beds.

**Definitions of hospital levels**

| **Level of care** | **Alternative terms commonly found in the literature** |
| --- | --- |
| *First-level hospitals:* Few specialties—mainly internal medicine, obstetrics and gynaecology, paediatrics, and general surgery; often only one general practice physician or a nonphysician practitioner; limited laboratory services available for general but not specialized pathological analysis; from 50 to 250 beds. | Primary-level hospital |
|  | District hospital |
|  | Rural hospital |
|  | Community hospital |
|  | General hospital |
| *Second-level hospitals:* More differentiated by function with as many as 5 to 10 clinical specialties; from 200 to 800 beds. | Regional hospital |
|  | Provincial hospital (or equivalent administrative area such as county) |
|  | General hospital |
| *Third-level hospitals:* Highly specialized staff and technical equipment—for example, cardiology, intensive care unit, and specialized imaging units; clinical services highly differentiated by function; could have teaching activities; from 300 to 1,500 beds. | National hospital |
|  | Central hospital |
|  | Academic or teaching or university hospital |

### Supplementary Material S11. Data missingness for the generalised linear mixed model

| **Variables** | **Missing (n)** | **Missing (%)** | **Valid (n)** |
| --- | --- | --- | --- |
| Age | 13 | 0.1% | 19425 |
| Critical illness | 118 | 0.6% | 19320 |
| Mortality | 85 | 0.4% | 19353 |
| Sex | 9 | 0.1% | 19429 |
| Pregnant | 0 | 0% | 19438 |
| Hypertension | 0 | 0% | 19438 |
| Diabetes | 0 | 0% | 19438 |
| Cancer | 0 | 0% | 19438 |
| COPD/ Asthma | 0 | 0% | 19438 |
| Heart disease | 0 | 0% | 19438 |
| HIV/AIDS | 0 | 0% | 19438 |
| Tuberculosis | 0 | 0% | 19438 |
| Other comorbidities | 0 | 0% | 19438 |
| Urgency of admission | 88 | 0.5% | 19350 |
| Main category for admission | 63 | 0.3% | 19375 |
| Patient position | 13 | 0.1% | 19425 |
| Airway patency | 17 | 0.1% | 19421 |
| Conscious level | 20 | 0.1% | 19418 |
| Heart rate | 18 | 0.1% | 19420 |
| Oxygen saturation | 29 | 0.1% | 19409 |
| Respiratory rate | 90 | 0.5% | 19348 |
| Systolic blood pressure | 13 | 0.1% | 19425 |
| Diastolic blood pressure | 14 | 0.1% | 19424 |

Data are n/N (%). Denominators vary with the completeness of the data. COPD chronic obstructive pulmonary disease; HIV/AIDS human immunodeficiency virus/ acquired immunodeficiency syndrome.

### Supplementary Material S12. Model diagnostics and fit for the logistic regression model with critical illness as the outcome

Model diagnostics and fit for the logistic regression model was assessed using simulated residuals generated by the DHARMa package in R (Hartig F, 2024). The multivariable logistic regression model indicated minor deviations from uniformity (Figure S3).

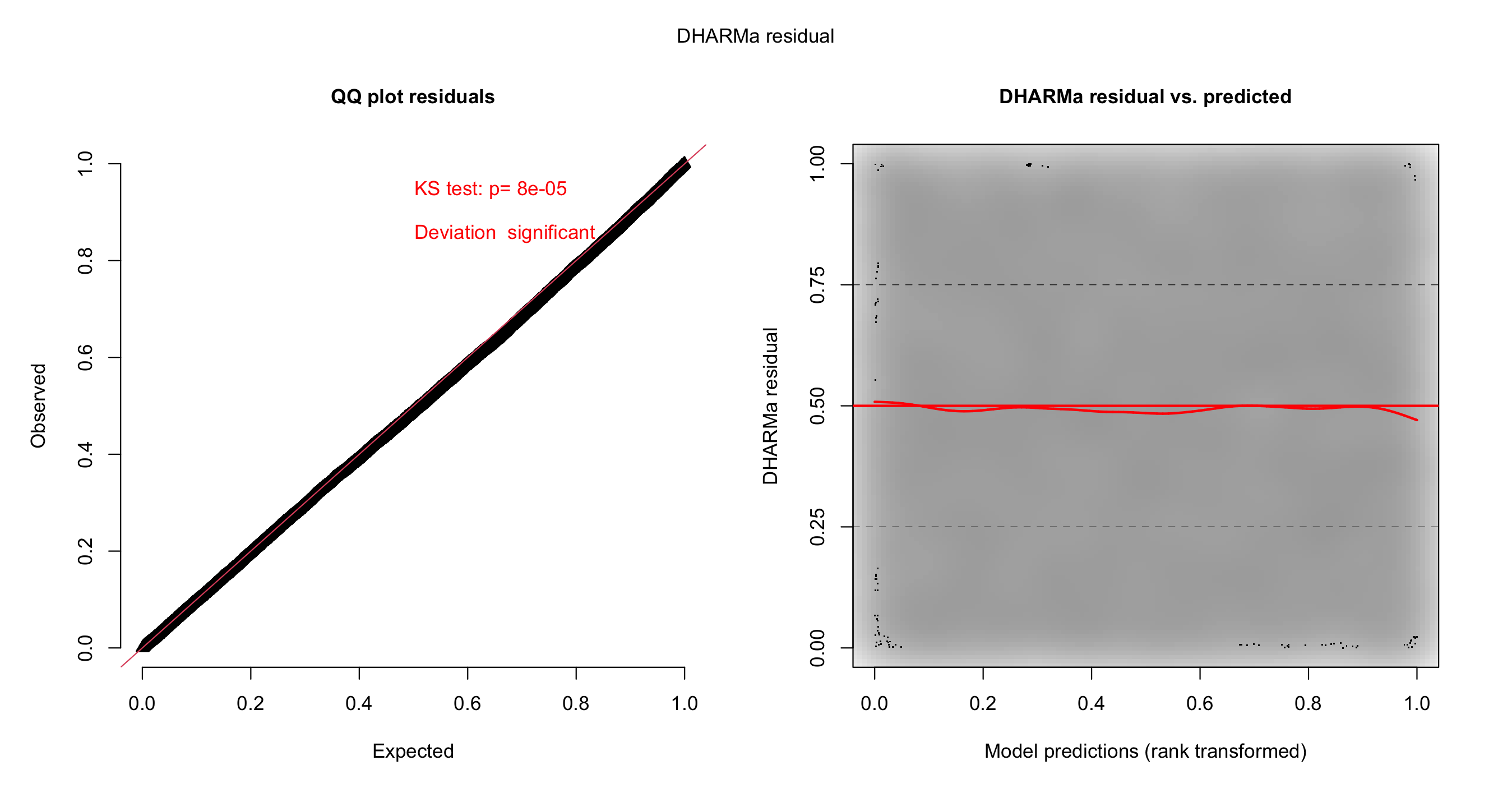

**Figure S3. Fit for multivariable logistic regression model with critical illness as the outcome.**

The assumption of linearity for the continuous predictors in the multivariable logistic regression model was assessed using partial-residual plots to detect deviations from linearity. The quantile test suggested some non-linearity for age. No further correction was applied in this supplementary analysis. The partial-residual plot for age in the univariable logistic regression model including age as a covariate is shown in Figure S4a, and in the multivariable model including age as a covariate in Figure S4b.

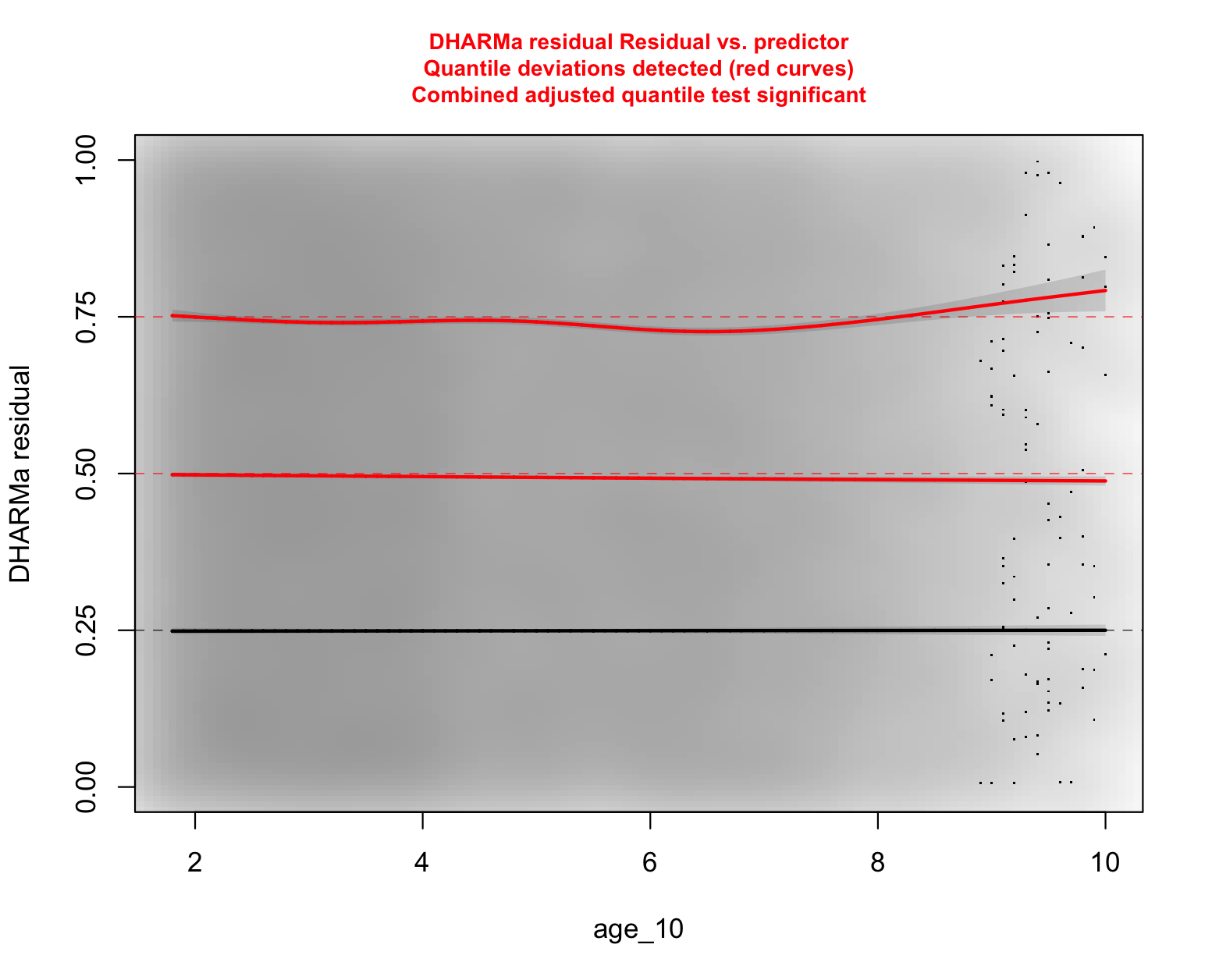

**Figure S4a. Partial-residual plot for age in the univariable logistic regression model**

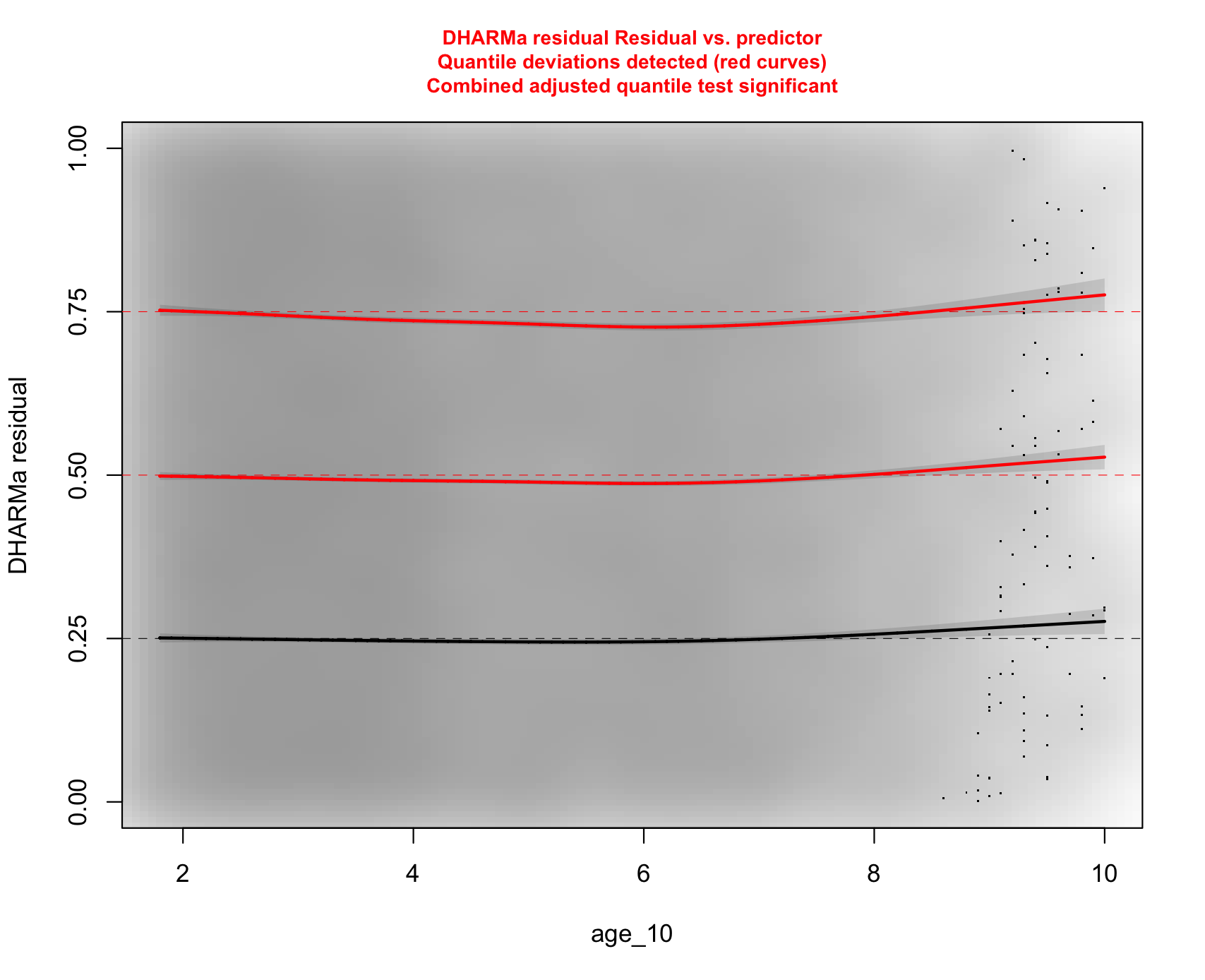

**Figure S4b. The partial-residual plot for age in the multivariable logistic regression model including age as a covariate.**

### Supplementary Material S13. Model diagnostics and fit for the logistic regression model with mortality as the outcome

Model diagnostics and fit for the logistic regression model was assessed using simulated residuals generated by the DHARMa package in R (Hartig F, 2024). The multivariable logistic regression model showed acceptable fit, and no significant violations of any model assumptions (Figure S1).

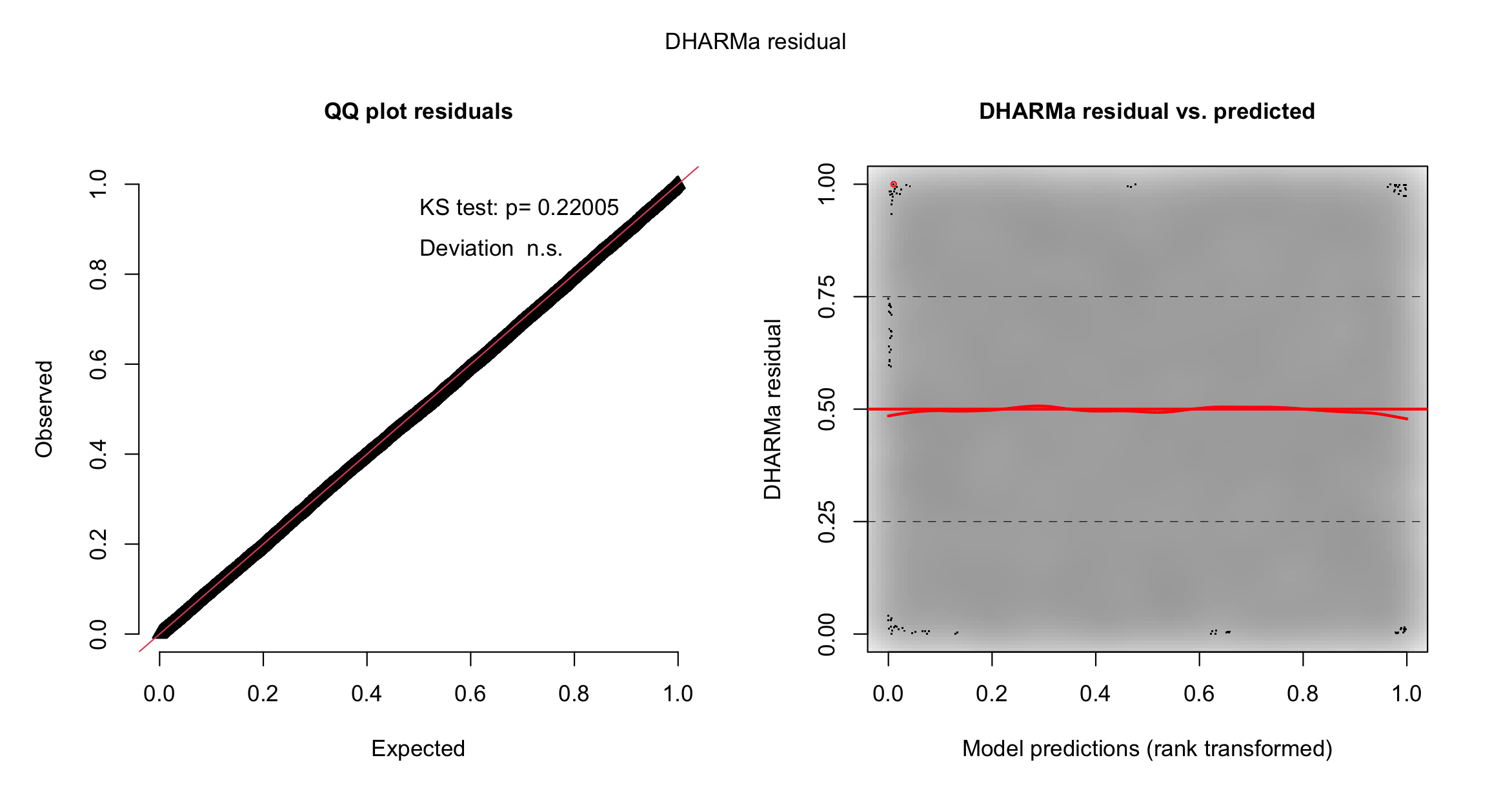

**Figure S1. Fit for multivariable logistic regression model with mortality as the outcome.**

The assumption of linearity for the continuous predictors in the multivariable logistic regression model was assessed using partial-residual plots to detect deviations from linearity. The multivariable logistic regression models did not show violations of the assumption of linearity. The partial-residual plot for age in the univariable logistic regression model including age as a covariate is shown in Figure S2a, and in the multivariable model including age as a covariate in Figure S2b.

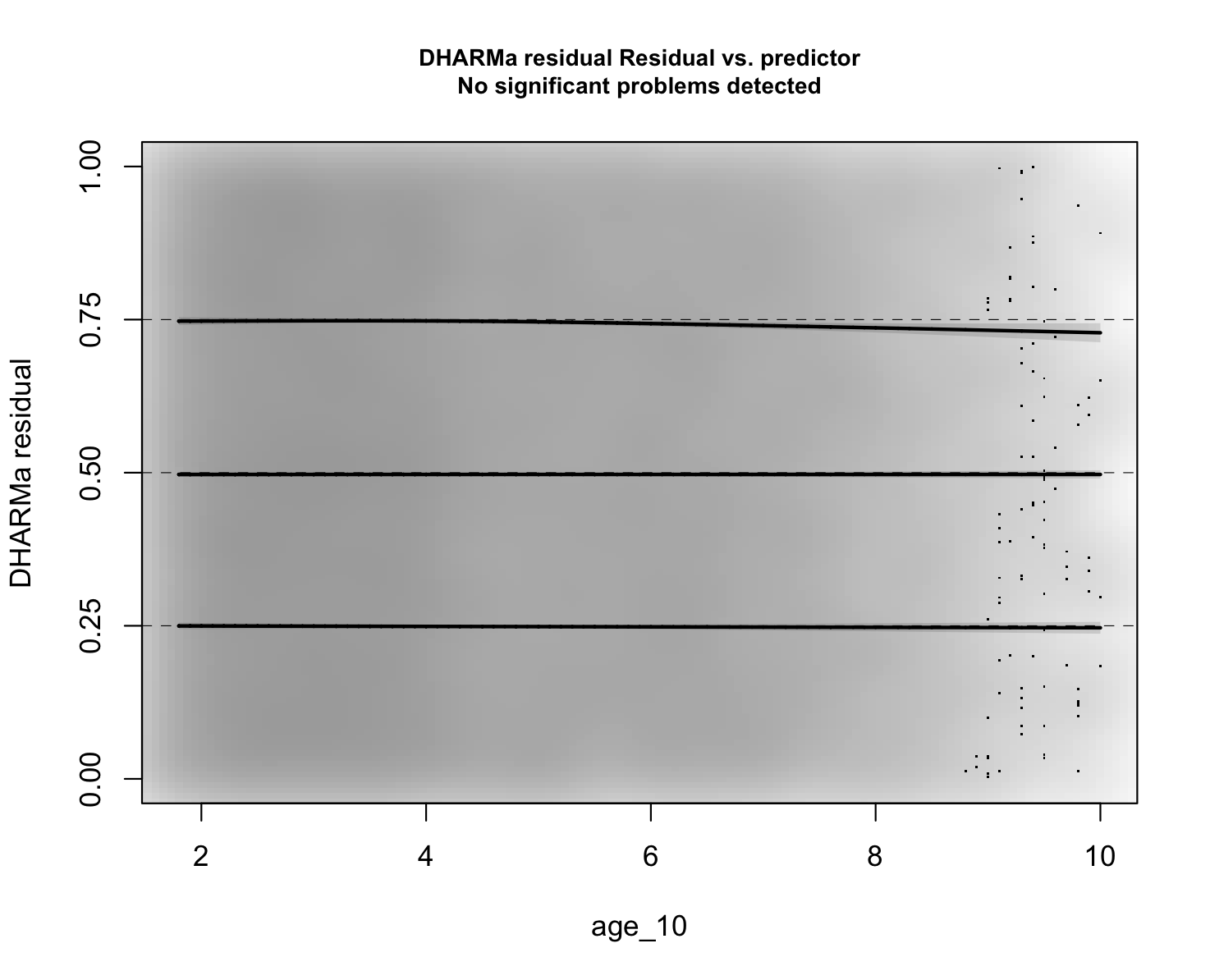

**Figure S2a. Partial-residual plot for age in the univariable logistic regression model**

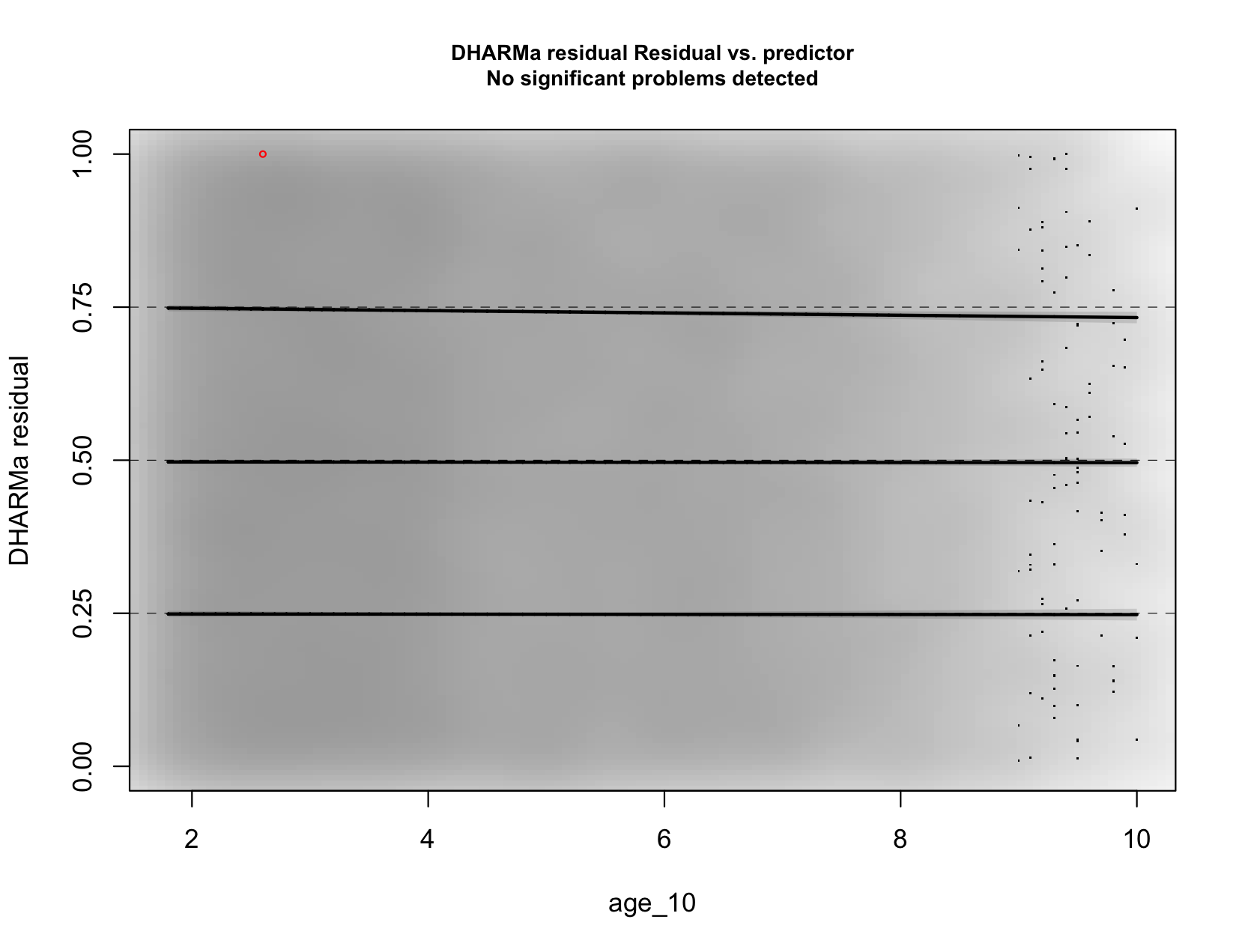

**Figure S2b. The partial-residual plot for age in the multivariable logistic regression model including age as a covariate.**

### Supplementary Material S14. Kaplan-Meier Plot

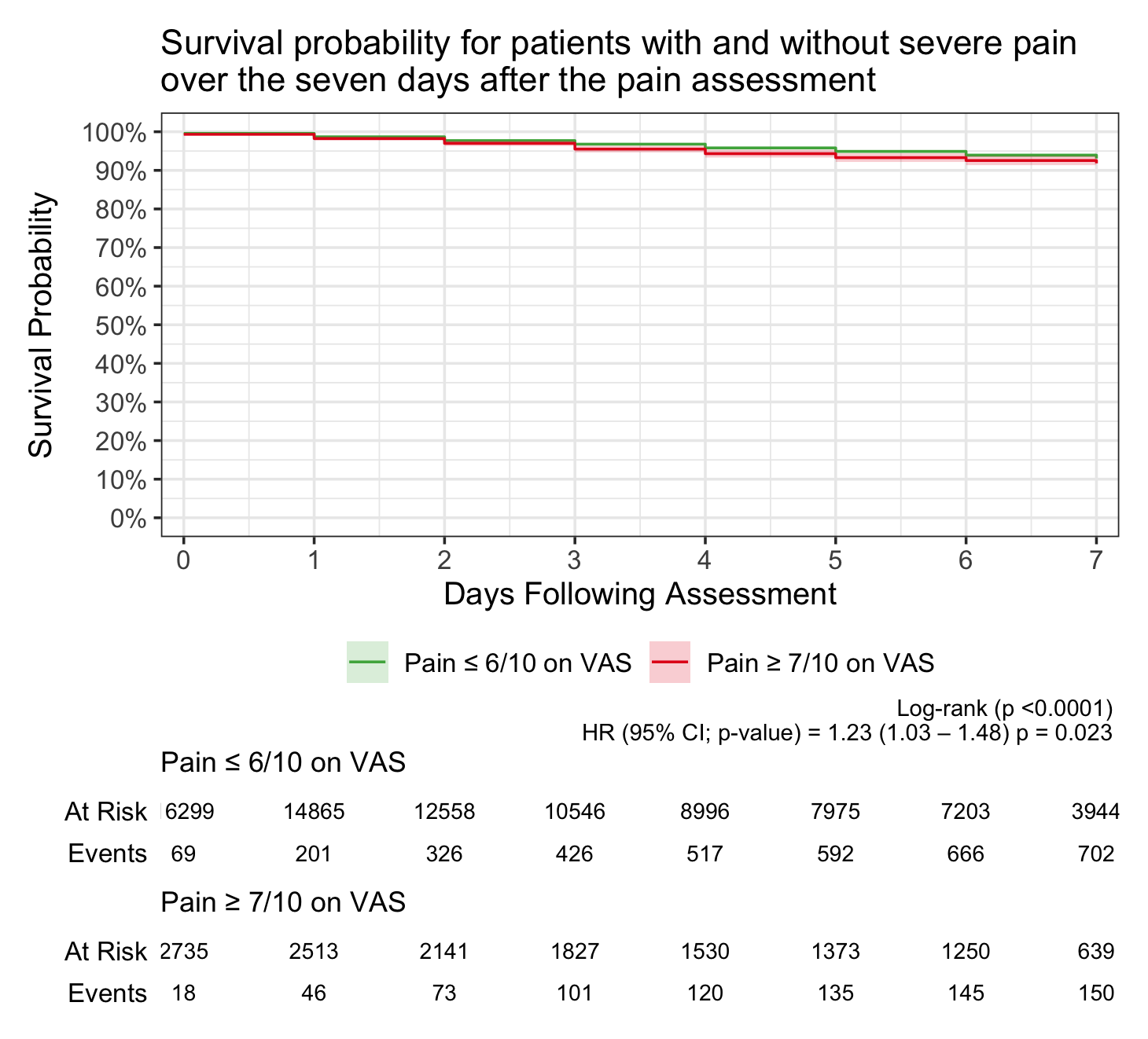

Figure 2: **Survival probability for patients with and without severe pain over the seven days after the pain assessment.** HR: Hazard ratio. Shaded bands show 95% confidence intervals (CI).

### Supplementary Material S15. Sensitivity analysis excluding the Nigerian and South African cohorts.

|  | **Outcome: critical illness** | | | **Outcome: in-hospital seven-day mortality** | | |
| --- | --- | --- | --- | --- | --- | --- |
|  | **Odds ratio** | **95% confidence interval** | **p value** | **Odds ratio** | **95% confidence interval** | **p value** |
| **Intercept (odds)** | 0.02 | 0.02 – 0.03 | **<0.0001** | 0.0009 | 0.005 – 0.001 | **<0.0001** |
| **Pain rating (0 – 10)** | 1.02 | 0.99 – 1.4 | 0.14 | 1.05 | 1.01 – 1.08 | **0**.**01** |
| **Age per 10 years** | 1.09 | 1.05 – 1.13 | **<0.0001** | 1.21 | 1.14 – 1.27 | **<0.0001** |
| **Pregnant** | 0.86 | 0.62 – 1.21 | 0.39 | - | - | **-** |
| **Known comorbidities** | | | | | | |
| Hypertension | 1.03 | 0.88 – 1.21 | 0.70 | - | - | **-** |
| Diabetes | 1.00 | 0.84 – 1.20 | 0.98 | - | - | **-** |
| Cancer | 1.31 | 1.02 – 1.68 | **0.03** | 2.19 | 1.57 – 3.04 | **<0.0001** |
| COPD/ Asthma | 2.08 | 1.65 – 2.63 | **<0.0001** | - | - | **-** |
| Heart disease | 1.62 | 1.31 – 1.20 | **<0.0001** | - | - | **-** |
| HIV/AIDS | 1.18 | 0.91 – 1.53 | 0.22 | 1.87 | 1.27 – 2.75 | **0.001** |
| Tuberculosis | 2.07 | 1.52 – 2.82 | **<0.0001** | - | - | **-** |
| Other | 1.25 | 1.07 – 1.47 | **0.006** | - | - | **-** |
| **Urgency of admission** | | | | | | |
| Emergency/ acute | 2.50 | 2.10 – 2.97 | **<0.0001** | 3.22 | 2.29 – 4.52 | **<0.0001** |
| Elective | Reference |  |  |  |  |  |
| **Main category for admission** | | | | | | |
| Infection | 1.62 | 1.18 – 2.23 | **0.003** | 2.97 | 1.59 – 5.55 | **0.0006** |
| Non-communicable disease | 2.09 | 1.56 – 2.79 | **<0.0001** | 5.45 | 3.06 – 9.71 | **<0.0001** |
| Trauma | 2.38 | 1.75 – 3.24 | **<0.0001** | 5.92 | 3.26 – 10.73 | **<0.0001** |
| Maternal health | Reference |  |  |  |  |  |
| Critical illness | - | - | **-** | 7.17 | 5.88 – 8.74 | **<0.0001** |

### Supplementary Material S16. Unadjusted three-level (nested) generalised logistic mixed model for sex and age predicting severe pain. P-values are rounded to 2 significant figures.

| Term | Odds Ratio | 95% CI | p-value |
| --- | --- | --- | --- |
| (Intercept) | 0.145 | 0.126; 0.167 | **<0.0001** |
| Sex (Male) | 0.969 | 0.890; 1.055 | 0.47 |
| Age | 0.894 | 0.844; 0.946 | **0.0001** |
| Sex (Male)*Age | 0.945 | 0.866; 1.031 | 0.20 |

### Supplementary Material S17. Unadjusted and adjusted three-level (nested) generalised logistic mixed model of the relationship between severe pain and critical illness status.

|  | **Unadjusted model** | | | **Adjusted model** | | |
| --- | --- | --- | --- | --- | --- | --- |
|  | **Odds ratio** | **95% confidence interval** | **p value** | **Odds ratio** | **95% confidence interval** | **p value** |
| **Intercept (odds)** | 0.12 | 0.11 – 0.13 | **<0.0001** | 0.02 | 0.02 – 0.03 | <**0.0001** |
| **Severe pain status (≥7/10)** | 1.25 | 1.10 – 1.41 | **0.0005** | 1.31 | 1.15 – 1.48 | <**0.0001** |
| **Age per 10 years** | 1.16 | 1.13 – 1.18 | **<0.0001** | 1.07 | 1.04 – 1.11 | <**0.0001** |
| **Pregnant** | 0.38 | 0.32 - 0.46 | **<0.0001** | 0.86 | 0.66 – 1.13 | 0.27 |
| **Known comorbidities** | | | | | | |
| Hypertension | 1.25 | 1.13 – 1.39 | **<0.0001** | 1.00 | 0.89 – 1.13 | 0.96 |
| Diabetes | 1.22 | 1.08 –1.39 | **0.0020** | 0.94 | 0.82 – 1.09 | 0.43 |
| Cancer | 1.22 | 1.02 – 1.46 | **0.026** | 1.25 | 1.03 – 1.51 | **0.019** |
| COPD/ Asthma | 2.70 | 2.27 – 3.23 | **<0.0001** | 2.12 | 1.76 – 2.55 | <**0.0001** |
| Heart disease | 2.08 | 1.79 – 2.43 | **<0.0001** | 1.60 | 1.36 – 1.88 | <**0.0001** |
| HIV/AIDS | 1.53 | 1.31 – 1.76 | **<0.0001** | 1.31 | 1.12 – 1.53 | **0.0008** |
| Tuberculosis | 2.93 | 2.41 – 3.57 | **<0.0001** | 2.07 | 1.67 – 2.56 | <**0.0001** |
| Other | 1.32 | 1.17 – 1.49 | **<0.0001** | 1.20 | 1.06 – 1.36 | **0.0046** |
| **Urgency of admission** | | | | | | |
| Emergency/ acute | 2.66 | 2.31 – 3.05 | **<0.0001** | 2.43 | 2.11 – 2.81 | <**0.0001** |
| Elective |  |  |  | Reference |  |  |
| **Main category for admission** | | | | | | |
| Infection | 1.80 | 1.48 – 2.18 | **<0.0001** | 1.30 | 1.00 – 1.68 | **0.046** |
| Non-communicable disease | 2.70 | 2.30 – 3.18 | **<0.0001** | 1.82 | 1.43 – 2.30 | <**0.0001** |
| Trauma | 3.76 | 3.15 – 4.48 | **<0.0001** | 2.13 | 1.66 – 2.73 | <**0.0001** |
| Maternal health |  |  |  | Reference |  |  |

### Supplementary Material S18: Unadjusted and covariate-adjusted three-level (nested) generalised logistic mixed model of the relationship between severe pain and in-hospital seven-day mortality

|  | **Unadjusted model** | | | **Adjusted model** | | |
| --- | --- | --- | --- | --- | --- | --- |
|  | **Odds ratio** | **95% confidence interval** | **p value** | **Odds ratio** | **95% confidence interval** | **p value** |
| **Intercept (odds)** | 0.04 | 0.03 – 0.04 | <**0.0001** | 0.005 | 0.003– 0.006 | <**0.0001** |
| **Severe pain status (≥7/10)** | 1.31 | 1.09 – 1.57 | **0.0045** | 1.27 | 1.04 – 1.56 | **0.020** |
| **Age per 10 years** | 1.38 | 1.33 – 1.43 | <**0.0001** | 1.23 | 1.18 – 1.29 | <**0.0001** |
| **Known chronic illness or pregnancy** | | | | | | |
| Cancer | 3.00 | 2.43 – 3.70 | <**0.0001** | 2.59 | 2.05 – 3.28 | <**0.0001** |
| HIV/AIDS | 1.56 | 1.26 – 1.94 | <**0.0001** | 1.39 | 1.09 – 1.76 | **0.0072** |
| **Urgency of admission** | | | | | | |
| Emergency/ acute | 4.17 | 3.21 – 5.43 | <**0.0001** | 3.25 | 2.47 – 4.28 | <**0.0001** |
| Elective |  |  |  | Reference |  |  |
| **Main category for admission** | | | | | | |
| Infection | 5.50 | 3.21 – 9.44 | <**0.0001** | 3.37 | 1.94 – 5.83 | <**0.0001** |
| Non-communicable disease | 14.51 | 8.84 – 23.81 | <**0.0001** | 6.31 | 3.78 – 10.54 | <**0.0001** |
| Trauma | 19.17 | 11.56 – 31.78 | <**0.0001** | 7.87 | 4.67 – 13.26 | <**0.0001** |
| Maternal health |  |  |  | Reference |  |  |
| **Critical illness** | | | | | | |
| Critically ill | 9.16 | 7.89 – 10.63 | <**0.0001** | 6.90 | 5.91 – 8.07 | <**0.0001** |
| Not critically ill |  |  |  | Reference |  |  |
